# Secrets of the hospital underbelly: patterns of abundance of antimicrobial resistance genes in hospital wastewater vary by specific antimicrobial and bacterial family

**DOI:** 10.1101/19006858

**Authors:** Meghan R. Perry, Hannah C. Lepper, Luke McNally, Bryan A. Wee, Patrick Munk, Amanda Warr, Barbara Moore, Pota Kalima, Carol Philip, Ana Maria de Roda Husman, Frank M. Aarestrup, Mark Woolhouse, Bram A.D. van Bunnik

## Abstract

**Background:** Hospital wastewater is a major source of antimicrobial resistance (AMR) outflow into the environment. This study uses metagenomics to study how hospital clinical activity impacts antimicrobial resistance genes (ARGs) abundances in hospital wastewater.

**Methods:** Sewage was collected over a 24-hour period from multiple wastewater collection points representing different specialties within a tertiary hospital site and simultaneously from community sewage works. High throughput shotgun sequencing was performed using Illumina HiSeq4000. ARG abundances were correlated to hospital antimicrobial usage (AMU), data on clinical activity and resistance prevalence in clinical isolates.

**Results:** Microbiota and ARG composition varied between collection points and overall ARG abundance was higher in hospital wastewater than in community influent. ARG and microbiota compositions were correlated (Procrustes analysis, *P*=0.014). Total antimicrobial usage was not associated with higher ARG abundance in wastewater. However, there was a small positive association between resistance genes and antimicrobial usage matched to ARG phenotype (IRR 1.11, CI 1.06 - 1.16, *P*<0.001). Furthermore, analysing carbapenem and vancomycin resistance separately indicated that counts of ARGs to these antimicrobials were positively associated with their increased usage (carbapenem rate ratio (RR) 1.91, 95% confidence intervals (CI) 1.01 – 3.72, *P*=0.07, and vancomycin RR 10.25, CI 2.32 – 49.10, *P*<0.01). Overall, ARG abundance within hospital wastewater did not reflect resistance patterns in clinical isolates from concurrent hospital inpatients. However, for clinical isolates of the family *Enterococcaceae* and *Staphylococcaceae*, there was a positive relationship with wastewater ARG abundance (odds ratio (OR) 1.62, CI 1.33 – 2.00, *P*<0.001, and OR 1.65, CI 1.21 – 2.30, *P*=0.006 respectively).

**Conclusions:** We found that the relationship between hospital wastewater ARGs and antimicrobial usage or clinical isolate resistance varies by specific antimicrobial and bacterial family studied. One explanation we consider is that relationships observed from multiple departments within a single hospital site will be detectable only for ARGs against parenteral antimicrobials uniquely used in the hospital setting. Our work highlights that using metagenomics to identify the full range of ARGs in hospital wastewater is a useful surveillance tool to monitor hospital ARG carriage and outflow and guide environmental policy on AMR.

## INTRODUCTION

In response to the antimicrobial resistance (AMR) crisis, a challenge for the research and medical communities is understanding the flow of AMR between different environmental niches (Woolhouse et al. 2015) and deciding where to focus surveillance and interventions to inform effective policies and action (Laxminarayan et al. 2016). There is an increasing interest in the contribution of hospital wastewater to AMR in the environment. Sewage treatment does not completely eradicate antimicrobial resistance genes (ARGs) and thus ARGs can enter the food chain through water and the use of sewage sludge in agriculture (Woolhouse and Ward 2013; Woolhouse et al. 2015). As a complex matrix representing human bodily waste the potential of community sewage as a surveillance tool to monitor the global epidemiology of AMR has recently been explored (Hendriksen et al. 2019; Aarestrup and Woolhouse 2020).

Hospitals are epidemiologically important nodal points for concentrated antimicrobial consumption and are sources of resistant pathogens (Versporten et al. 2018). Secondary care surveillance, guided by national and international policies, is based on passive reporting of phenotypic and molecular laboratory results for specific pathogens or from screening samples on specific high risk patients (Tornimbene et al. 2018; Department of Health and Social Care 2019). These methods do not represent the full impact of antimicrobial use and inpatient activity on AMR carriage within a hospital and thus risk of transmission. Nor do they capture all pertinent ARGs. As hospital wastewater contains inpatient bodily waste we hypothesised that it could be used as a representation of hospital inpatient carriage of AMR and as such may be a useful surveillance tool.

Many previous studies have identified key pathogens and resistant genes in hospital wastewater and attempts have been made to correlate resistance of specific organisms from hospital clinical isolates with hospital wastewater isolates with conflicting results (Tuméo et al. 2008; Drieux et al. 2016; Talebi et al. 2008; Yang et al. 2009; Maheshwari et al. 2016; Santoro, Romao, and Clementino 2012). There is currently a knowledge gap on how resistance in hospital wastewater quantitatively reflects clinical activity within hospitals. By applying the technique of metagenomics (Hendriksen et al. 2019) to obtain a universal view of ARG composition in hospital wastewater in this study we were able to interrogate this relationship in a multi-departmental study.

## MATERIALS AND METHODS

### Sewage collection and antibiotic residue analysis

Sampling was performed in June 2017 on eight wastewater collection points (CP), representing different clinical departments, identified to capture the effluent from the majority of the Western General Hospital, Edinburgh (Supplementary Figure 1). Using composite sampling machines, 100 mL of wastewater was sampled every 15 minutes over a 24-hour period thus aiming to collect a representative sample of waste from the hospital inpatient population. Simultaneously, a 24-hour time proportional sample was collected at the inflow site to Seafield community sewage works (hereafter “Seafield”), which serves a population equivalent of 760,000 from Edinburgh and the Lothians. Samples were transported from the site on dry ice and stored at -80°C. Antibiotic residue analysis was performed on 1L of composite hospital wastewaters and 1L of domestic sewage using LC-MS/MS as previously described (Berendsen et al. 2015; Hendriksen et al. 2019).

### DNA extraction and analysis

DNA was extracted from sewage using the QIAamp Fast DNA Stool mini kit with an optimized protocol as previously described(Knudsen et al. 2016) and sequenced on the HiSeq4000 platform (Illumina) using 2x 150bp paired-end sequencing. The taxonomic origin of paired reads were assigned using Kraken2 (Wood and Salzberg 2014) to the standard database, a database of representative bacterial genomes and a database of known vector sequences, UniVec_Core (downloaded 9^th^ April 2019). Taxonomic assignments were summarized at the genus level using kraken-biom (Dabdoub 2019). One sample, CP2, was heavily contaminated and removed from further analysis. We used KMA version 1.2.12 to assign the paired and singleton reads to a database consisting of ResFinder reference genes (downloaded 5th of September, 2019). The following flags were used: “-mem_mode -ef -1t1 -cge -nf -shm 1 -t 1” [20]. Reads mapping to the human reference genome (GCA_000001405.15) were removed prior to submission to public sequence databases according to the protocol used in the Human Microbiome Project (Human Microbiome Project 2021; Sherry 2011).

### Data collection

Data was collected on clinical isolates from the week surrounding the hospital wastewater sampling to represent pathogens in hospital inpatients. All types of clinical isolate were included but duplicate samples from the same patient within a 48-hour period were excluded. Antimicrobial usage was collated from weekly pharmacy issues to each ward over the 3 months prior to sampling and presented as defined daily dose per 100 occupied bed days (DDD/100OBDs). Pharmacy issues for prescriptions for outpatient use and for theatres were excluded.

### Data analysis

All statistical analysis and plots were produced using R version 3.6.0. The abundance of ARGs and bacterial genera were calculated as Fragments Per Kilobase of transcript per Million mapped bacterial reads (FPKM) (Munk et al. 2018) Bray-Curtis dissimilarity matrices were determined using Hellinger transformation of the FPKM. Resistance genes from the ResFinder database were grouped into clusters with 90% sequence homology. The top 50 ARGs were visualised using a heatmap and gene-wise and collection point dendrograms as previously described (Hendriksen et al. 2019). Procrustes analysis was used to test the association between the resistome and bacteriome dissimilarities.

### Correlation between inpatient activity and ARG abundance

The source of variance in the abundance of ARGs between the collection points was investigated using a multilevel Poisson model with the dependent variable as counts of ARG reads at each collection point aggregated at the 90% homology cluster level. We used an offset term with the log of the average gene-length per cluster in the ResFinder database, multiplied by the total bacterial reads per collection point. Random effects of collection point, 70% sequence homology cluster, and observation were included in the model, the latter to model the over dispersion inherent to count data (Harrison 2014).

In the main model, we accounted for co- and cross-resistance by fitting both a measure of direct selection for resistance (effect of department-level usage of antimicrobials on ARGs that confer resistance to those antimicrobials) and indirect selection (effect of total department-level AMU on ARG abundance). In a second set of three models we tested the association between resistance genes and antimicrobial usage of three specific antimicrobials of interest chosen to represent parenteral antimicrobials only used in a hospital setting (carbapenems, vancomycin) and an antimicrobial widely used in both community and hospital (amoxicillin). We use a Bonferroni correction on P values of these additional tests to account for increased risk of type I error. We used all antimicrobial resistance phenotypes suggested for any gene in a 90% homology cluster from either the ResFinder or STARAMR (National Microbiology Laboratory 2021) databases. The average length of stay per department was also used to assess the role of clinical activity on sewage resistance abundance in the main model. The fixed effects structure of the main model was further adjusted using AIC minimising methods, assessing whether any interaction effect should be included.

To assess the relationship between AMR in clinical isolates and ARG abundance in hospital wastewater a binomial generalised linear mixed effects model was used including random effects for site, the class of the antimicrobial used to test the isolates, and for the species of the isolate to control for inter-species heterogeneity. Two fixed effects were estimated for the log FPKM of all resistance genes in the sewage that had the same resistance phenotype as the isolates: one for isolates that were urinary or faecal, and a second for all other isolate types, due to the different dynamics of inpatient bodily waste being represented in the wastewater system. Using separate binomial regression models, we accounted for heterogeneity between the taxonomic family of the isolates in the relationship between AMR in clinical isolates and sewage ARGs. As some families were rarely tested, the sample size was too small for this heterogeneity to be assessed in a single model. Therefore, the three most frequently isolated families were assessed (*Enterobacteriaceae, Enterococcaceae,* and *Staphylococcaceae*), with the log FPKM of phenotypically matched resistance genes as the only model effect. A Bonferroni correction was used to adjust the P values of the effects of these models to account for multiple testing. A similar model was used to evaluate the relationship between AMU and AMR in clinical isolates.

### Ethics

This study was conducted following approval from NHS Lothian Research and Development committee under the sponsorship of University of Edinburgh. There was no direct patient contact and therefore the study did not require ethical board approval.

## RESULTS

The hospital departments served by the wastewater collection points differed by pattern of antimicrobial use (Table 1, S2) and resistance in the 181 clinical isolates identified in the week surrounding wastewater sampling (Figure S3).

**Table 1.** Demographics of hospital collection points. Standard deviation only represents standard deviation of the average age and length of stay per week. Antimicrobial usage from previous three months does not include antibiotics issued for outpatient prescriptions or in theatres. Clinical isolates are from inpatients in the week surrounding wastewater collection. Abbreviations: pts=patients, DDD=defined daily dose, OBDs=occupied bed days, s.d.=standard deviation.

| Collection Point | Specialties | No. of wards | No. of pts | Average length of stay in days (s.d.) | Average age in years (s.d.) | DDD per 100 OBDs | No. of clinical isolates |
| --- | --- | --- | --- | --- | --- | --- | --- |
| CP1 | Cardiology, Urology | 3 | 46 | 4.9 (0.8) | 62.6 (2.2) | 123.7 | 19 |
| CP3 | Oncology, Haematology | 7 | 67 | 3.7 (2.7) | 62.1 (1.0) | 200.5 | 27 |
| CP4 | Acute receiving unit | 5 | 35 | 0.9 (0.7) | 70.5 (2.3) | 325.8 | 45 |
| CP5 | Neuroscience | 3 | 59 | 3.3 (1.1) | 53.5 (2.1) | 73.5 | 8 |
| CP6 | Intensive care, Surgery, Medicine | 3 | 70 | 7.6 (2.5) | 66.6 (1.7) | 223.8 | 17 |
| CP7 | Infectious Diseases, Surgery, Medicine | 6 | 105 | 6.1 (3.2) | 63.5 (0.8) | 148.1 | 20 |
| CP8 | Respiratory, Medicine for the Elderly, Urology, Surgical High Dependency | 6 | 133 | 12.8 (9.0) | 69.0 (1.0) | 116.4 | 25 |

### Metagenomics of wastewater

An average read pair count of 38.4 million (range 35.7-39.2 million) was obtained with an average of 62% (range 52-73%) of reads allocated to bacteria from the seven hospital wastewater samples and one community sewage sample (https://www.ebi.ac.uk/ena/data/view/PRJEB34410). An average of 0.25% of reads mapped to ARGs in the seven hospital wastewater samples versus 0.1% from Seafield (Table S1).

One thousand, one hundred and fifty-four unique bacterial genera were detected across all samples (range 1151 - 1154 genera per sample, Table S2). The top nineteen genera accounted for >70% of bacterial abundance in all samples (Figure 1.D). The most predominant genera were *Pseudomonas* and *Acinetobacter*, mainly environmental species such as *Pseudomonas fluorescens, Acinetobacter johnsonii*, likely representing bacteria usually present in the hospital pipes. When compared with Seafield, there was a difference in diversity in the hospital samples with a higher predominance of gut associated bacteria including *Faecalibacterium, Bacteroides, Bifidobacterium* and *Escherichia*. (Figure 1.B & D).

**Figure 1.**
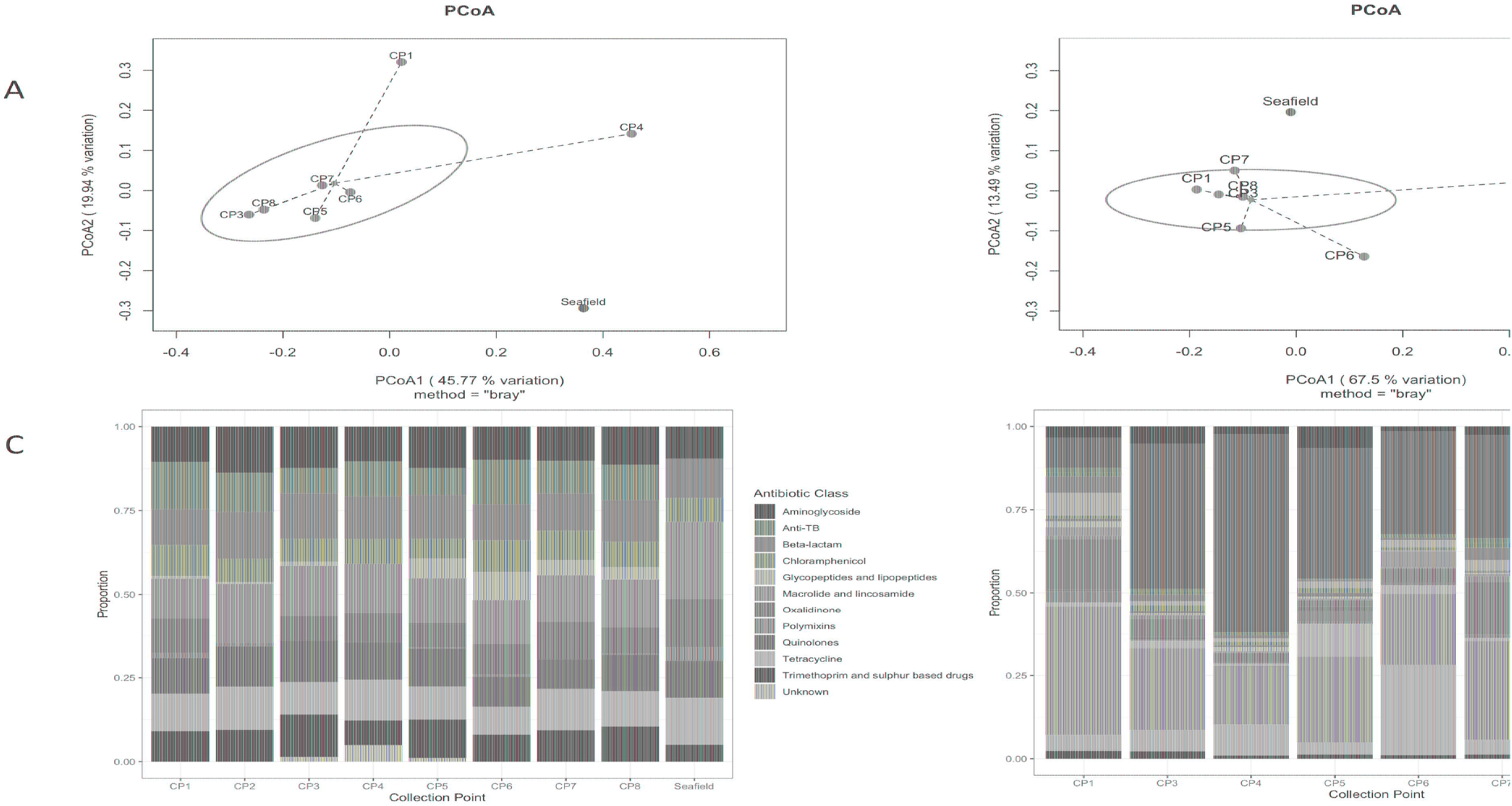
Hospital wastewater and community sewage resistome and microbiome abundance composition. A) Principal coordinate analyses of resistome based on Bray-Curtis dissimilarity. The percentage of variation explained is noted on the axis labels. B) Principal coordinate analyses for the microbiome. C) Relative abundance of ARGs by antimicrobial class. D) Relative abundance of the 19 most abundant bacterial genera in the wastewater and sewage microbiome. Abbreviations: CP=collection point within hospital, Seafield=community sewage works, TB=tuberculosis.

ARG abundance and composition varied across different hospital collection points and Seafield (Figures 1.A & C, Figure 2, Figures S4 & S6). Apart from the wastewater collected at CP4 which represents the acute receiving unit with patients directly admitted from the community, ARG abundance from hospital wastewater was higher than ARG abundance in Seafield (Figure 2, Fig S4). ARG composition was strongly correlated with bacterial genus level composition (Procrustes, p=0.014) (Supplementary Figure 5).

**Figure 2.**
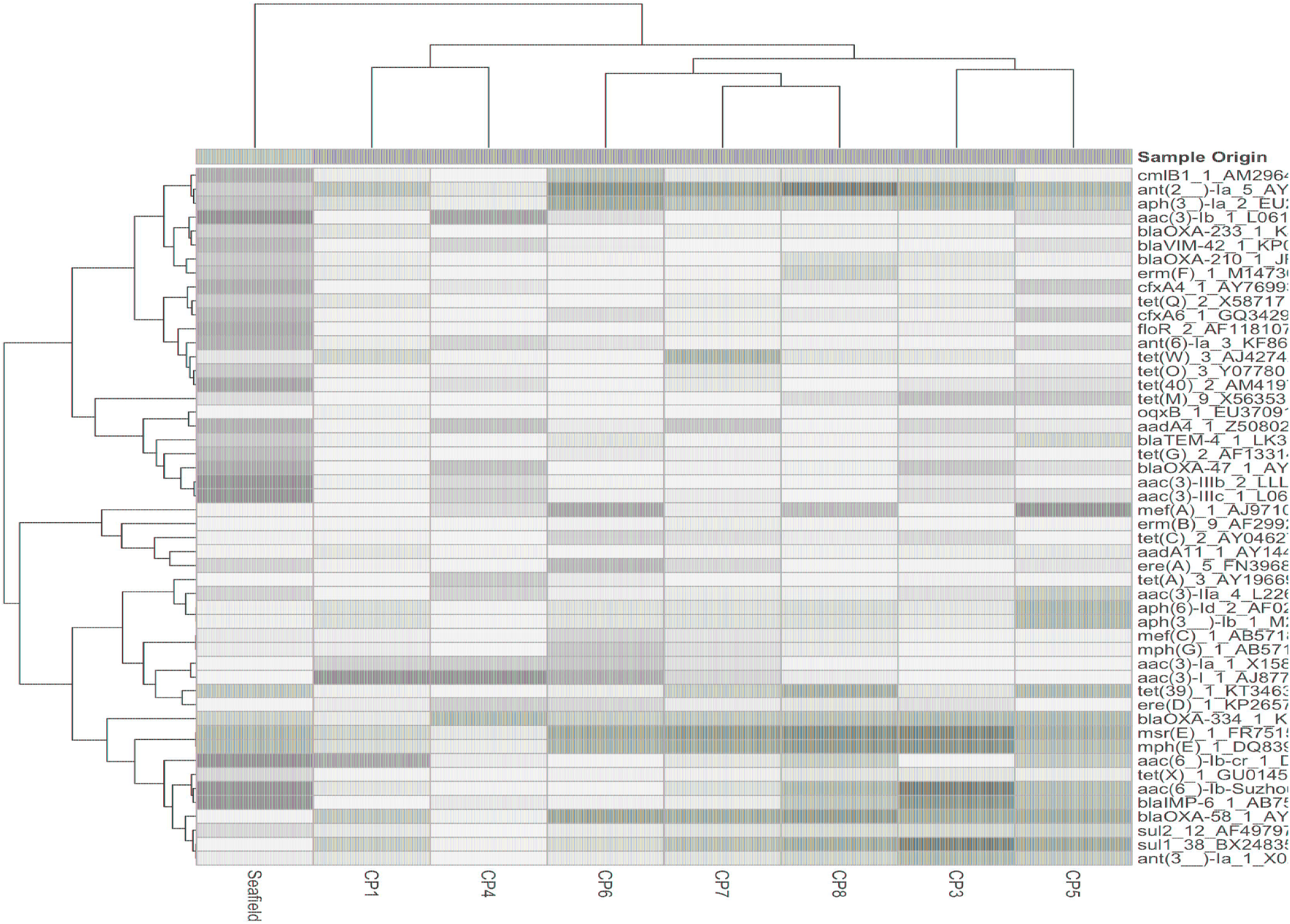
Heat map of 50 most abundant antimicrobial resistance genes (ARGs). Relative abundance of ARGs (FPKM) were log transformed and both ARGs and collection points were clustered using complete-linkage clustering. For ARGs clustering was based on Pearson correlation coefficients, for collection points clustering was based on the BC-dissimilarity matrix (Figure 1) which uses all genes.

We detected 502 different resistance genes belonging to ten different antimicrobial classes (Table S3) but over 65% of the sample resistomes were composed of the 15 most abundant genes (Figure S6), mainly belonging to the aminoglycoside and macrolide antimicrobial classes (Figure 1.C). Key ARGs of interest to infection control including *bla*OXA, *bla*IMP and genes of the *vanA* cluster were identified.

### Inpatient activity and ARG abundance

No significant relationships were observed between total antimicrobial usage or length of stay and the abundance of ARGs in sewage (Fig. 3, Table S5). This result indicates there was no evidence for indirect selection or for the impact of transmission among hospital patients on ARG abundance in sewage when all resistance phenotypes were modelled. There was a significant positive effect of increased phenotypically-matched antimicrobial usage on resistance gene abundance, indicating support for a small role of direct selection (IRR 1.11, CI 1.06 - 1.16, *P* < 0.001). AIC comparison of fixed effect structures for the model indicated that no interaction effects improved model fit.

**Figure 3.**
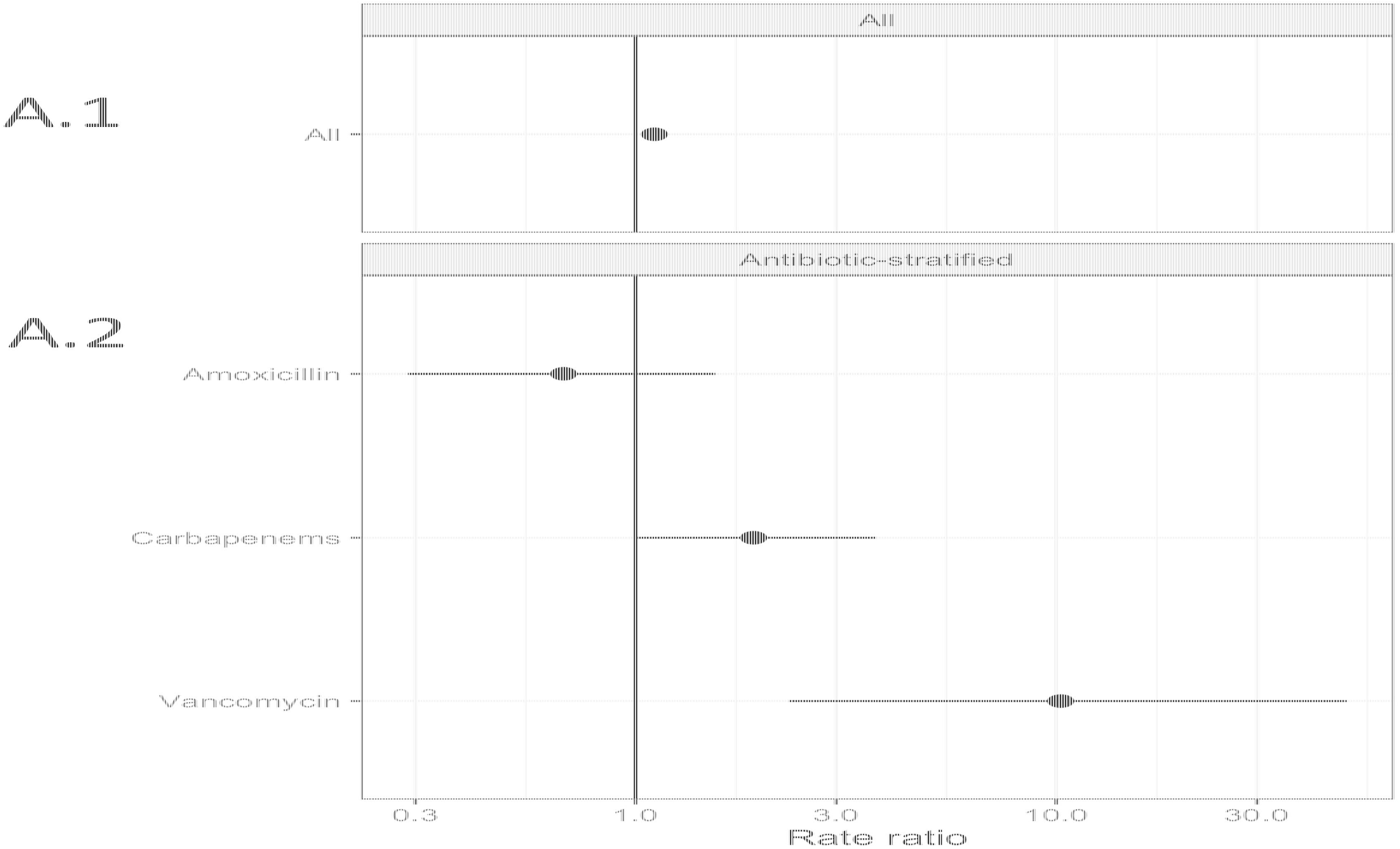
**Generalised linear mixed effects models for the relationship between antimicrobial resistance gene abundance, hospital department antibiotic consumption rates, and hospital department rates of resistance in clinical isolates.** A) Effect of antimicrobial usage (AMU) measured in defined daily dose per 100 occupied bed days (DDD/100 OBDs) on antimicrobial resistance gene (ARG) abundance A.1.) The main model, with a single coefficient for all resistance phenotypes. A.2.) Separate models with coefficients for each antimicrobial. B) Association between antimicrobial resistance gene abundance in the sewage and clinical resistance rates. B.1.) Main model, with a single coefficient for all clinical isolate taxonomic family, stratified by sample type - urine or faecal samples (All: Urine), and for resistance genes and any other sample source (All: Other). B.2.) Separate models with coefficients for each isolate taxonomic family.

We next analysed data on the association between carbapenem, vancomycin and amoxicillin usage and ARGs conferring resistance to these specific antimicrobials in 3 separate models (Fig 3A, Table S5). We found positive associations that were significant between vancomycin ARGs and vancomycin usage (IRR 10.25, CI 2.32 – 49.10, *P* < 0.001) and showed a trend towards significance between carbapenem ARG abundance and carbapenem antimicrobial usage (IRR 1.91, CI 1.01 – 3.72, *P* = 0.07). No evidence for an association between amoxicillin usage and amoxicillin ARGs was identified. We omitted the observation-level random effect from vancomycin model due to singular model fits, so overdispersion was not accounted for.

ARG abundance at a class level within hospital wastewater did not reflect resistance patterns in clinical isolates when all the data was analysed in one model (Fig 3B, Table S6). There was no difference between the relationship of isolates from urine and faecal samples with ARG abundance and isolates from other sample types, e.g. skin, which we expect to enter the wastewater system at different rates via sinks and showers. We next separately modelled the three most frequently isolated taxonomic families (Fig 3, Table S6). *Enterococcaceae* and *Staphylococcaceae* had a significant positive association with the abundance of ARGs conferring resistance to the same antimicrobial class (OR: 1.62, C.I. 1.32 – 2.00, p < 0.001, and OR: 1.65, C.I. 1.21 – 2.30, p < 0.01, respectively), but there was no such relationship for resistance levels in Enterobacteriaceae. At an antimicrobial class level, clinical isolate resistance did not reflect the antimicrobial usage of that class in the preceding 3 months (Supplementary Table 4).

Analysis of antibiotic residues reflected the high AMU within the hospital compared to the community with an average 12-fold increased residue concentration in hospital effluent (ranging between 4 and 13 μL^-1^) for the five classes measured (Supplementary Figure 7). Our residue data only represents the residue levels from the whole hospital and not individual collection points and thus could not be specifically correlated with ARG abundance.

## DISCUSSION

This study identified that hospital AMU impacts ARG abundances in hospital effluent, with implications upstream for infection control in the hospital and downstream for AMR in the environment. Overall, the distribution of bacterial genera and ARGs in our hospital wastewater samples and domestic sewage sample is similar to previously described sewage composition in European regions (Hendriksen et al. 2019; Buelow et al. 2018).

There was a significant positive relationship between inpatient department-level AMU and the abundance of antimicrobial resistance phenotype matched ARGs when all data was considered together. No relationship was found for total department AMU and ARG abundance. This supports a role of direct selection from antimicrobial usage in overall patterns of ARGs in hospital waste water, but not for indirect selection. Previous studies have found a relationship at a country level between antimicrobial residues and ARG abundance in sewage from the community (Hendriksen et al. 2019). Indeed, our data shows that the hospital antimicrobial residues are within the minimum selection concentration range for *Escherichia coli* and ciprofloxacin resistance (Sandegren 2014), although recent work suggests that higher antimicrobial concentrations are needed to select for resistance in microbial communities such as sewage (Klümper et al. 2019).

The association between phenotype-matched ARGs and AMU was weak. Sewage captures resistance acquired in both the community and in the hospital, but drivers of hospital- and community-acquired resistance differ. For example, amoxicillin is used in both the community and hospitals, and resistance is widespread in the UK (60% hospital isolates resistant to amoxicillin or ampicillin in 2019) (European Centre for Disease Prevention and Control 2020), suggesting patients commonly arrive in hospital with carriage of amoxicillin resistance genes. The acquisition of vancomycin or carbapenem resistance, on the other hand, is associated with prior use of these antibiotics in hospital (Vasilakopoulou et al. 2020; Zhao et al. 2020), and these antibiotics are solely used parenterally in a hospital setting. Factors affecting within-hospital selection for and transmission of resistance, such as hospital antimicrobial usage, may play a stronger role in patterns of ARGs of vancomycin and carbapenems in hospital waste water than the ubiquitously used antibiotic amoxicillin. In support of this theory, we found a positive relationship between AMU and waste water ARGs for vancomycin and carbapenems, but not amoxicillin. Where a particular ward or department consumes high levels of carbapenem or vancomycin then this work demonstrates that there could be high levels of undetected faecal or urinary carriage of carbapenem and vancomycin resistance genes. This could warrant more stringent isolation of these patients, in fitting with concerns about “unsampled transmission chains” in carbapenem-resistant *Enterobacteriaceae* (Cerqueira et al. 2017). In addition, if the 70% renal excretion of unchanged meropenem (Mouton and van den Anker 1995) selects for resistant organisms in waste water, then procedures for treatment of the bodily waste of patients on meropenem may need to be reconsidered.

Length of stay did not impact ARG abundance in this dataset, despite prolonged duration of inpatient stay being a risk factor for carriage and infection with resistant microorganisms in previous studies (Safdar and Maki 2002; Gupta et al. 2011; Founou, Founou, and Essack 2018). This appears not to support the theory of transmission of antimicrobial resistant organisms amongst patients and their local environment, including from the hospital water system (Kotay et al. 2017), during their inpatient stay. However, as these data were aggregated at the department-level there were few observations of length of stay, and further research with a greater sample size is needed to investigate this relationship.

Metagenomics can capture ARGs carried by a wide variety of bacterial genera, which is of benefit as the majority of ARGs are carried by non-pathogenic commensal bacteria (Sommer, Dantas, and Church 2009). Although short-read sequencing cannot conclusively resolve associations between bacteria and ARGs, in our results ARGs are highly correlated with the bacteria identified at that collection point (Supplementary Fig 7). This can explain why levels of ARGs for aminoglycosides, tetracyclines and macrolides are higher than levels of phenotypic resistance in clinical isolates; the composition of bacterial genera within wastewater may have intrinsic or high levels of resistance to these antimicrobial classes. The potential for transfer of ARGs within the sewage network onto and between human pathogens has been demonstrated indicating the benefit of obtaining a universal view of ARGs (Ludden et al. 2017).

No quantitative relationship was observed between clinical isolates and ARG abundance in hospital wastewater when all data was considered together. In addition there was no relationship between AMU in the previous three months and resistance in clinical isolates. This may be because clinical isolates are not representative enough of carriage of resistance in the inpatient population as there is a low rate of culture positivity. However, when examined separately, there was a positive relationship between resistance in *Enterococcaceae* or *Staphylococcaceae,* but not *Enterobactericeae,* and hospital wastewater ARG abundance. The literature on these relationships is divided (Tuméo et al. 2008; Talebi et al. 2008; Yang et al. 2009; Ory et al. 2016; Hutinel et al. 2019; Zarfel et al. 2013) and future work on antimicrobial usage, specific organisms, isolate types and ARG abundance in sewage potentially over a longer time period is required to interrogate these relationships further (Mladenovic-Antic et al. 2016; Rogues et al. 2007).

There was a higher abundance of ARGs in all hospital wastewater samples, bar one (CP4) which represents acute admissions unit, compared to Seafield. The lower abundance in Seafield could be due to dilution, and a decline in the relative abundance of AMR-gene carrying human commensal bacteria in the environment of sewerage system (Pehrsson et al. 2016), or possibly lower exposure to antimicrobial residues in community waste water. Associations between antimicrobial residues in community waste water and ARGs have been found (Hendriksen et al. 2019; Ju et al. 2019), and hospital waste water has been previously shown to have higher antimicrobial residue levels (Booth, Aga, and Wester 2020). Some studies comparing sewage influent in paired communities with and without a hospital have found minimal effect of a hospital on community influent (Gouliouris et al. 2019; Buelow et al. 2018). In other work, comparing resistance in hospital and community waste water has indicated some associations (Ludden et al. 2017; Rogues et al. 2007; Pehrsson et al. 2016), although not all studies making this comparison have found evidence for a relationship (Paulshus et al. 2019).

Concern has been raised about the impact of hospital wastewater on urban influent and effluent and specific water treatments for hospital wastewater have been called for. This work highlights that physicians could consider prescribing environmentally degradable antimicrobials such as beta-lactams over antimicrobials which have persistent residues across environmental niches e.g. tetracycline to minimise the impact of antimicrobials on the environmental resistome (Wellington et al. 2013). The ultimate effect of environmental ARGs on human disease is an ongoing important research question (Bürgmann et al. 2018).

The use of metagenomics is a key strength of this study, allowing quantification of resistance genes to a wide range of antibiotics and retrospective investigation if new resistance genes emerge. The 24-hour composite samplers provide a representative sample of the hospital (Chau et al. 2020), although hospital staff, outpatients and visitors will have also contributed to the effluent. In addition, some patients will have moved around the hospital during the sampling period. Although this study is limited to one hospital site at one time point the variation in antimicrobial use and inpatient characteristics in each department has allowed us to treat them as discrete treatment centres and draw conclusions about factors affecting ARG abundance.

There is little doubt that hospital resistant pathogens can be abundant in wastewater systems (Gouliouris et al. 2019; Ludden et al. 2017; Maheshwari et al. 2016). However, using metagenomic sequencing we show that resistance in hospital wastewater may quantitatively reflect clinical isolate resistance for some bacterial species (*Enterococcaceae* and *Staphylococcaceae*), although not all. As a surveillance tool this novel technique can represent the burden of AMR carriage in hospital inpatients and hospital pipes for specific resistance genes relating to important parenteral antimicrobials such as carbapenems and vancomycin. It may also aid in identification of emerging patterns of ARG abundance and novel ARGs, and how they may relate to changing patterns of transmission, infection control policies and antimicrobial usage. Further longitudinal work evaluating the wastewater from multiple hospital sites is needed to establish AMU/ARG relationships, optimal collection points and sampling methods to be able to develop this as a surveillance technique.

In conclusion, we show in a multi-departmental study that the relationships between ARG abundance in hospital wastewater and hospital AMU or clinical resistance levels may vary by antimicrobial type and bacterial species. Our study emphasises in a novel way the ARG burden from the high antimicrobial consuming and high resistance carriage environment of the hospital and that promoting active antimicrobial stewardship, particularly of key parenteral antimicrobials such as carbapenems and vancomycin, would impact the burden of environmental AMR. Hospital wastewater is an important source of AMR into the environment; this should be considered in environmental policy to reduce the flow of AMR between different environmental reservoirs.

## Data Availability

Raw reads will be available in a public accessible database

## Acknowledgements

The authors would like to thank NHS Lothian estates and Scottish Water for their help with the sampling and Mick Watson for his laboratory support.

## Author contributions

MRP conceived the project and developed it with input from BvB, HL, FA and MW. MRP facilitated sampling and DNA extraction with AW. PK, CP and BM provided clinical and pharmaceutical databases and input. ARH performed antibiotic residue analysis. Bio-informatics and analyses by HL, BvB, LMcN, BW, PM and MRP with input from FA and MW. Manuscript drafted by MRP and HL with input from BVB and review and comments from all authors.

## STATEMENT OF CONTRIBUTION TO THE FIELD

Sewage is an attractive medium for surveillance of antimicrobial resistance (AMR). In this study we interrogate the contribution of hospitals, as focal points of antimicrobial usage and bacterial infections, to resistance in urban waste water. Previous studies have compared resistance in hospital patients and their sewage effluent, but focus on only a single (or a few) bacterial species and antimicrobials, therefore insufficiently addressing the diverse microbiome(s) and resistome(s) in hospitals and wastewater.

In this study, we apply metagenomics to hospital wastewater and investigate the relationship between the abundance of antimicrobial resistance genes (ARGs) in the sewage and clinical activity. Clinical activity included antimicrobial usage and resistance in disease-causing bacteria cultured from inpatients. This innovative analysis of sewage allows quantification of both the full range of ARGs to all antimicrobials and specific ARGs of clinical interest.

We demonstrate variation between hospital departments in the abundance in ARGs in the sewage which reflected differences in inpatients’ resistant bacteria and antimicrobial usage. Furthermore we show that the relationship between clinical activity and ARGs in wastewater vary by resistance type and bacterial species. We suggest that detection of these relationships is driven by ARGs to antibiotics only used in the hospital setting.

