## supplementary material for "Secrets of the hospital underbelly: patterns of abundance of antimicrobial resistance genes in hospital wastewater vary by specific antimicrobial and bacterial family"

**Supplementary tables**

| Name | All read pairs | Non-human read pairs | % of total reads | Bacterial read pairs | % of non-human reads | Viral read pairs | % of non-human reads | Unclassified read pairs | % of non-human reads | AMR gene read pairs | % of non-human reads |
|---|---|---|---|---|---|---|---|---|---|---|---|
| CP1 | 38869360 | 37007788 | 95 | 20284420 | 52 | 36350 | 0.09 | 19414692 | 50 | 57318 | 0.15 |
| CP3 | 38950823 | 37111066 | 95 | 23077786 | 59 | 7052 | 0.02 | 15802719 | 41 | 153806 | 0.39 |
| CP4 | 38854316 | 38656944 | 99 | 27184108 | 70 | 2945 | 0.01 | 12865522 | 33 | 27632 | 0.07 |
| CP5 | 39062827 | 38294655 | 98 | 27458178 | 70 | 8784 | 0.02 | 12632702 | 32 | 123020 | 0.31 |
| CP6 | 38503994 | 28249480 | 73 | 21423749 | 56 | 30321 | 0.08 | 8045607 | 21 | 73101 | 0.19 |
| CP7 | 39176215 | 37801241 | 96 | 21622738 | 55 | 5525 | 0.01 | 18844242 | 48 | 102444 | 0.26 |
| CP8 | 35716605 | 35093020 | 98 | 22709754 | 64 | 6201 | 0.02 | 14292670 | 40 | 132598 | 0.37 |
| Seafield | 38298451 | 38143462 | 100 | 28016716 | 73 | 3594 | 0.01 | 11196189 | 29 | 40977 | 0.11 |

**Table S1. Total read pairs, read pairs assigned to major taxonomic groups and antimicrobial resistance gene read pairs.**

**Table S2 Bacterial genera at each collection point**

Please see separate excel file

**Table S3 AMR genes at each collection point (FPKM)**

Please see separate excel file

| | Clinical Isolate Model | | |
|---|---|---|---|
| *Coefficients* | *Odds Ratios* | *Conf. Int. (95%)* | *P-Value* |
| Intercept | 0.02 | 0.00 – 0.11 | **<0.001** |
| AMU (log, urine/faecal samples) | 0.92 | 0.64 – 1.31 | 0.629 |
| AMU (log, non-urine/faecal samples) | 1.27 | 0.85 – 1.89 | 0.245 |
| Isolate Type = Urine | 8.05 | 2.69 – 24.15 | **<0.001** |
| **Random Effects** | | | |
| $\sigma^2$ | 3.29 | | |
| $\tau_{00}$ Observation | 1.82 | | |
| $\tau_{00}$ Organism | 3.40 | | |
| $\tau_{00}$ Antibiotic Class | 4.04 | | |
| $\tau_{00}$ Site | 0.01 | | |
| ICC Observation | 0.14 | | |
| ICC Organism | 0.27 | | |
| ICC Antibiotic Class | 0.32 | | |
| ICC Site | 0.00 | | |
| Observations | 1124 | | |
| Marginal $R^2$ / Conditional $R^2$ | 0.034 / 0.607 | | |

**Table S4. Output from model examining the relationship between resistance in clinical isolates and antimicrobial usage (AMU)**

| Data subset | Number of gene cluster groups | Number of observations | Fixed effects | | | Random effects | | |
|---|---|---|---|---|---|---|---|---|
| | | | **Variable** | **IRR (CI)** | **P value** | $\tau_{Observation}$ | $\tau_{Cluster}$ | $\tau_{Site}$ |
| All data | 106 | 584 | Log of total DDDs | 0.87 (0.43 - 1.73) | 0.64 | 1.21 | 2.61 | 0.13 |
| | | | Log of phenotype-matched DDDs | 1.11, (1.06 - 1.16) | <0.001 | | | |
| | | | Average length of stay | 1.06 (0.97 - 1.17) | 0.11 | | | |
| Carbapenems | 12 | 56 | Log of phenotype-matched DDDs | 1.91 (1.01 – 3.72) | 0.07† | 2.01 | 2.51 | 0.13 |
| Vancomycin | 3 | 11 | Log of phenotype-matched DDDs | 10.25 (2.32 – 49.10) | <0.001† | - | 0.46 | 1.88 |
| Amoxicillin | 42 | 172 | Log of phenotype-matched DDDs | 0.68 (0.29 – 1.55) | 0.84† | 1.51 | 2.42 | 0.12 |

**Table S5. Data summary and output from models examining the relationship between antimicrobial usage (AMU) and antimicrobial resistance gene abundance in waste water. Each row represents a separate model. Abbreviations: DDDs=daily defined doses, CI=confidence interval.** † P values Bonferroni corrected

| Data subset | Number of bacterial species | Number of observations | Fixed effects | | | Random effects | | |
|---|---|---|---|---|---|---|---|---|
| | | | Variable | Odds Ratios (CI) | P value | $\tau_{Observation}$ | $\tau_{Class}$ | $\tau_{Organism}$ |
| All data | 28 | 2595 | Urine or faecal samples: Log sewage FPKM<br><br>Other sample sources: log of sewage FPKM | 1.13 (0.90 – 1.42)<br>0.88 (0.69 – 1.12) | 0.281<br>0.293 | 1.84 | 3.87 | 3.54 |
| *Enterococcaceae* | 3 | 201 | Log sewage FPKM | 1.62 (1.33 – 2.00) | <0.001† | - | - | - |
| *Staphylococcaceae* | 5 | 224 | Log sewage FPKM | 1.65 (1.21 – 2.30) | 0.006 † | - | - | - |
| *Enterobacteriaceae* | 9 | 395 | Log sewage FPKM | 0.92 (0.79 – 1.01) | 0.92 † | - | - | - |

**Table S6. Summary of data and output of models examining the relationship between clinical isolates and ARG abundance in waste water. ORs are for the log FPKM of ARGs matched to the antimicrobial class of the resistance test of the isolate. The three bacterial families were modelled separately.** † P values Bonferroni corrected

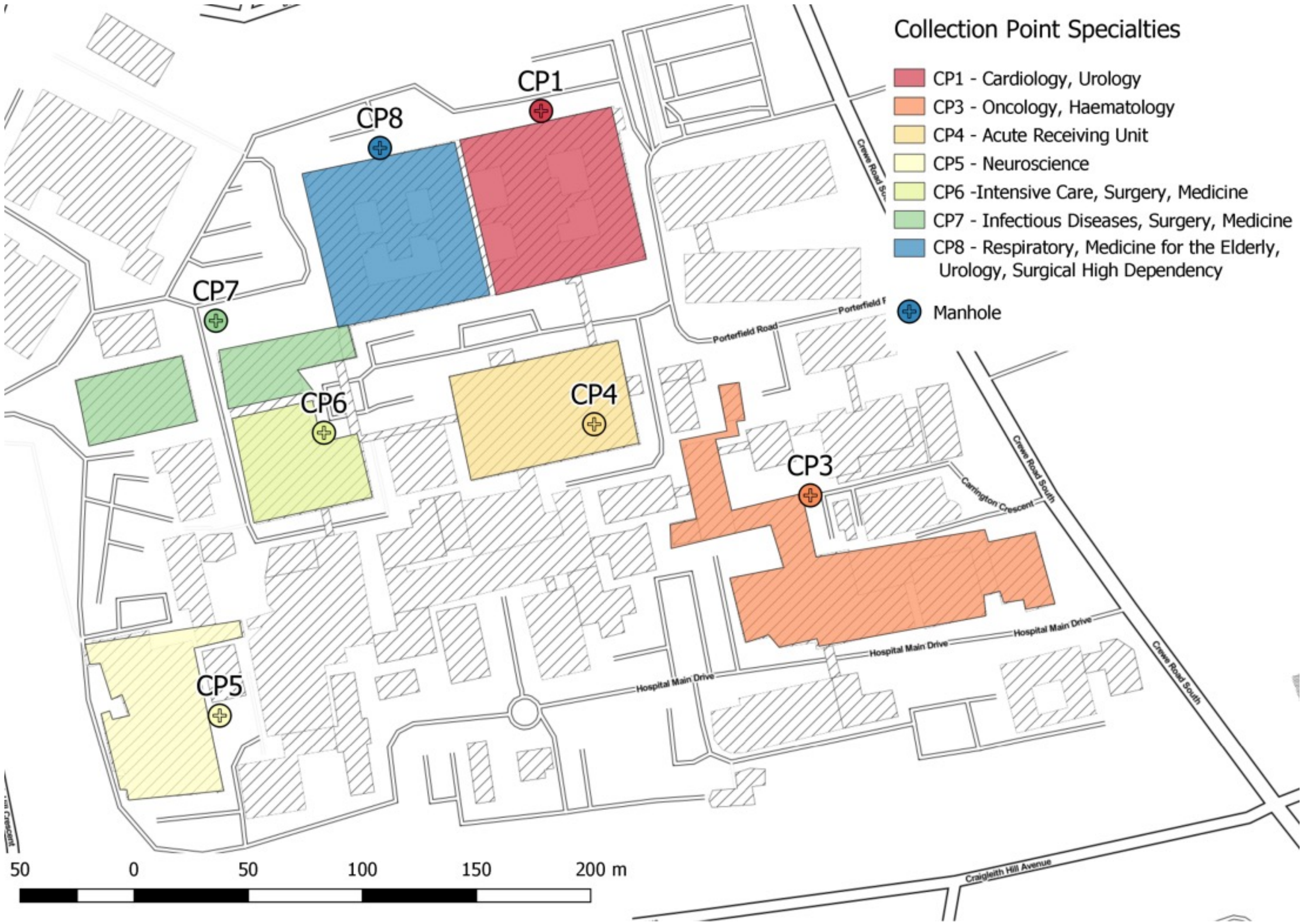


**Supplementary Figure 1** Schemata of location of analysed wastewater Collection Points (CP) on tertiary hospital site Hospital wastewater was sampled from indicated locations, which are the main clinical areas of the hospital site, over a 24-hour period using composite samplers. Shaded areas represent the drainage of the sampled manholes. CP4 was a manhole located underneath the hospital in a plant room.

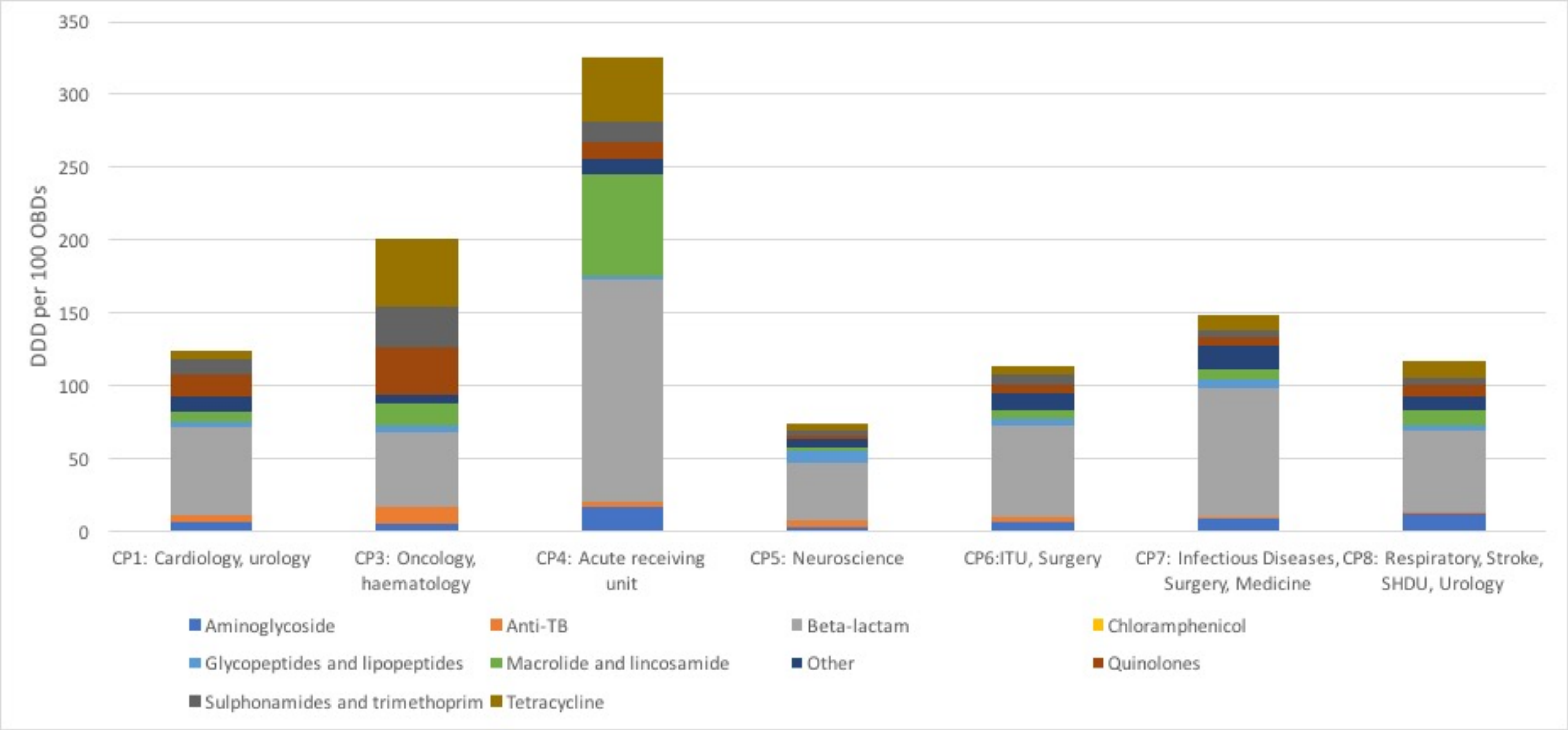


**Supplementary Figure 2** Antimicrobial consumption by class over different hospital departments

Pharmacy issues to the wards within the collection points were collated over a three -month period and defined daily dose (DDD) per 100 occupied bed days (OBD) calculated to represent the antimicrobial consumption at each collection point.

DDD= defined daily dose, OBD = occupied bed day, ITU=intensive care unit, SHDU = surgical high dependency unit.

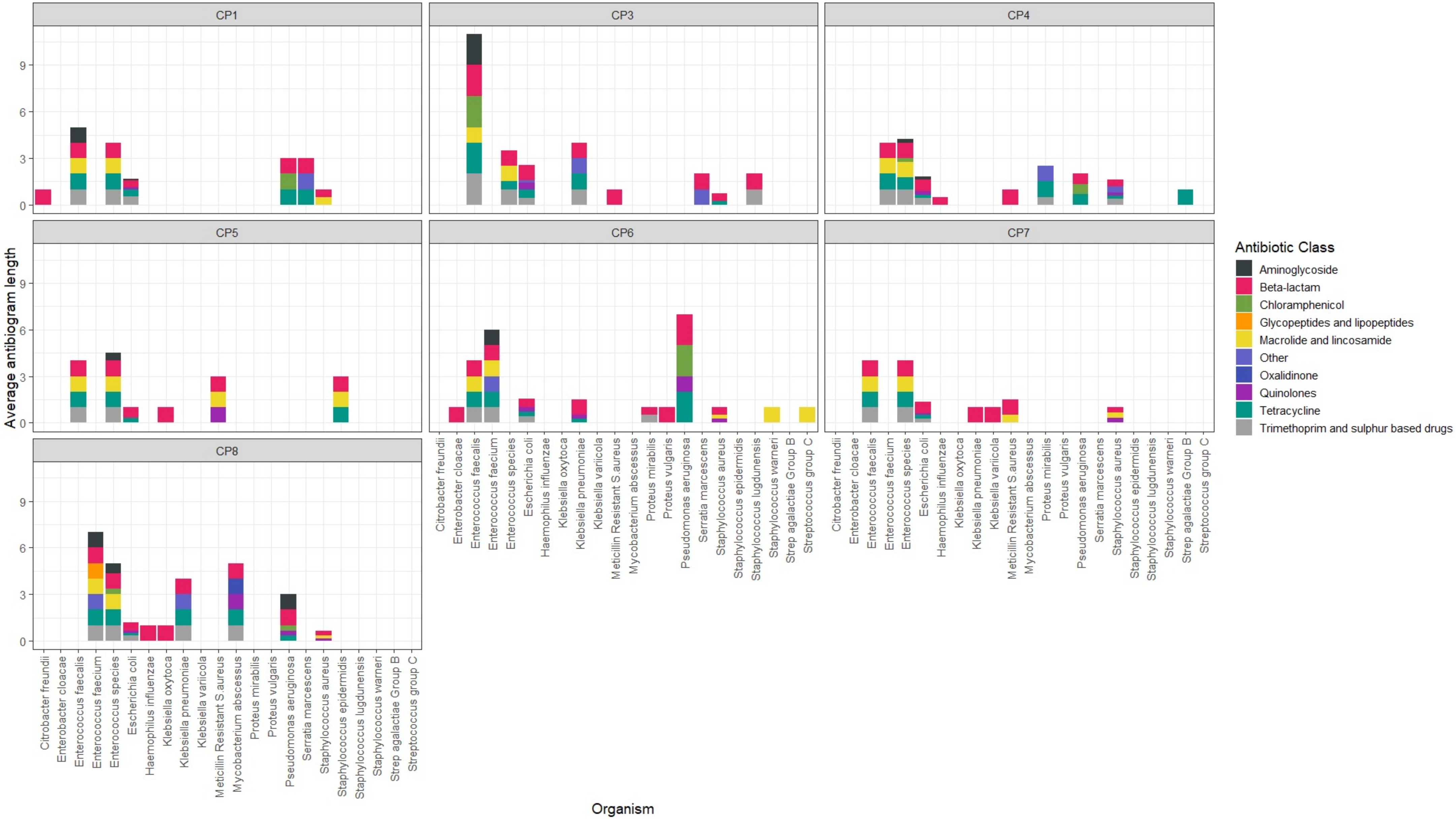


**Supplementary Figure 3** Phenotypic resistance in clinical isolates by antimicrobial class represented by average antibiogram length per organism at each collection point

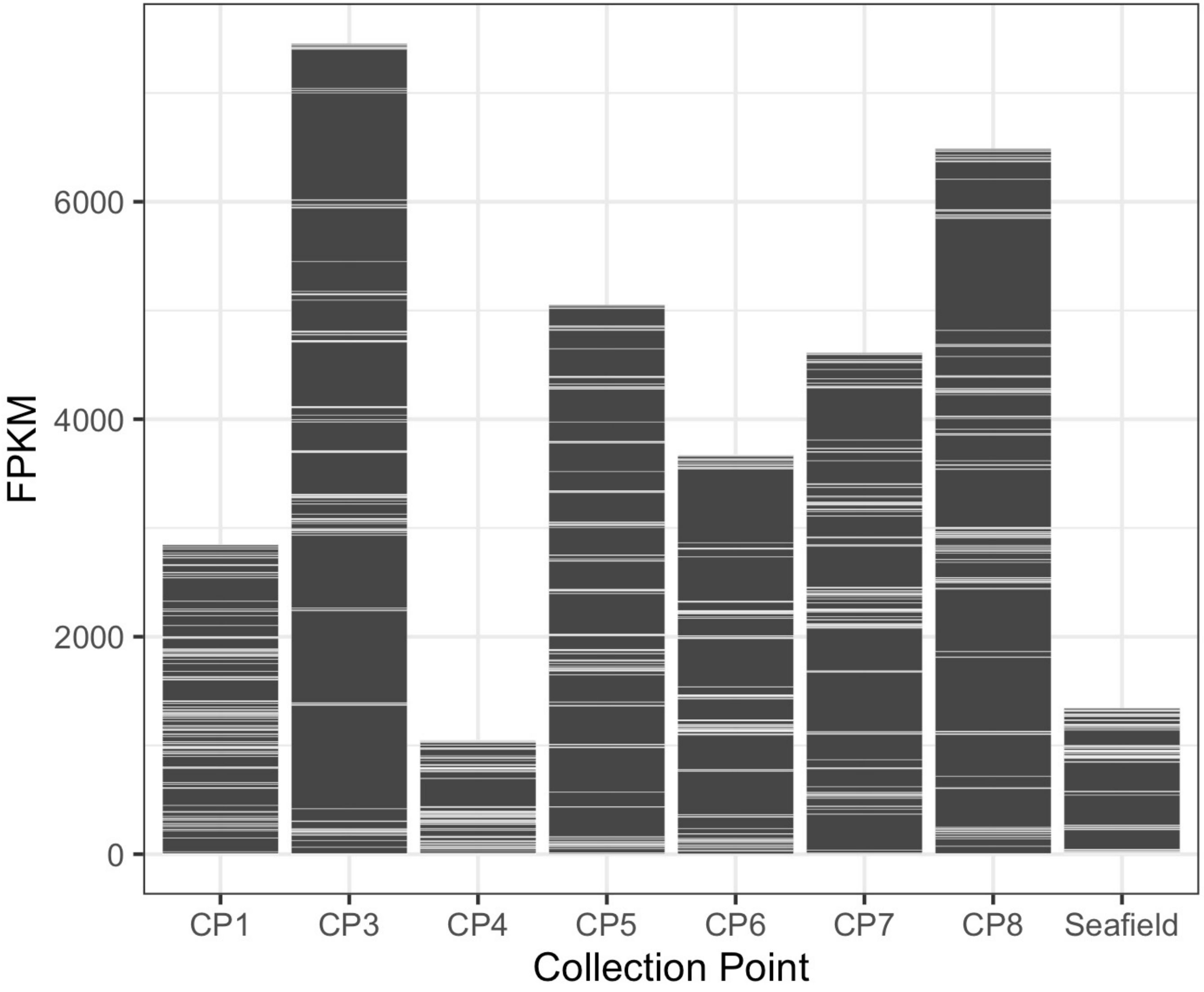


**Supplementary Figure 4** Total AMR gene abundance in FPKM

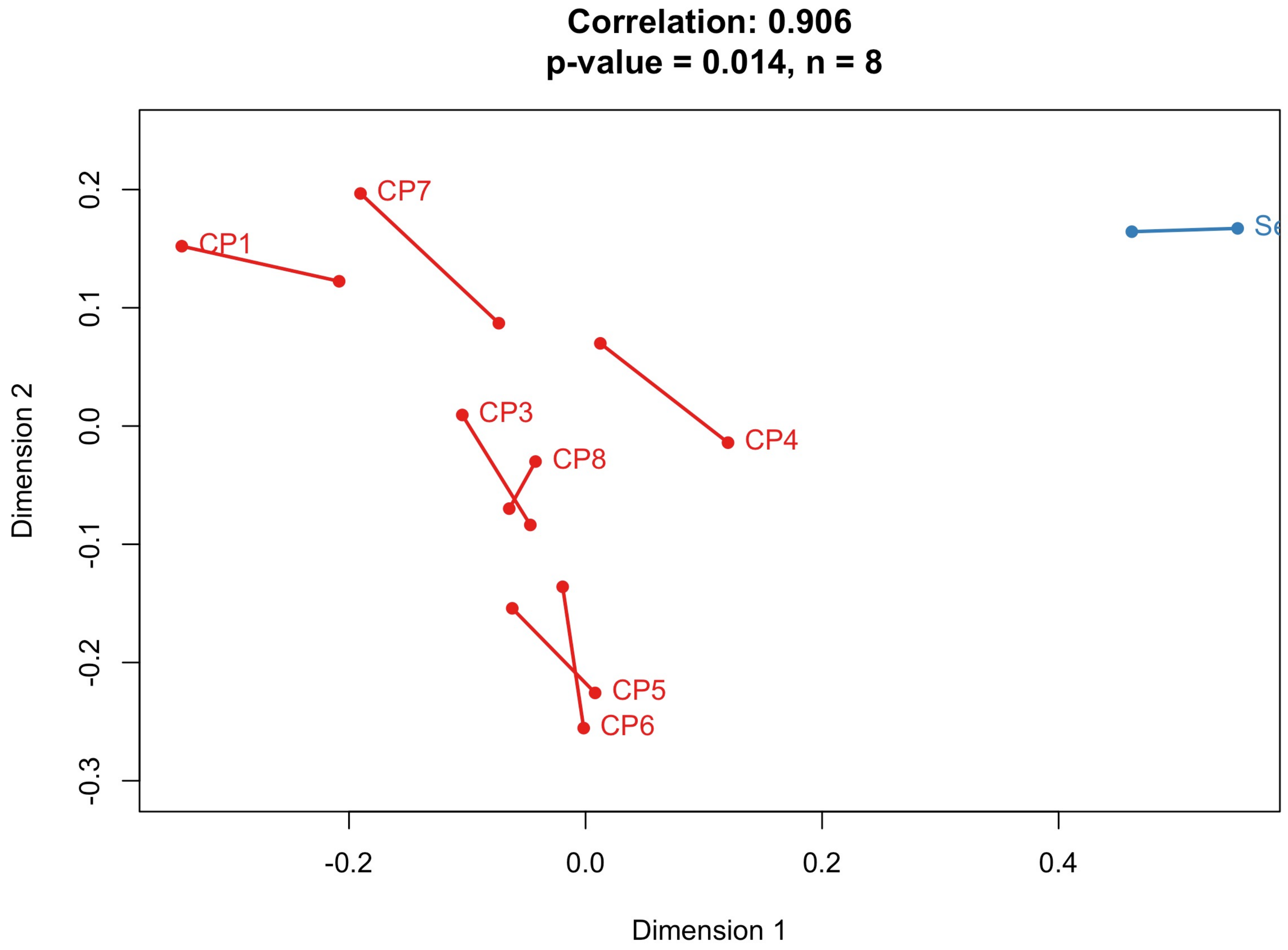


**Supplementary Figure 5** Procrustes analysis on ordinations of AMR genes and bacterial composition

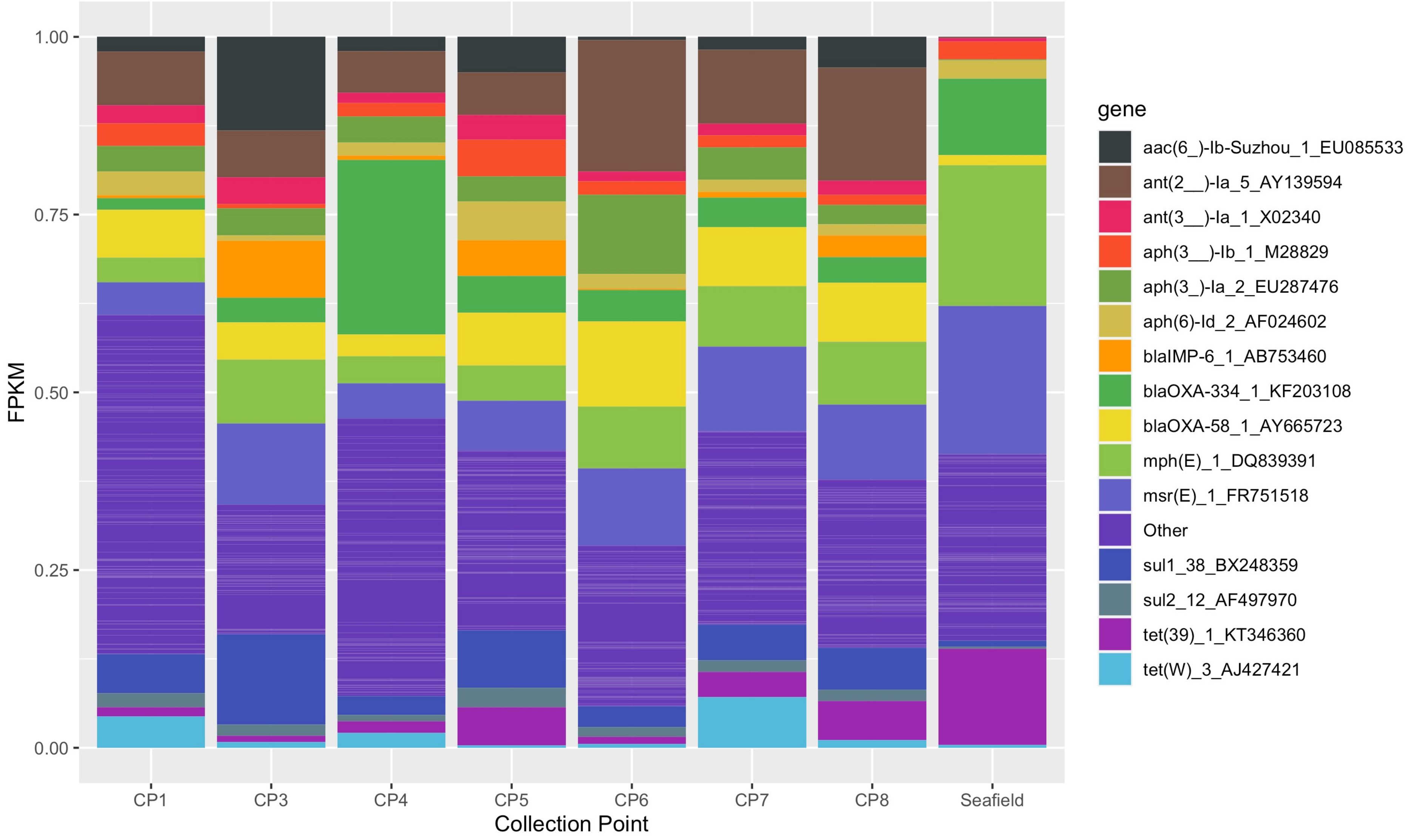


**Supplementary Figure 6** Fifteen most common AMR genes relative to total abundance within samples

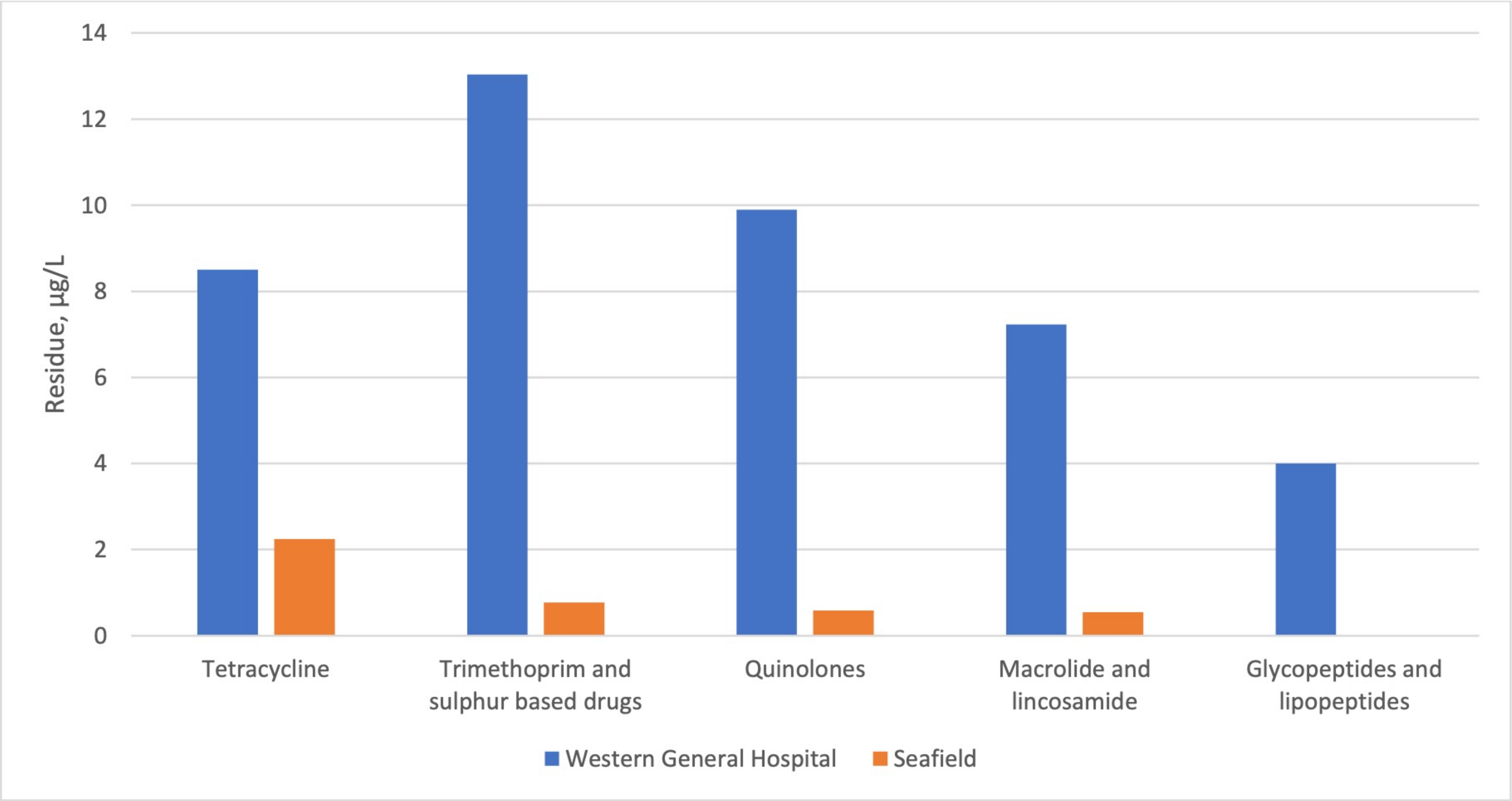


**Supplementary Figure 7** Antibiotic residues from composite hospital wastewater sample and urban sewage as measured by LC-MS/MS

Table S2

| Bacterial genus | CP1 | CP3 | CP4 | CP5 | CP6 | CP7 | CP8 | Seafield |
|---|---|---|---|---|---|---|---|---|
| Acinetobacter | 1604595 | 8858221 | 14545264 | 9253608 | 5301071 | 6084958 | 7617102 | 9367455 |
| Moraxella | 5012 | 8479 | 22845 | 8956 | 4858 | 8743 | 6683 | 751049 |
| Psychrobacter | 1252 | 5393 | 13810 | 2168 | 808 | 2426 | 2179 | 247366 |
| Pseudomonas | 1337571 | 2075434 | 3603155 | 1362168 | 7330230 | 1387450 | 2464891 | 8155080 |
| Azotobacter | 3588 | 4395 | 3655 | 5564 | 3015 | 2741 | 3052 | 4728 |
| Oblitimonas | 251 | 661 | 1513 | 339 | 133 | 257 | 484 | 1117 |
| Klebsiella | 854067 | 61633 | 42472 | 220763 | 62338 | 210958 | 324999 | 55315 |
| Escherichia | 626094 | 64854 | 49225 | 126536 | 1116220 | 210557 | 327398 | 26513 |
| Citrobacter | 27522 | 14909 | 40420 | 54589 | 13373 | 38541 | 58932 | 49004 |
| Enterobacter | 25852 | 13976 | 31045 | 30815 | 26297 | 53850 | 35869 | 95756 |
| Salmonella | 5921 | 2945 | 3674 | 3677 | 2605 | 4764 | 3806 | 8412 |
| Candidatus Blochmannia | 313 | 156 | 174 | 128 | 63 | 152 | 312 | 239 |
| Candidatus Hamiltonella | 158 | 136 | 214 | 109 | 86 | 149 | 206 | 843 |
| Candidatus Doolittlea | 90 | 27 | 13 | 10 | 9 | 26 | 25 | 35 |
| Candidatus Tachikawaea | 47 | 24 | 13 | 7 | 10 | 25 | 29 | 28 |
| Candidatus Hoaglandella | 39 | 45 | 19 | 18 | 12 | 26 | 35 | 36 |
| Candidatus Purcelliella | 33 | 27 | 23 | 32 | 4 | 22 | 15 | 56 |
| Candidatus Annandia | 32 | 15 | 1 | 5 | 1 | 12 | 6 | 15 |
| Candidatus Gullanella | 29 | 19 | 35 | 6 | 10 | 42 | 11 | 34 |
| Candidatus Mikella | 17 | 15 | 38 | 13 | 4 | 10 | 17 | 14 |
| Raoultella | 3701 | 7294 | 3595 | 7532 | 5081 | 11160 | 15986 | 21321 |
| Cronobacter | 2664 | 1892 | 1710 | 1864 | 1033 | 2194 | 2214 | 5896 |
| Shigella | 2149 | 218 | 131 | 299 | 1123 | 359 | 1077 | 325 |
| Kosakonia | 1948 | 1211 | 1999 | 1278 | 925 | 1741 | 1294 | 5056 |
| Pluralibacter | 1826 | 1032 | 1005 | 1264 | 632 | 1387 | 1128 | 2605 |
| Lelliottia | 1314 | 893 | 1444 | 2044 | 727 | 1416 | 1035 | 65515 |
| Cedecea | 1186 | 941 | 1520 | 912 | 538 | 1124 | 885 | 5535 |
| Metakosakonia | 793 | 115 | 281 | 427 | 223 | 660 | 447 | 206 |
| Leclercia | 694 | 736 | 2595 | 555 | 437 | 604 | 607 | 5465 |
| Gibbsiella | 467 | 313 | 590 | 513 | 240 | 394 | 363 | 818 |
| Kluyvera | 411 | 175 | 625 | 167 | 606 | 1107 | 266 | 36089 |
| Atlantibacter | 364 | 672 | 168 | 267 | 169 | 293 | 274 | 1206 |
| Shimwellia | 343 | 196 | 163 | 189 | 110 | 228 | 213 | 710 |
| Buttiauxella | 224 | 377 | 1407 | 353 | 132 | 249 | 224 | 41672 |
| Limnobaculum | 185 | 128 | 915 | 67 | 58 | 177 | 185 | 357 |
| Candidatus Ishikawaella | 69 | 28 | 16 | 20 | 4 | 32 | 25 | 113 |
| Candidatus Riesia | 49 | 51 | 14 | 18 | 17 | 80 | 48 | 43 |
| Candidatus Moranella | 47 | 31 | 5 | 22 | 11 | 19 | 22 | 17 |

| | | | | | | | | |
|---|---|---|---|---|---|---|---|---|
| **Serratia** | 12923 | 18865 | 8705 | 18952 | 5314 | 10265 | 9238 | 45081 |
| **Yersinia** | 5281 | 2627 | 7156 | 12727 | 2233 | 2806 | 2213 | 105850 |
| **Rahnella** | 1273 | 1174 | 938 | 1099 | 881 | 1033 | 1196 | 75871 |
| **Chania** | 278 | 215 | 171 | 253 | 119 | 286 | 231 | 629 |
| **Candidatus Fukatsuia** | 117 | 169 | 109 | 399 | 49 | 141 | 137 | 175 |
| **Proteus** | 7858 | 1200 | 3296 | 1661 | 11821 | 2600 | 1410 | 2789 |
| **Morganella** | 4465 | 3104 | 1067 | 2398 | 3100 | 2455 | 2519 | 1460 |
| **Providencia** | 927 | 1559 | 5254 | 863 | 500 | 1305 | 1399 | 3933 |
| **Xenorhabdus** | 717 | 728 | 2760 | 391 | 356 | 675 | 594 | 1978 |
| **Photorhabdus** | 360 | 403 | 1838 | 286 | 131 | 345 | 367 | 1133 |
| **Arsenophonus** | 124 | 165 | 167 | 27 | 45 | 81 | 65 | 230 |
| **Dickeya** | 4408 | 3710 | 3294 | 4274 | 1883 | 3607 | 3502 | 7807 |
| **Pectobacterium** | 1875 | 1692 | 2195 | 1728 | 742 | 1852 | 1603 | 6714 |
| **Brenneria** | 898 | 747 | 1141 | 668 | 341 | 832 | 677 | 1167 |
| **Sodalis** | 855 | 710 | 526 | 607 | 361 | 713 | 487 | 762 |
| **Lonsdalea** | 227 | 194 | 124 | 266 | 138 | 179 | 186 | 311 |
| **Pantoea** | 3335 | 2832 | 4232 | 2967 | 1553 | 3335 | 2551 | 8308 |
| **Erwinia** | 1604 | 1419 | 2176 | 1485 | 777 | 1395 | 1401 | 5097 |
| **Buchnera** | 459 | 416 | 135 | 202 | 120 | 280 | 273 | 420 |
| **Mixta** | 365 | 306 | 233 | 322 | 173 | 272 | 237 | 422 |
| **Tatumella** | 334 | 322 | 199 | 211 | 132 | 325 | 227 | 638 |
| **Wigglesworthia** | 51 | 44 | 26 | 22 | 11 | 39 | 36 | 137 |
| **Plesiomonas** | 2831 | 542 | 273 | 726 | 172 | 1557 | 761 | 1350 |
| **Phytobacter** | 1833 | 953 | 2854 | 1000 | 2232 | 3887 | 1244 | 648 |
| **Edwardsiella** | 1705 | 1265 | 1197 | 1227 | 672 | 1609 | 1110 | 1526 |
| **Hafnia** | 451 | 625 | 1257 | 2323 | 762 | 1064 | 1375 | 6343 |
| **Obesumbacterium** | 117 | 80 | 53 | 111 | 865 | 334 | 124 | 1872 |
| **Leminorella** | 337 | 195 | 171 | 164 | 94 | 338 | 150 | 318 |
| **Pragia** | 295 | 347 | 723 | 145 | 160 | 401 | 252 | 1010 |
| **Stenotrophomonas** | 129607 | 272935 | 232378 | 137529 | 46509 | 37691 | 55058 | 58882 |
| **Xanthomonas** | 66411 | 49720 | 39052 | 65787 | 20620 | 22048 | 33576 | 24331 |
| **Lysobacter** | 65277 | 36462 | 24246 | 49947 | 12289 | 13463 | 25705 | 18990 |
| **Pseudoxanthomonas** | 48212 | 23750 | 15725 | 43240 | 14476 | 9043 | 14187 | 8727 |
| **Luteimonas** | 36083 | 17673 | 12756 | 25889 | 5827 | 6881 | 10198 | 8315 |
| **Xylella** | 1063 | 716 | 504 | 949 | 277 | 528 | 634 | 724 |
| **Dyella** | 9855 | 7904 | 3067 | 6179 | 2361 | 3569 | 5141 | 2283 |
| **Dokdonella** | 8473 | 6873 | 1991 | 4290 | 1666 | 2163 | 5308 | 1386 |
| **Rhodanobacter** | 5853 | 4424 | 1936 | 4384 | 1213 | 2151 | 3149 | 1353 |
| **Luteibacter** | 2311 | 1914 | 701 | 1413 | 642 | 1041 | 1278 | 581 |
| **Ahniella** | 1070 | 837 | 494 | 751 | 355 | 450 | 631 | 464 |

| | | | | | | | | |
|---|---|---|---|---|---|---|---|---|
| **Frateuria** | 918 | 776 | 822 | 743 | 303 | 503 | 616 | 455 |
| **Haemophilus** | 49097 | 12316 | 3163 | 3159 | 3787 | 17155 | 10240 | 5388 |
| **Aggregatibacter** | 973 | 744 | 525 | 302 | 229 | 684 | 615 | 1069 |
| **Actinobacillus** | 892 | 716 | 982 | 416 | 220 | 777 | 596 | 1438 |
| **Pasteurella** | 623 | 1649 | 1553 | 565 | 220 | 555 | 1077 | 1769 |
| **Mannheimia** | 522 | 461 | 1258 | 373 | 156 | 471 | 404 | 1070 |
| **Glaesserella** | 363 | 287 | 570 | 120 | 123 | 282 | 285 | 739 |
| **Avibacterium** | 197 | 171 | 1285 | 103 | 69 | 151 | 332 | 445 |
| **Histophilus** | 191 | 141 | 80 | 88 | 48 | 143 | 219 | 315 |
| **Bibersteinia** | 167 | 184 | 344 | 53 | 34 | 176 | 124 | 314 |
| **Basfia** | 118 | 103 | 538 | 57 | 33 | 150 | 158 | 224 |
| **Gallibacterium** | 116 | 172 | 152 | 70 | 58 | 153 | 129 | 324 |
| **Aeromonas** | 46560 | 29421 | 45972 | 52462 | 52976 | 189850 | 169742 | 613987 |
| **Tolumonas** | 1346 | 1079 | 417 | 641 | 290 | 1022 | 832 | 10857 |
| **Zobellella** | 1154 | 853 | 703 | 921 | 512 | 828 | 706 | 691 |
| **Oceanimonas** | 631 | 501 | 519 | 441 | 271 | 512 | 407 | 592 |
| **Oceanisphaera** | 331 | 311 | 1958 | 477 | 128 | 411 | 253 | 984 |
| **Thioalkalivibrio** | 4654 | 3009 | 2298 | 3284 | 1563 | 2496 | 2569 | 1652 |
| **Acidihalobacter** | 2325 | 1818 | 1420 | 2218 | 795 | 1297 | 1469 | 1305 |
| **Ectothiorhodospira** | 1105 | 877 | 645 | 853 | 354 | 829 | 705 | 635 |
| **Halorhodospira** | 911 | 582 | 526 | 612 | 341 | 665 | 470 | 383 |
| **Alkalilimnicola** | 819 | 604 | 536 | 725 | 303 | 484 | 485 | 311 |
| **Spiribacter** | 466 | 338 | 276 | 442 | 186 | 421 | 329 | 286 |
| **Thiodictyon** | 1590 | 1288 | 805 | 1414 | 598 | 964 | 829 | 749 |
| **Marichromatium** | 1131 | 749 | 581 | 944 | 464 | 644 | 590 | 377 |
| **Allochromatium** | 1047 | 683 | 520 | 720 | 366 | 516 | 468 | 364 |
| **Thioflavicoccus** | 984 | 749 | 520 | 632 | 349 | 609 | 556 | 414 |
| **Thiocystis** | 875 | 792 | 520 | 745 | 383 | 547 | 550 | 480 |
| **Nitrosococcus** | 423 | 378 | 283 | 282 | 152 | 381 | 278 | 476 |
| **Rheinheimera** | 190 | 322 | 463 | 1077 | 146 | 301 | 174 | 543 |
| **Candidatus Nitrosoglobus** | 156 | 74 | 391 | 135 | 28 | 88 | 92 | 256 |
| **Sulfurivermis** | 1956 | 1047 | 746 | 1748 | 603 | 1020 | 993 | 651 |
| **Halothiobacillus** | 846 | 765 | 503 | 800 | 422 | 724 | 632 | 672 |
| **Wenzhouxiangella** | 800 | 617 | 359 | 742 | 295 | 478 | 479 | 294 |
| **Sulfuriflexus** | 334 | 274 | 261 | 193 | 101 | 287 | 292 | 221 |
| **Granulosicoccus** | 319 | 221 | 230 | 265 | 124 | 282 | 229 | 293 |
| **Woeseia** | 313 | 254 | 150 | 191 | 114 | 233 | 239 | 184 |
| **Halomonas** | 9330 | 8146 | 8044 | 8597 | 3970 | 7061 | 7016 | 7430 |
| **Halotalea** | 955 | 820 | 575 | 997 | 458 | 588 | 591 | 512 |
| **Salinicola** | 833 | 684 | 505 | 809 | 371 | 630 | 599 | 406 |

| | | | | | | | | |
|---|---|---|---|---|---|---|---|---|
| **Kushneria** | 804 | 547 | 423 | 591 | 311 | 649 | 502 | 492 |
| **Chromohalobacter** | 652 | 516 | 406 | 653 | 292 | 369 | 372 | 357 |
| **Cobetia** | 436 | 378 | 235 | 381 | 176 | 318 | 283 | 328 |
| **Zymobacter** | 235 | 204 | 419 | 192 | 79 | 228 | 139 | 153 |
| **Candidatus Carsonella** | 22 | 31 | 15 | 14 | 14 | 19 | 17 | 28 |
| **Candidatus Evansia** | 22 | 14 | 18 | 6 | 4 | 21 | 20 | 15 |
| **Candidatus Portiera** | 8 | 9 | 15 | 14 | 2 | 11 | 8 | 25 |
| **Alcanivorax** | 3246 | 2586 | 2890 | 3040 | 1645 | 2363 | 2488 | 2551 |
| **Ketobacter** | 247 | 247 | 166 | 171 | 102 | 274 | 153 | 309 |
| **Marinobacterium** | 597 | 542 | 374 | 716 | 343 | 503 | 500 | 557 |
| **Marinomonas** | 562 | 609 | 1238 | 453 | 213 | 656 | 732 | 1628 |
| **Thalassolituus** | 460 | 541 | 1805 | 570 | 138 | 347 | 510 | 1203 |
| **Bacterioplanes** | 181 | 163 | 137 | 112 | 68 | 180 | 109 | 413 |
| **Oleispira** | 131 | 386 | 322 | 998 | 52 | 280 | 192 | 963 |
| **Saccharospirillum** | 377 | 291 | 230 | 305 | 185 | 357 | 298 | 307 |
| **Gynuella** | 266 | 266 | 822 | 187 | 103 | 263 | 171 | 404 |
| **Reinekea** | 187 | 177 | 123 | 201 | 91 | 157 | 155 | 223 |
| **Hahella** | 650 | 606 | 726 | 598 | 328 | 605 | 486 | 708 |
| **Kangiella** | 330 | 360 | 1822 | 240 | 123 | 262 | 360 | 981 |
| **Endozoicomonas** | 223 | 263 | 389 | 132 | 61 | 278 | 150 | 347 |
| **Oleiphilus** | 146 | 201 | 127 | 96 | 55 | 162 | 86 | 263 |
| **Marinobacter** | 4332 | 3900 | 3863 | 4148 | 1849 | 3791 | 3502 | 4416 |
| **Alteromonas** | 1407 | 1627 | 3882 | 1217 | 469 | 1361 | 1341 | 3069 |
| **Glaciecola** | 741 | 852 | 849 | 911 | 279 | 585 | 990 | 744 |
| **Salinimonas** | 322 | 263 | 174 | 211 | 101 | 296 | 187 | 542 |
| **Lacimicrobium** | 240 | 210 | 440 | 158 | 126 | 273 | 159 | 327 |
| **Catenovulum** | 155 | 204 | 872 | 137 | 57 | 152 | 127 | 309 |
| **Paraglaciecola** | 122 | 153 | 162 | 103 | 39 | 138 | 128 | 428 |
| **Agarivorans** | 117 | 76 | 139 | 95 | 41 | 84 | 133 | 246 |
| **Shewanella** | 5072 | 5360 | 11262 | 6674 | 3964 | 4975 | 4756 | 817172 |
| **Pseudoalteromonas** | 2321 | 4381 | 8330 | 2527 | 980 | 2795 | 3160 | 8340 |
| **Colwellia** | 739 | 1443 | 3822 | 647 | 312 | 755 | 1376 | 2537 |
| **Thalassotalea** | 134 | 117 | 67 | 58 | 21 | 111 | 71 | 487 |
| **Litorilituus** | 63 | 114 | 661 | 51 | 30 | 79 | 154 | 351 |
| **Idiomarina** | 437 | 464 | 845 | 277 | 165 | 386 | 484 | 954 |
| **Ferrimonas** | 488 | 358 | 231 | 402 | 193 | 331 | 309 | 322 |
| **Moritella** | 268 | 299 | 2237 | 309 | 111 | 264 | 253 | 883 |
| **Psychromonas** | 255 | 328 | 715 | 340 | 78 | 218 | 212 | 676 |
| **Vibrio** | 7078 | 7250 | 16650 | 6862 | 3018 | 7364 | 6597 | 23410 |
| **Photobacterium** | 652 | 702 | 1337 | 431 | 518 | 748 | 521 | 1226 |

| | | | | | | | | |
|---|---|---|---|---|---|---|---|---|
| **Aliivibrio** | 482 | 433 | 1843 | 435 | 325 | 505 | 615 | 1180 |
| **Grimontia** | 190 | 146 | 416 | 91 | 58 | 212 | 113 | 208 |
| **Paraphotobacterium** | 92 | 47 | 29 | 41 | 33 | 120 | 85 | 137 |
| **Enterovibrio** | 45 | 27 | 32 | 35 | 13 | 35 | 27 | 87 |
| **Methylomonas** | 1959 | 2020 | 1131 | 1705 | 921 | 1579 | 1406 | 2093 |
| **Methylococcus** | 1455 | 727 | 529 | 979 | 540 | 748 | 743 | 367 |
| **Methylomicrobium** | 1282 | 944 | 701 | 1175 | 440 | 884 | 778 | 913 |
| **Methylocaldum** | 908 | 765 | 452 | 853 | 395 | 547 | 532 | 579 |
| **Methylovulum** | 374 | 3552 | 614 | 1831 | 554 | 1064 | 1194 | 2718 |
| **Thiohalobacter** | 1020 | 866 | 574 | 832 | 365 | 688 | 667 | 516 |
| **Sedimenticola** | 496 | 298 | 246 | 327 | 155 | 367 | 268 | 350 |
| **Candidatus Ruthia** | 57 | 47 | 79 | 16 | 19 | 52 | 34 | 63 |
| **Thiolapillus** | 305 | 201 | 182 | 213 | 90 | 286 | 193 | 207 |
| **Gallaecimonas** | 239 | 243 | 164 | 174 | 64 | 271 | 181 | 354 |
| **Candidatus Thioglobus** | 235 | 221 | 607 | 79 | 64 | 131 | 329 | 434 |
| **Pseudohongiella** | 206 | 173 | 155 | 101 | 75 | 189 | 110 | 271 |
| **Candidatus Nardonella** | 42 | 48 | 11 | 17 | 5 | 48 | 29 | 42 |
| **Cellvibrio** | 1255 | 3710 | 1870 | 1342 | 549 | 919 | 982 | 1425 |
| **Simiduia** | 334 | 344 | 314 | 333 | 135 | 290 | 243 | 470 |
| **Teredinibacter** | 187 | 172 | 110 | 129 | 54 | 174 | 119 | 286 |
| **Saccharophagus** | 128 | 179 | 346 | 102 | 38 | 84 | 71 | 226 |
| **Agarilytica** | 123 | 122 | 595 | 59 | 51 | 107 | 185 | 231 |
| **Microbulbifer** | 1203 | 1007 | 1824 | 871 | 434 | 1038 | 702 | 916 |
| **Halioglobus** | 745 | 539 | 372 | 569 | 263 | 554 | 413 | 554 |
| **Congregibacter** | 290 | 290 | 277 | 226 | 155 | 348 | 239 | 258 |
| **Zhongshania** | 206 | 235 | 165 | 567 | 106 | 169 | 146 | 466 |
| **Oceanicoccus** | 154 | 156 | 447 | 84 | 52 | 124 | 103 | 210 |
| **Spongiibacter** | 113 | 155 | 240 | 111 | 44 | 83 | 106 | 329 |
| **Solimonas** | 1973 | 1584 | 1133 | 1683 | 878 | 1071 | 1302 | 757 |
| **Steroidobacter** | 1323 | 1151 | 865 | 1338 | 366 | 615 | 763 | 413 |
| **Methylophaga** | 375 | 659 | 1200 | 435 | 140 | 358 | 327 | 1221 |
| **Thiomicrospira** | 350 | 317 | 1141 | 210 | 109 | 309 | 581 | 782 |
| **Thiomicrorhabdus** | 242 | 285 | 274 | 126 | 59 | 276 | 173 | 443 |
| **Cycloclasticus** | 153 | 171 | 125 | 190 | 56 | 213 | 165 | 366 |
| **Hydrogenovibrio** | 143 | 170 | 185 | 166 | 55 | 159 | 164 | 281 |
| **Piscirickettsia** | 111 | 188 | 851 | 59 | 30 | 85 | 83 | 232 |
| **Francisella** | 1083 | 921 | 1319 | 741 | 409 | 1041 | 1162 | 2049 |
| **Allofrancisella** | 83 | 38 | 10 | 17 | 36 | 44 | 29 | 56 |
| **Thioploca** | 144 | 152 | 94 | 51 | 54 | 107 | 91 | 262 |
| **Beggiatoa** | 141 | 251 | 145 | 83 | 49 | 100 | 178 | 794 |

| | | | | | | | | |
|---|---|---|---|---|---|---|---|---|
| **Legionella** | 2004 | 2358 | 3480 | 1430 | 1037 | 1903 | 1674 | 2795 |
| **Tatlockia** | 126 | 142 | 54 | 150 | 263 | 124 | 108 | 135 |
| **Fluoribacter** | 81 | 51 | 33 | 40 | 24 | 91 | 60 | 119 |
| **Coxiella** | 187 | 226 | 264 | 158 | 71 | 170 | 132 | 258 |
| **Rickettsiella** | 44 | 73 | 562 | 49 | 33 | 59 | 45 | 124 |
| **Sulfurifustis** | 1170 | 871 | 615 | 838 | 431 | 634 | 626 | 324 |
| **Sulfuricaulis** | 840 | 647 | 367 | 677 | 351 | 525 | 448 | 430 |
| **Acidiferrobacter** | 444 | 336 | 178 | 274 | 176 | 276 | 250 | 180 |
| **Immundisolibacter** | 1537 | 1131 | 922 | 1373 | 558 | 918 | 947 | 604 |
| **Salinisphaera** | 722 | 726 | 414 | 627 | 373 | 455 | 525 | 301 |
| **Cardiobacterium** | 511 | 363 | 337 | 375 | 200 | 429 | 317 | 369 |
| **Dichelobacter** | 151 | 153 | 554 | 56 | 56 | 135 | 103 | 395 |
| **Gilliamella** | 166 | 230 | 107 | 84 | 55 | 207 | 222 | 433 |
| **Frischella** | 98 | 69 | 46 | 34 | 38 | 64 | 109 | 150 |
| **Acidovorax** | 781425 | 1369344 | 758282 | 1990867 | 326274 | 653834 | 1014081 | 1335766 |
| **Comamonas** | 369001 | 91937 | 393272 | 153795 | 44170 | 82376 | 282577 | 364347 |
| **Delftia** | 125105 | 56636 | 63662 | 254659 | 56084 | 89227 | 172752 | 64968 |
| **Variovorax** | 101855 | 139067 | 60486 | 191064 | 63874 | 53079 | 135785 | 66088 |
| **Ottowia** | 89737 | 39380 | 28935 | 427908 | 18443 | 40606 | 82365 | 43083 |
| **Alicycliphilus** | 72772 | 74776 | 108903 | 96916 | 25107 | 25766 | 50996 | 31937 |
| **Hydrogenophaga** | 47746 | 51325 | 46694 | 85918 | 24161 | 28232 | 62556 | 42989 |
| **Melaminivora** | 46265 | 47921 | 62281 | 73121 | 18549 | 21990 | 46336 | 35583 |
| **Rhodoferax** | 35159 | 50962 | 28713 | 96087 | 25980 | 25708 | 70217 | 51570 |
| **Polaromonas** | 28206 | 39704 | 23669 | 70164 | 19613 | 19650 | 52955 | 45421 |
| **Ramlibacter** | 22453 | 25050 | 17008 | 36844 | 12243 | 11749 | 28907 | 14923 |
| **Simplicispira** | 14768 | 17877 | 14635 | 32410 | 6927 | 10731 | 21101 | 27093 |
| **Verminephrobacter** | 13542 | 13904 | 12296 | 26675 | 5710 | 9183 | 19784 | 16459 |
| **Serpentinomonas** | 11585 | 9354 | 9652 | 17361 | 5451 | 6499 | 11469 | 11033 |
| **Limnohabitans** | 9520 | 11031 | 8650 | 23975 | 5083 | 7533 | 16447 | 15870 |
| **Curvibacter** | 6585 | 8347 | 6429 | 16253 | 3667 | 4901 | 10954 | 10385 |
| **Diaphorobacter** | 5666 | 5806 | 13122 | 14256 | 2453 | 3307 | 5872 | 4802 |
| **Candidatus Symbiobacter** | 1665 | 1232 | 1191 | 2100 | 530 | 1082 | 1602 | 1832 |
| **Burkholderia** | 86269 | 68978 | 41345 | 84613 | 42003 | 53193 | 60221 | 34190 |
| **Cupriavidus** | 74739 | 62019 | 43885 | 64067 | 59721 | 72060 | 60302 | 29694 |
| **Ralstonia** | 27680 | 22537 | 18240 | 31573 | 11900 | 14772 | 24697 | 14416 |
| **Paraburkholderia** | 18625 | 18710 | 11564 | 21047 | 11389 | 12114 | 15724 | 11813 |
| **Pandoraea** | 17646 | 17268 | 10561 | 17358 | 7997 | 9628 | 14802 | 10213 |
| **Lautropia** | 1893 | 1507 | 1153 | 2128 | 2765 | 1385 | 2437 | 1041 |
| **Polynucleobacter** | 1856 | 2107 | 3806 | 1826 | 644 | 1708 | 1931 | 4809 |
| **Hydromonas** | 382 | 236 | 240 | 351 | 85 | 208 | 294 | 41656 |

| | | | | | | | | |
|---|---|---|---|---|---|---|---|---|
| **Mycoavidus** | 182 | 101 | 131 | 197 | 44 | 102 | 177 | 279 |
| **Methylibium** | 12238 | 9457 | 6402 | 10953 | 6521 | 4836 | 8626 | 3811 |
| **Thiomonas** | 10829 | 8475 | 6966 | 14336 | 4045 | 7592 | 9513 | 10374 |
| **Rubrivivax** | 9960 | 7764 | 6608 | 8618 | 6275 | 4087 | 6543 | 2680 |
| **Rhizobacter** | 8551 | 7061 | 4261 | 8484 | 4894 | 3733 | 6753 | 3338 |
| **Leptothrix** | 8375 | 7499 | 5134 | 9210 | 4965 | 3902 | 7113 | 4043 |
| **Aquabacterium** | 6961 | 5728 | 3541 | 8070 | 4700 | 3179 | 4826 | 3619 |
| **Mitsuaria** | 6388 | 4955 | 2968 | 5577 | 3202 | 2490 | 4275 | 1913 |
| **Paucibacter** | 4447 | 5416 | 3154 | 5916 | 2127 | 2856 | 5399 | 4152 |
| **Roseateles** | 4059 | 3801 | 2389 | 5367 | 2231 | 2019 | 3709 | 2361 |
| **Achromobacter** | 33403 | 30773 | 22674 | 40314 | 17138 | 17419 | 29834 | 22627 |
| **Bordetella** | 31927 | 29465 | 24133 | 41870 | 15775 | 19656 | 28811 | 20678 |
| **Pigmentiphaga** | 3291 | 2899 | 2447 | 3536 | 1544 | 1597 | 2561 | 1540 |
| **Alcaligenes** | 3125 | 3647 | 3782 | 9841 | 1441 | 2538 | 3439 | 5709 |
| **Castellaniella** | 2898 | 2095 | 1958 | 2896 | 1113 | 1275 | 2036 | 1191 |
| **Orrella** | 2370 | 2103 | 1564 | 2974 | 1203 | 1464 | 2228 | 1393 |
| **Pusillimonas** | 1357 | 1094 | 876 | 1625 | 568 | 841 | 1215 | 1465 |
| **Advenella** | 949 | 846 | 1403 | 1059 | 433 | 645 | 894 | 1496 |
| **Oligella** | 336 | 142 | 645 | 124 | 88 | 129 | 142 | 433 |
| **Taylorella** | 192 | 165 | 161 | 78 | 43 | 202 | 105 | 288 |
| **Paenalcaligenes** | 122 | 94 | 73 | 112 | 30 | 75 | 84 | 446 |
| **Basilea** | 117 | 98 | 254 | 45 | 59 | 131 | 103 | 425 |
| **Massilia** | 25884 | 138986 | 30106 | 281865 | 53527 | 27453 | 121951 | 17248 |
| **Janthinobacterium** | 17401 | 49221 | 35649 | 1237082 | 159598 | 23175 | 45713 | 429464 |
| **Herbaspirillum** | 9875 | 11723 | 7114 | 18094 | 5861 | 5898 | 11445 | 7469 |
| **Collimonas** | 5441 | 6990 | 4823 | 11269 | 3538 | 3766 | 7321 | 5883 |
| **Oxalobacter** | 1228 | 761 | 274 | 243 | 155 | 687 | 581 | 383 |
| **Herminiimonas** | 1099 | 1268 | 767 | 1976 | 639 | 751 | 1171 | 1542 |
| **Undibacterium** | 608 | 547 | 417 | 891 | 253 | 377 | 652 | 969 |
| **Sutterella** | 1538 | 2256 | 569 | 559 | 415 | 2057 | 688 | 297 |
| **Thauera** | 42137 | 22057 | 47484 | 33662 | 9912 | 12682 | 16811 | 18759 |
| **Azoarcus** | 29849 | 17111 | 14502 | 21463 | 8760 | 11544 | 14085 | 12237 |
| **Dechloromonas** | 29222 | 13267 | 2884 | 7081 | 7560 | 4746 | 5052 | 9515 |
| **Azospira** | 12307 | 5084 | 2629 | 5880 | 7629 | 5475 | 4428 | 2457 |
| **Aromatoleum** | 3524 | 1964 | 1914 | 2613 | 1044 | 1428 | 1554 | 1485 |
| **Aquaspirillum** | 11046 | 3379 | 1552 | 6540 | 9857 | 5467 | 6044 | 3984 |
| **Chromobacterium** | 8288 | 6231 | 4580 | 7663 | 4071 | 4945 | 5606 | 4707 |
| **Iodobacter** | 216 | 160 | 747 | 123 | 110 | 128 | 134 | 471 |
| **Laribacter** | 3138 | 1690 | 1239 | 3134 | 885 | 1924 | 2639 | 1836 |
| **Jeongeupia** | 2315 | 1377 | 1008 | 1863 | 788 | 1015 | 1259 | 865 |

| | | | | | | | | |
|---|---|---|---|---|---|---|---|---|
| **Aquitalea** | 2250 | 1395 | 810 | 1376 | 791 | 965 | 1337 | 1428 |
| **Pseudogulbenkiania** | 1876 | 1207 | 828 | 1536 | 802 | 909 | 1217 | 930 |
| **Vogesella** | 1772 | 1302 | 947 | 1728 | 803 | 1061 | 1305 | 1189 |
| **Microvirgula** | 1650 | 6038 | 743 | 2344 | 906 | 8861 | 1670 | 1802 |
| **Neisseria** | 6275 | 4674 | 3852 | 3839 | 2486 | 4828 | 5731 | 48488 |
| **Vitreoscilla** | 3527 | 2333 | 2160 | 3681 | 1243 | 2165 | 3334 | 14739 |
| **Crenobacter** | 1560 | 1017 | 672 | 1135 | 621 | 674 | 829 | 600 |
| **Eikenella** | 848 | 465 | 736 | 532 | 229 | 731 | 427 | 4118 |
| **Kingella** | 203 | 113 | 77 | 96 | 46 | 177 | 98 | 1385 |
| **Snodgrassella** | 187 | 135 | 94 | 133 | 63 | 169 | 205 | 559 |
| **Simonsiella** | 97 | 144 | 67 | 55 | 23 | 72 | 103 | 721 |
| **Sulfuritalea** | 3108 | 2323 | 1215 | 1708 | 1092 | 1078 | 1236 | 1615 |
| **Methyloversatilis** | 2597 | 1666 | 1109 | 1846 | 959 | 1027 | 1286 | 1130 |
| **Nitrosomonas** | 3309 | 2289 | 3318 | 2355 | 997 | 2164 | 2837 | 2159 |
| **Nitrosospira** | 1659 | 1061 | 1149 | 1277 | 418 | 1524 | 2772 | 1007 |
| **Sulfuritortus** | 3346 | 1708 | 1141 | 2736 | 827 | 1473 | 1572 | 743 |
| **Thiobacillus** | 1545 | 1423 | 789 | 1117 | 550 | 719 | 800 | 582 |
| **Sideroxydans** | 967 | 671 | 399 | 664 | 360 | 525 | 523 | 529 |
| **Sulfuriferula** | 964 | 638 | 402 | 1042 | 285 | 905 | 1895 | 1553 |
| **Sulfuricella** | 765 | 559 | 307 | 514 | 259 | 456 | 411 | 530 |
| **Gallionella** | 685 | 469 | 406 | 738 | 216 | 479 | 470 | 683 |
| **Ferriphaselus** | 612 | 446 | 298 | 475 | 265 | 385 | 334 | 518 |
| **Methylovorus** | 799 | 563 | 504 | 791 | 281 | 539 | 582 | 811 |
| **Methylobacillus** | 710 | 605 | 367 | 678 | 252 | 492 | 493 | 475 |
| **Methylotenera** | 253 | 252 | 752 | 171 | 86 | 252 | 329 | 1021 |
| **Candidatus Methylopumilu** | 221 | 161 | 254 | 131 | 116 | 198 | 160 | 617 |
| **Methylophilus** | 205 | 176 | 120 | 145 | 71 | 137 | 121 | 330 |
| **Candidatus Accumulibact** | 4572 | 2693 | 1343 | 2811 | 1597 | 1747 | 1990 | 2740 |
| **Candidatus Kinetoplastiba** | 250 | 171 | 363 | 104 | 143 | 233 | 219 | 272 |
| **Candidatus Profftella** | 35 | 34 | 5 | 12 | 5 | 34 | 13 | 32 |
| **Candidatus Tremblaya** | 21 | 1 | 1 | 0 | 0 | 2 | 2 | 3 |
| **Candidatus Nasuia** | 6 | 5 | 0 | 1 | 0 | 1 | 1 | 9 |
| **Sphingopyxis** | 449715 | 478936 | 241500 | 311381 | 201253 | 261738 | 276576 | 52743 |
| **Sphingobium** | 182771 | 127888 | 44622 | 66615 | 53582 | 74606 | 66456 | 14898 |
| **Sphingomonas** | 135456 | 140573 | 71474 | 93070 | 75580 | 69807 | 77493 | 15133 |
| **Novosphingobium** | 45116 | 37791 | 27249 | 36756 | 22324 | 23940 | 21569 | 7602 |
| **Blastomonas** | 26400 | 10666 | 6427 | 8594 | 17658 | 9709 | 14404 | 2262 |
| **Sphingosinicella** | 6445 | 6593 | 3260 | 4921 | 3417 | 3532 | 3785 | 1213 |
| **Sphingorhabdus** | 5816 | 4082 | 2843 | 3217 | 2415 | 3004 | 3894 | 1160 |
| **Rhizorhabdus** | 5455 | 5275 | 3169 | 3755 | 2909 | 3007 | 3190 | 756 |

| | | | | | | | | |
|---|---|---|---|---|---|---|---|---|
| **Citromicrobium** | 2151 | 1893 | 1340 | 1332 | 1048 | 1162 | 1272 | 425 |
| **Zymomonas** | 598 | 619 | 791 | 1531 | 276 | 585 | 556 | 314 |
| **Altererythrobacter** | 15848 | 14047 | 11553 | 11652 | 7349 | 8373 | 8835 | 3421 |
| **Erythrobacter** | 15497 | 12834 | 9935 | 11295 | 7595 | 8438 | 8576 | 3375 |
| **Porphyrobacter** | 11769 | 8047 | 6488 | 7027 | 5584 | 5630 | 5315 | 2370 |
| **Croceicoccus** | 6647 | 5160 | 4024 | 4618 | 2608 | 3295 | 3305 | 1220 |
| **Rhizobium** | 107812 | 69086 | 53894 | 176433 | 61446 | 50908 | 56575 | 22551 |
| **Agrobacterium** | 49369 | 31409 | 20133 | 65226 | 22198 | 34265 | 60888 | 16859 |
| **Neorhizobium** | 13246 | 8752 | 7506 | 25210 | 8578 | 6600 | 7773 | 3695 |
| **Sinorhizobium** | 78499 | 62633 | 38566 | 240200 | 45667 | 55303 | 64386 | 15567 |
| **Ensifer** | 30756 | 18876 | 14701 | 73892 | 25193 | 36418 | 24042 | 4043 |
| **Shinella** | 71676 | 51318 | 51765 | 301418 | 46739 | 43017 | 49971 | 11111 |
| **Liberibacter** | 306 | 208 | 110 | 241 | 277 | 250 | 302 | 263 |
| **Bosea** | 133744 | 115627 | 104354 | 325463 | 230968 | 67955 | 106940 | 21617 |
| **Bradyrhizobium** | 82499 | 75080 | 41065 | 85966 | 102791 | 49578 | 60862 | 17218 |
| **Rhodopseudomonas** | 23070 | 16265 | 9575 | 22741 | 21169 | 10662 | 12913 | 4626 |
| **Nitrobacter** | 4157 | 4051 | 1927 | 6883 | 5204 | 2265 | 3535 | 953 |
| **Oligotropha** | 3505 | 3285 | 1651 | 3644 | 4836 | 1675 | 2439 | 615 |
| **Afipia** | 2296 | 2113 | 876 | 2240 | 3115 | 1137 | 1794 | 702 |
| **Variibacter** | 1656 | 1425 | 884 | 2016 | 1773 | 927 | 1177 | 392 |
| **Mesorhizobium** | 66158 | 51062 | 29743 | 96259 | 50310 | 30963 | 41958 | 14066 |
| **Aminobacter** | 16839 | 16089 | 7622 | 36977 | 14752 | 9372 | 13352 | 3587 |
| **Hoeflea** | 2366 | 1627 | 1256 | 3334 | 1435 | 1333 | 1555 | 801 |
| **Chelativorans** | 2362 | 2198 | 1592 | 5072 | 1894 | 1424 | 1805 | 689 |
| **Nitratireductor** | 2336 | 1803 | 1539 | 3260 | 1498 | 1594 | 1527 | 750 |
| **Phyllobacterium** | 1580 | 1133 | 789 | 2590 | 1048 | 889 | 961 | 424 |
| **Methylobacterium** | 42643 | 40436 | 20972 | 55474 | 50859 | 21989 | 24989 | 7120 |
| **Methylorubrum** | 8385 | 7590 | 4506 | 11893 | 7908 | 5231 | 5579 | 1781 |
| **Microvirga** | 5008 | 4185 | 2657 | 7085 | 4398 | 3000 | 3146 | 1100 |
| **Devosia** | 10889 | 9364 | 8009 | 13423 | 8654 | 5631 | 5727 | 2981 |
| **Hyphomicrobium** | 5334 | 4206 | 2442 | 4628 | 9439 | 3466 | 3691 | 1195 |
| **Blastochloris** | 4243 | 3437 | 2519 | 7915 | 3368 | 3230 | 2578 | 997 |
| **Rhodoplanes** | 2491 | 2071 | 1579 | 3245 | 2323 | 1419 | 1598 | 708 |
| **Pelagibacterium** | 1033 | 818 | 784 | 1532 | 680 | 679 | 651 | 378 |
| **Rhodomicrobium** | 1031 | 793 | 526 | 1278 | 620 | 654 | 570 | 252 |
| **Filomicrobium** | 362 | 311 | 184 | 382 | 251 | 286 | 170 | 112 |
| **Maritalea** | 282 | 248 | 216 | 278 | 122 | 246 | 212 | 290 |
| **Xanthobacter** | 8412 | 7408 | 2705 | 8144 | 7972 | 4740 | 3836 | 2277 |
| **Azorhizobium** | 4707 | 3746 | 2157 | 6055 | 5414 | 3045 | 2971 | 829 |
| **Starkeya** | 3872 | 3624 | 1937 | 6271 | 3651 | 2778 | 2442 | 911 |

| | | | | | | | | |
|---|---|---|---|---|---|---|---|---|
| **Pseudolabrys** | 2423 | 1941 | 1445 | 3135 | 2270 | 1315 | 1532 | 651 |
| **Pleomorphomonas** | 5915 | 3686 | 3423 | 23954 | 3293 | 4761 | 3498 | 1092 |
| **Methylocystis** | 4089 | 3014 | 2024 | 3680 | 3600 | 2192 | 2166 | 900 |
| **Methylosinus** | 2112 | 1692 | 1132 | 2538 | 2166 | 1321 | 1192 | 479 |
| **Ochrobactrum** | 8866 | 5680 | 5135 | 20738 | 3647 | 8437 | 5398 | 7359 |
| **Brucella** | 2237 | 1774 | 1474 | 3628 | 1483 | 1440 | 1431 | 904 |
| **Martelella** | 7519 | 4863 | 4783 | 12724 | 4363 | 4587 | 4720 | 1994 |
| **Aureimonas** | 2518 | 1968 | 1334 | 3937 | 1719 | 1540 | 1503 | 538 |
| **Hartmannibacter** | 3075 | 2379 | 1664 | 7954 | 2382 | 2316 | 2239 | 737 |
| **Pseudorhodoplanes** | 1854 | 1752 | 1733 | 2220 | 1956 | 1109 | 1272 | 529 |
| **Methyloceanibacter** | 1330 | 1085 | 660 | 1568 | 875 | 910 | 827 | 345 |
| **Chelatococcus** | 6021 | 4801 | 3441 | 9596 | 6243 | 3487 | 3947 | 1225 |
| **Methylocella** | 1407 | 1495 | 755 | 1668 | 1362 | 939 | 865 | 352 |
| **Methylovirgula** | 1246 | 1051 | 529 | 1215 | 1143 | 749 | 749 | 327 |
| **Beijerinckia** | 853 | 955 | 419 | 841 | 890 | 605 | 598 | 317 |
| **Parvibaculum** | 1568 | 1768 | 1175 | 1809 | 952 | 989 | 1025 | 1084 |
| **Anderseniella** | 646 | 431 | 275 | 549 | 401 | 408 | 331 | 303 |
| **Candidatus Phaeomarinob** | 452 | 342 | 556 | 464 | 258 | 313 | 249 | 198 |
| **Breoghania** | 2016 | 1445 | 1136 | 3259 | 1295 | 1388 | 1118 | 509 |
| **Cohaesibacter** | 655 | 484 | 349 | 835 | 380 | 611 | 381 | 272 |
| **Bartonella** | 1405 | 1016 | 580 | 841 | 448 | 1330 | 1245 | 1384 |
| **Paracoccus** | 159716 | 93309 | 64487 | 114550 | 33489 | 84812 | 150593 | 30223 |
| **Rhodobacter** | 33255 | 15044 | 27048 | 24724 | 9567 | 14330 | 17775 | 10601 |
| **Pannonibacter** | 15673 | 11545 | 7165 | 18879 | 7404 | 8015 | 10309 | 1805 |
| **Defluviimonas** | 7840 | 5051 | 6790 | 6517 | 2855 | 3685 | 5897 | 1985 |
| **Celeribacter** | 7401 | 4767 | 4967 | 6472 | 2930 | 4137 | 5301 | 2570 |
| **Rhodovulum** | 6602 | 4331 | 4336 | 5885 | 2319 | 3789 | 4371 | 2126 |
| **Sulfitobacter** | 6382 | 3679 | 4912 | 4693 | 2209 | 3370 | 3987 | 3102 |
| **Tabrizicola** | 6339 | 2908 | 4751 | 4110 | 1479 | 1940 | 3092 | 3836 |
| **Phaeobacter** | 5243 | 2959 | 4600 | 4074 | 1856 | 2968 | 3460 | 2603 |
| **Gemmobacter** | 4140 | 1762 | 3891 | 2730 | 1100 | 1450 | 2444 | 2946 |
| **Yangia** | 4128 | 2640 | 2682 | 3880 | 1698 | 2472 | 2886 | 1421 |
| **Ruegeria** | 3226 | 1911 | 2642 | 2327 | 1102 | 1628 | 2058 | 1435 |
| **Thalassococcus** | 2963 | 1658 | 2157 | 2330 | 1140 | 1417 | 1831 | 1079 |
| **Ketogulonicigenium** | 2465 | 1515 | 2105 | 2001 | 823 | 1544 | 1926 | 1000 |
| **Labrenzia** | 2400 | 1629 | 1889 | 3139 | 1313 | 1469 | 1538 | 881 |
| **Sagittula** | 2277 | 1454 | 1398 | 2148 | 794 | 1206 | 1314 | 707 |
| **Pelagibaca** | 2245 | 1377 | 1583 | 2202 | 965 | 1247 | 1564 | 859 |
| **Salipiger** | 2237 | 1484 | 1377 | 2385 | 1030 | 1298 | 1444 | 627 |
| **Leisingera** | 2223 | 1280 | 1662 | 1835 | 896 | 1170 | 1512 | 1016 |

| | | | | | | | | |
|---|---|---|---|---|---|---|---|---|
| **Confluentimicrobium** | 2152 | 1105 | 1640 | 1832 | 637 | 986 | 1285 | 868 |
| **Thioclava** | 2150 | 1073 | 1450 | 1902 | 735 | 1194 | 1373 | 685 |
| **Marinovum** | 2105 | 1315 | 1769 | 1990 | 853 | 1206 | 1533 | 774 |
| **Dinoroseobacter** | 1689 | 951 | 1167 | 1240 | 659 | 747 | 947 | 588 |
| **Brevirhabdus** | 1664 | 934 | 1081 | 1140 | 612 | 700 | 890 | 505 |
| **Yoonia** | 1599 | 717 | 1197 | 1151 | 536 | 735 | 1026 | 656 |
| **Roseibacterium** | 1587 | 1588 | 1084 | 1390 | 603 | 975 | 1151 | 520 |
| **Silicimonas** | 1547 | 1004 | 1055 | 1401 | 701 | 794 | 1009 | 480 |
| **Roseovarius** | 1367 | 777 | 1104 | 1040 | 463 | 681 | 878 | 667 |
| **Stappia** | 1230 | 1030 | 766 | 2118 | 818 | 889 | 847 | 391 |
| **Antarctobacter** | 1159 | 659 | 787 | 960 | 396 | 667 | 785 | 542 |
| **Epibacterium** | 1142 | 684 | 763 | 813 | 398 | 616 | 702 | 525 |
| **Octadecabacter** | 1071 | 610 | 983 | 845 | 341 | 569 | 744 | 685 |
| **Roseobacter** | 1051 | 865 | 706 | 756 | 443 | 573 | 851 | 522 |
| **Tateyamaria** | 951 | 525 | 659 | 721 | 365 | 447 | 476 | 369 |
| **Rhodobaca** | 920 | 584 | 722 | 735 | 303 | 467 | 718 | 613 |
| **Jannaschia** | 801 | 417 | 628 | 637 | 336 | 436 | 493 | 348 |
| **Sedimentitalea** | 435 | 305 | 393 | 323 | 162 | 296 | 331 | 282 |
| **Pseudovibrio** | 362 | 276 | 306 | 415 | 223 | 339 | 266 | 238 |
| **Halocynthiibacter** | 312 | 144 | 212 | 190 | 131 | 149 | 167 | 169 |
| **Marivivens** | 289 | 400 | 197 | 275 | 148 | 147 | 286 | 164 |
| **Hyphomonas** | 1887 | 1809 | 1162 | 1611 | 1026 | 1187 | 1177 | 624 |
| **Glycocaulis** | 800 | 672 | 360 | 675 | 337 | 524 | 459 | 258 |
| **Maricaulis** | 591 | 584 | 378 | 720 | 310 | 439 | 347 | 188 |
| **Hirschia** | 125 | 101 | 168 | 59 | 29 | 109 | 63 | 118 |
| **Brevundimonas** | 158265 | 313413 | 217941 | 283348 | 111888 | 66683 | 81594 | 44327 |
| **Caulobacter** | 42663 | 58210 | 17729 | 36679 | 28130 | 20476 | 25436 | 9836 |
| **Phenylobacterium** | 7126 | 10638 | 4077 | 7211 | 7515 | 3865 | 5688 | 2481 |
| **Asticcacaulis** | 1742 | 1936 | 1268 | 1628 | 919 | 1134 | 1116 | 596 |
| **Azospirillum** | 19924 | 15906 | 11027 | 21841 | 11494 | 12629 | 12323 | 5611 |
| **Magnetospirillum** | 7193 | 5003 | 3614 | 6576 | 3846 | 4648 | 4012 | 1941 |
| **Rhodospirillum** | 2972 | 2556 | 1801 | 3578 | 1809 | 2123 | 2106 | 913 |
| **Tistrella** | 2471 | 2048 | 1470 | 2794 | 1451 | 1463 | 1666 | 714 |
| **Nitrospirillum** | 2434 | 2410 | 1402 | 3150 | 1638 | 2152 | 1657 | 735 |
| **Niveispirillum** | 2000 | 2806 | 1161 | 2081 | 1885 | 1361 | 1343 | 629 |
| **Thalassospira** | 1836 | 1514 | 1238 | 1612 | 769 | 1333 | 1211 | 835 |
| **Indioceanicola** | 1311 | 1222 | 776 | 1262 | 837 | 976 | 877 | 431 |
| **Pararhodospirillum** | 934 | 779 | 592 | 1269 | 787 | 737 | 617 | 345 |
| **Magnetospira** | 471 | 374 | 279 | 549 | 255 | 356 | 282 | 191 |
| **Haematospirillum** | 163 | 126 | 87 | 168 | 77 | 144 | 96 | 70 |

| | | | | | | | | |
|---|---|---|---|---|---|---|---|---|
| **Candidatus Endolissoclinu** | 74 | 88 | 106 | 36 | 29 | 64 | 55 | 82 |
| **Roseomonas** | 9933 | 6795 | 5249 | 10634 | 6549 | 5158 | 6985 | 2779 |
| **Komagataeibacter** | 3793 | 3118 | 2631 | 3800 | 1861 | 3178 | 2542 | 1390 |
| **Acetobacter** | 2205 | 1852 | 1615 | 2350 | 1081 | 2594 | 1724 | 1340 |
| **Acidiphilium** | 1957 | 1574 | 1283 | 2201 | 1272 | 1088 | 1221 | 541 |
| **Acidisphaera** | 1395 | 1128 | 959 | 1417 | 848 | 788 | 869 | 324 |
| **Gluconacetobacter** | 1233 | 1122 | 820 | 1359 | 686 | 1486 | 843 | 353 |
| **Granulibacter** | 1174 | 943 | 714 | 1135 | 584 | 824 | 690 | 381 |
| **Gluconobacter** | 1099 | 1012 | 633 | 983 | 579 | 1039 | 652 | 477 |
| **Kozakia** | 509 | 337 | 291 | 441 | 223 | 378 | 267 | 226 |
| **Neoasaia** | 490 | 405 | 277 | 391 | 212 | 417 | 261 | 140 |
| **Asaia** | 236 | 205 | 219 | 226 | 103 | 250 | 155 | 146 |
| **Parasaccharibacter** | 217 | 174 | 95 | 141 | 70 | 214 | 85 | 80 |
| **Commensalibacter** | 72 | 62 | 27 | 15 | 23 | 89 | 40 | 71 |
| **Polymorphum** | 3068 | 1872 | 1660 | 5617 | 1705 | 1847 | 1905 | 669 |
| **Phreatobacter** | 2761 | 2527 | 2755 | 4110 | 2952 | 1689 | 1723 | 665 |
| **Micavibrio** | 655 | 588 | 825 | 596 | 262 | 509 | 481 | 292 |
| **Candidatus Puniceispirillu** | 159 | 109 | 109 | 86 | 50 | 128 | 79 | 101 |
| **Rickettsia** | 670 | 464 | 256 | 263 | 160 | 639 | 579 | 430 |
| **Orientia** | 92 | 91 | 148 | 43 | 42 | 85 | 60 | 65 |
| **Candidatus Phycoricketts** | 67 | 55 | 36 | 35 | 14 | 65 | 38 | 29 |
| **Wolbachia** | 308 | 184 | 213 | 108 | 74 | 249 | 282 | 239 |
| **Ehrlichia** | 192 | 135 | 63 | 91 | 39 | 221 | 144 | 214 |
| **Anaplasma** | 87 | 74 | 31 | 16 | 20 | 111 | 48 | 54 |
| **Neorickettsia** | 67 | 44 | 21 | 56 | 7 | 51 | 29 | 29 |
| **Candidatus Midichloria** | 35 | 48 | 8 | 41 | 10 | 34 | 19 | 23 |
| **Candidatus Fokinia** | 28 | 17 | 9 | 6 | 6 | 9 | 9 | 6 |
| **Candidatus Pelagibacter** | 326 | 320 | 111 | 183 | 91 | 266 | 598 | 419 |
| **Candidatus Fonsibacter** | 117 | 153 | 27 | 65 | 74 | 125 | 86 | 553 |
| **Magnetococcus** | 293 | 181 | 156 | 141 | 61 | 281 | 161 | 176 |
| **Parvularcula** | 271 | 193 | 116 | 201 | 139 | 188 | 130 | 103 |
| **Candidatus Nucleicultrix** | 110 | 47 | 21 | 58 | 35 | 59 | 45 | 98 |
| **Candidatus Paracaedibact** | 100 | 76 | 330 | 63 | 36 | 63 | 51 | 53 |
| **Desulfovibrio** | 72801 | 20786 | 11924 | 17251 | 13481 | 33480 | 37447 | 5855 |
| **Pseudodesulfovibrio** | 2329 | 1608 | 999 | 1150 | 757 | 2465 | 1308 | 707 |
| **Lawsonia** | 129 | 67 | 30 | 20 | 26 | 172 | 59 | 72 |
| **Desulfomicrobium** | 1496 | 903 | 573 | 782 | 368 | 1485 | 782 | 819 |
| **Desulfohalobium** | 208 | 169 | 79 | 177 | 68 | 183 | 124 | 58 |
| **Myxococcus** | 5382 | 5084 | 3697 | 4458 | 42988 | 4210 | 3651 | 1888 |
| **Corallococcus** | 1741 | 1640 | 1015 | 1452 | 1106 | 1472 | 1190 | 652 |

| | | | | | | | | |
|---|---|---|---|---|---|---|---|---|
| **Archangium** | 1346 | 1280 | 1016 | 1357 | 729 | 1061 | 951 | 431 |
| **Cystobacter** | 1244 | 1191 | 736 | 1199 | 723 | 886 | 819 | 453 |
| **Stigmatella** | 955 | 915 | 577 | 879 | 579 | 740 | 659 | 412 |
| **Melittangium** | 854 | 829 | 518 | 720 | 446 | 724 | 592 | 322 |
| **Anaeromyxobacter** | 3716 | 3575 | 2618 | 2770 | 1762 | 3081 | 2294 | 986 |
| **Vulgatibacter** | 724 | 585 | 396 | 538 | 292 | 537 | 444 | 245 |
| **Sorangium** | 6417 | 6079 | 4459 | 5727 | 3373 | 4436 | 4319 | 1842 |
| **Chondromyces** | 1018 | 1060 | 773 | 1021 | 521 | 850 | 750 | 334 |
| **Pajaroellobacter** | 30 | 30 | 16 | 24 | 8 | 31 | 11 | 21 |
| **Sandaracinus** | 1588 | 1538 | 1134 | 1202 | 819 | 1083 | 1013 | 381 |
| **Haliangium** | 1627 | 1570 | 1088 | 1563 | 828 | 1258 | 1143 | 730 |
| **Geobacter** | 6789 | 4901 | 3085 | 3761 | 2237 | 6653 | 3494 | 1898 |
| **Geoalkalibacter** | 553 | 354 | 243 | 263 | 146 | 573 | 251 | 179 |
| **Pelobacter** | 1882 | 1407 | 881 | 866 | 574 | 2070 | 990 | 750 |
| **Desulfuromonas** | 1255 | 1450 | 818 | 758 | 508 | 1317 | 904 | 539 |
| **Desulfococcus** | 925 | 686 | 496 | 574 | 287 | 867 | 473 | 327 |
| **Desulfobacter** | 508 | 299 | 636 | 145 | 166 | 607 | 307 | 225 |
| **Desulfatibacillum** | 462 | 296 | 183 | 177 | 123 | 446 | 254 | 162 |
| **Desulfobacula** | 247 | 210 | 228 | 98 | 83 | 354 | 193 | 112 |
| **Desulfobacterium** | 194 | 178 | 124 | 81 | 60 | 234 | 93 | 123 |
| **Desulfobulbus** | 726 | 454 | 251 | 318 | 174 | 550 | 414 | 154 |
| **Desulfurivibrio** | 463 | 413 | 260 | 492 | 242 | 547 | 474 | 263 |
| **Desulfotalea** | 250 | 363 | 98 | 675 | 107 | 510 | 547 | 196 |
| **Desulfocapsa** | 159 | 180 | 106 | 122 | 48 | 177 | 350 | 65 |
| **Syntrophus** | 256 | 181 | 119 | 86 | 85 | 246 | 113 | 71 |
| **Desulfomonile** | 228 | 194 | 108 | 92 | 68 | 276 | 193 | 135 |
| **Desulfobacca** | 150 | 123 | 97 | 54 | 53 | 145 | 86 | 124 |
| **Syntrophobacter** | 452 | 316 | 250 | 241 | 166 | 464 | 212 | 168 |
| **Desulfarculus** | 1037 | 732 | 483 | 668 | 345 | 1005 | 532 | 337 |
| **Bradymonas** | 488 | 412 | 285 | 368 | 166 | 423 | 301 | 131 |
| **Desulfurella** | 104 | 86 | 35 | 35 | 25 | 108 | 66 | 66 |
| **Hippea** | 93 | 78 | 35 | 25 | 17 | 104 | 113 | 31 |
| **Candidatus Desulfofervidu** | 121 | 67 | 80 | 48 | 31 | 90 | 97 | 55 |
| **Arcobacter** | 16301 | 3525 | 4144 | 4532 | 7171 | 5125 | 11221 | 358481 |
| **Aliiarcobacter** | 227 | 80 | 54 | 68 | 88 | 120 | 146 | 3252 |
| **Campylobacter** | 9209 | 5364 | 4466 | 2735 | 2028 | 14133 | 6578 | 4816 |
| **Sulfurospirillum** | 1726 | 1168 | 719 | 1422 | 2685 | 1566 | 1477 | 2302 |
| **Helicobacter** | 1856 | 1351 | 1513 | 976 | 757 | 1727 | 1662 | 1807 |
| **Sulfuricurvum** | 409 | 132 | 47 | 56 | 121 | 192 | 91 | 195 |
| **Sulfurimonas** | 330 | 237 | 123 | 132 | 83 | 285 | 160 | 1240 |

| | | | | | | | | |
|---|---|---|---|---|---|---|---|---|
| **Wolinella** | 112 | 127 | 64 | 70 | 104 | 108 | 73 | 50 |
| **Nitratifractor** | 198 | 170 | 196 | 93 | 39 | 260 | 159 | 66 |
| **Nautilia** | 291 | 208 | 120 | 81 | 57 | 287 | 228 | 161 |
| **Sulfurovum** | 141 | 136 | 58 | 43 | 57 | 225 | 99 | 91 |
| **Nitratiruptor** | 77 | 42 | 25 | 112 | 19 | 73 | 42 | 60 |
| **Bacteriovorax** | 3728 | 546 | 826 | 803 | 518 | 379 | 571 | 361 |
| **Halobacteriovorax** | 418 | 241 | 672 | 98 | 99 | 272 | 166 | 344 |
| **Bdellovibrio** | 2231 | 1290 | 1219 | 748 | 577 | 1084 | 1022 | 491 |
| **Hydrogenophilus** | 4389 | 581 | 225 | 1320 | 494 | 1334 | 884 | 277 |
| **Acidithiobacillus** | 3287 | 1676 | 1305 | 3193 | 648 | 2163 | 2160 | 1984 |
| **Mariprofundus** | 412 | 246 | 246 | 213 | 156 | 388 | 279 | 199 |
| **Faecalibacterium** | 1834440 | 768956 | 468748 | 300790 | 449834 | 2120253 | 608140 | 136167 |
| **Ruminococcus** | 106606 | 35763 | 39703 | 8717 | 6917 | 374392 | 30438 | 3220 |
| **Flavonifractor** | 31147 | 28451 | 14465 | 6419 | 7519 | 58547 | 14088 | 2700 |
| **Ethanoligenens** | 4876 | 3208 | 1442 | 915 | 843 | 7476 | 2151 | 650 |
| **Caproiciproducens** | 4484 | 2697 | 1414 | 829 | 611 | 6920 | 2064 | 699 |
| **Acutalibacter** | 2 | 1 | 1 | 0 | 2 | 4 | 0 | 1 |
| **Lachnoclostridium** | 136804 | 54890 | 32232 | 23728 | 43475 | 172774 | 65896 | 7782 |
| **Anaerostipes** | 95153 | 53465 | 33849 | 50516 | 23556 | 150250 | 41182 | 10727 |
| **Roseburia** | 91714 | 64520 | 32725 | 18949 | 20122 | 153149 | 49064 | 6744 |
| **Blautia** | 55003 | 36936 | 20558 | 13573 | 21296 | 92226 | 28324 | 5925 |
| **Butyrivibrio** | 11252 | 7737 | 3531 | 2230 | 2391 | 17207 | 4744 | 1352 |
| **Lachnoanaerobaculum** | 2947 | 1910 | 873 | 570 | 667 | 4324 | 1346 | 336 |
| **Cellulosilyticum** | 2382 | 1445 | 569 | 469 | 394 | 3106 | 1093 | 439 |
| **Anaerotignum** | 2024 | 1073 | 624 | 362 | 385 | 3190 | 909 | 253 |
| **Herbinix** | 1781 | 1317 | 561 | 363 | 481 | 2660 | 868 | 261 |
| **Eubacterium** | 138736 | 148497 | 53136 | 32232 | 41506 | 235772 | 80061 | 11519 |
| **Acetobacterium** | 1216 | 752 | 668 | 305 | 400 | 1609 | 713 | 706 |
| **Intestinimonas** | 42696 | 31155 | 13718 | 6914 | 5269 | 75108 | 20427 | 4148 |
| **Monoglobus** | 4129 | 2917 | 1179 | 920 | 765 | 6715 | 3817 | 995 |
| **Clostridium** | 60649 | 42248 | 21344 | 14885 | 21548 | 73365 | 33727 | 19187 |
| **Mordavella** | 15149 | 9188 | 4765 | 3804 | 3976 | 29023 | 7418 | 1651 |
| **Alkaliphilus** | 1153 | 618 | 406 | 184 | 227 | 1136 | 498 | 567 |
| **Geosporobacter** | 979 | 493 | 740 | 369 | 176 | 991 | 377 | 519 |
| **Candidatus Arthromitus** | 430 | 297 | 140 | 140 | 88 | 450 | 251 | 200 |
| **Oscillibacter** | 63294 | 56306 | 29374 | 12689 | 10638 | 156479 | 38731 | 7574 |
| **Clostridioides** | 46052 | 26973 | 15095 | 9404 | 9949 | 75928 | 23848 | 4775 |
| **Acetoanaerobium** | 9774 | 490 | 2923 | 319 | 354 | 1186 | 779 | 1803 |
| **Paeniclostridium** | 1205 | 721 | 311 | 229 | 311 | 1453 | 961 | 402 |
| **Peptoclostridium** | 1138 | 477 | 1119 | 196 | 156 | 1058 | 313 | 645 |

| | | | | | | | | |
|---|---|---|---|---|---|---|---|---|
| **Filifactor** | 664 | 424 | 225 | 177 | 163 | 896 | 379 | 119 |
| **Christensenella** | 25516 | 18857 | 8721 | 4865 | 4186 | 44010 | 12640 | 2669 |
| **Desulfitobacterium** | 3117 | 1568 | 1386 | 678 | 550 | 3406 | 1285 | 652 |
| **Syntrophobotulus** | 2242 | 362 | 245 | 167 | 154 | 845 | 296 | 127 |
| **Desulfosporosinus** | 1935 | 1117 | 1144 | 470 | 390 | 2019 | 1035 | 439 |
| **Desulfotomaculum** | 1712 | 1008 | 584 | 544 | 425 | 1938 | 751 | 494 |
| **Dehalobacterium** | 1677 | 708 | 787 | 327 | 246 | 1710 | 613 | 384 |
| **Dehalobacter** | 1197 | 690 | 452 | 258 | 245 | 1650 | 669 | 262 |
| **Desulfallas** | 739 | 417 | 241 | 175 | 143 | 913 | 315 | 154 |
| **Desulfofarcimen** | 448 | 260 | 188 | 157 | 99 | 421 | 218 | 105 |
| **Candidatus Desulforudis** | 386 | 329 | 247 | 169 | 84 | 520 | 202 | 150 |
| **Thermincola** | 358 | 184 | 172 | 140 | 62 | 416 | 164 | 125 |
| **Hungateiclostridium** | 2896 | 1501 | 1345 | 637 | 531 | 3245 | 1000 | 637 |
| **Mageeibacillus** | 1417 | 919 | 439 | 332 | 275 | 1806 | 723 | 222 |
| **Pseudoclostridium** | 1127 | 638 | 356 | 236 | 231 | 1364 | 450 | 140 |
| **Ruminiclostridium** | 844 | 511 | 265 | 155 | 166 | 1167 | 388 | 200 |
| **Thermoclostridium** | 749 | 490 | 274 | 147 | 166 | 1178 | 330 | 136 |
| **Fastidiosipila** | 570 | 326 | 111 | 98 | 77 | 601 | 259 | 193 |
| **Aminipila** | 2126 | 1050 | 614 | 485 | 392 | 2654 | 861 | 357 |
| **Mogibacterium** | 1571 | 749 | 411 | 369 | 326 | 1881 | 671 | 214 |
| **Thermaerobacter** | 696 | 589 | 378 | 428 | 220 | 924 | 484 | 213 |
| **Carboxydocella** | 512 | 283 | 167 | 125 | 139 | 542 | 212 | 92 |
| **Symbiobacterium** | 1186 | 939 | 630 | 523 | 308 | 1496 | 657 | 265 |
| **Heliobacterium** | 1143 | 696 | 430 | 311 | 211 | 1301 | 473 | 195 |
| **Syntrophomonas** | 268 | 141 | 105 | 69 | 48 | 295 | 115 | 163 |
| **Syntrophothermus** | 168 | 118 | 45 | 40 | 36 | 214 | 68 | 107 |
| **Caldicellulosiruptor** | 1993 | 1212 | 795 | 583 | 576 | 2305 | 1075 | 626 |
| **Thermoanaerobacterium** | 1731 | 902 | 728 | 447 | 766 | 1597 | 799 | 443 |
| **Thermosediminibacter** | 364 | 182 | 105 | 82 | 36 | 407 | 145 | 150 |
| **Thermoanaerobacter** | 1073 | 608 | 372 | 318 | 251 | 1109 | 488 | 556 |
| **Moorella** | 721 | 312 | 197 | 278 | 149 | 649 | 296 | 138 |
| **Ammonifex** | 245 | 178 | 143 | 107 | 73 | 332 | 107 | 49 |
| **Tepidanaerobacter** | 356 | 180 | 94 | 82 | 63 | 436 | 169 | 211 |
| **Thermacetogenium** | 336 | 217 | 149 | 126 | 89 | 391 | 156 | 86 |
| **Caldanaerobacter** | 315 | 200 | 94 | 77 | 51 | 311 | 125 | 105 |
| **Thermanaeromonas** | 179 | 103 | 120 | 49 | 50 | 215 | 96 | 58 |
| **Carboxydothermus** | 147 | 98 | 54 | 33 | 37 | 173 | 72 | 43 |
| **Mahella** | 589 | 263 | 153 | 94 | 78 | 574 | 166 | 208 |
| **Thermodesulfobium** | 152 | 198 | 47 | 84 | 48 | 171 | 124 | 106 |
| **Halanaerobium** | 509 | 407 | 176 | 128 | 133 | 507 | 378 | 286 |

| | | | | | | | | |
|---|---|---|---|---|---|---|---|---|
| **Halocella** | 331 | 202 | 83 | 113 | 71 | 380 | 142 | 104 |
| **Halothermothrix** | 148 | 118 | 36 | 37 | 36 | 160 | 76 | 68 |
| **Halobacteroides** | 356 | 197 | 92 | 81 | 59 | 277 | 470 | 170 |
| **Acetohalobium** | 352 | 150 | 93 | 81 | 72 | 305 | 129 | 80 |
| **Anoxybacter** | 357 | 211 | 129 | 127 | 62 | 338 | 158 | 106 |
| **Natranaerobius** | 229 | 134 | 61 | 42 | 47 | 206 | 205 | 90 |
| **Lactobacillus** | 109132 | 18997 | 18877 | 22959 | 165652 | 34166 | 34892 | 14491 |
| **Pediococcus** | 5240 | 510 | 934 | 982 | 991 | 871 | 480 | 825 |
| **Streptococcus** | 61959 | 19556 | 21987 | 15664 | 22571 | 51253 | 37344 | 33483 |
| **Lactococcus** | 9212 | 14681 | 9311 | 9205 | 8212 | 15436 | 13572 | 515659 |
| **Enterococcus** | 47805 | 18690 | 33515 | 227459 | 303323 | 42718 | 32440 | 19397 |
| **Tetragenococcus** | 958 | 571 | 2081 | 380 | 220 | 1670 | 770 | 1089 |
| **Vagococcus** | 534 | 286 | 1285 | 216 | 153 | 799 | 342 | 766 |
| **Melissococcus** | 284 | 127 | 397 | 57 | 96 | 378 | 254 | 291 |
| **Carnobacterium** | 1582 | 948 | 10687 | 4772 | 561 | 3233 | 2267 | 8333 |
| **Jeotgalibaca** | 1091 | 591 | 14600 | 866 | 406 | 3653 | 1551 | 7192 |
| **Marinilactibacillus** | 284 | 199 | 1348 | 138 | 112 | 565 | 245 | 554 |
| **Leuconostoc** | 1099 | 875 | 2050 | 572 | 552 | 1071 | 680 | 2476 |
| **Weissella** | 661 | 415 | 778 | 208 | 368 | 701 | 532 | 660 |
| **Oenococcus** | 385 | 300 | 591 | 191 | 142 | 477 | 375 | 236 |
| **Aerococcus** | 1323 | 810 | 7345 | 750 | 1180 | 2128 | 1373 | 4214 |
| **Bacillus** | 30526 | 18269 | 24339 | 11460 | 10593 | 30296 | 18251 | 19042 |
| **Anoxybacillus** | 2813 | 832 | 818 | 1146 | 4163 | 1645 | 1101 | 617 |
| **Geobacillus** | 2657 | 1560 | 1828 | 965 | 1540 | 2718 | 1656 | 1075 |
| **Virgibacillus** | 1859 | 1153 | 1305 | 607 | 512 | 2093 | 1088 | 1330 |
| **Lysinibacillus** | 1752 | 1032 | 1061 | 531 | 438 | 1793 | 978 | 1412 |
| **Oceanobacillus** | 910 | 657 | 636 | 291 | 234 | 1044 | 739 | 492 |
| **Halobacillus** | 882 | 693 | 823 | 824 | 212 | 1081 | 640 | 686 |
| **Fictibacillus** | 582 | 363 | 270 | 187 | 168 | 584 | 323 | 249 |
| **Lentibacillus** | 363 | 155 | 102 | 81 | 67 | 266 | 141 | 83 |
| **Parageobacillus** | 362 | 166 | 355 | 156 | 311 | 310 | 174 | 189 |
| **Paraliobacillus** | 306 | 148 | 158 | 58 | 51 | 249 | 111 | 167 |
| **Aeribacillus** | 304 | 176 | 145 | 104 | 179 | 388 | 150 | 110 |
| **Salimicrobium** | 289 | 162 | 184 | 77 | 56 | 320 | 109 | 130 |
| **Terribacillus** | 259 | 189 | 309 | 136 | 57 | 352 | 167 | 253 |
| **Amphibacillus** | 222 | 151 | 568 | 68 | 61 | 312 | 126 | 189 |
| **Paenibacillus** | 26596 | 18425 | 17066 | 9349 | 7097 | 34451 | 14316 | 9824 |
| **Brevibacillus** | 2807 | 1895 | 1475 | 944 | 699 | 2949 | 1629 | 1275 |
| **Thermobacillus** | 1314 | 1001 | 528 | 467 | 316 | 1623 | 643 | 243 |
| **Cohnella** | 1049 | 722 | 495 | 464 | 227 | 1264 | 515 | 316 |

| | | | | | | | | |
|---|---|---|---|---|---|---|---|---|
| **Aneurinibacillus** | 754 | 386 | 302 | 735 | 187 | 905 | 474 | 323 |
| **Staphylococcus** | 8277 | 4847 | 8043 | 2809 | 2376 | 7408 | 5467 | 6581 |
| **Macrococcus** | 566 | 332 | 503 | 200 | 117 | 610 | 274 | 489 |
| **Salinicoccus** | 237 | 146 | 139 | 59 | 49 | 288 | 144 | 86 |
| **Jeotgalicoccus** | 181 | 123 | 93 | 49 | 53 | 266 | 93 | 80 |
| **Auricoccus** | 133 | 114 | 199 | 35 | 31 | 134 | 155 | 72 |
| **Planococcus** | 2436 | 2178 | 3512 | 1188 | 598 | 2943 | 1404 | 2427 |
| **Sporosarcina** | 1439 | 911 | 1170 | 667 | 373 | 1813 | 1097 | 874 |
| **Solibacillus** | 721 | 381 | 497 | 165 | 174 | 754 | 380 | 401 |
| **Kurthia** | 444 | 1253 | 293 | 168 | 85 | 406 | 303 | 881 |
| **Jeotgalibacillus** | 395 | 236 | 189 | 91 | 64 | 423 | 301 | 103 |
| **Ureibacillus** | 362 | 195 | 161 | 72 | 69 | 306 | 144 | 192 |
| **Rummeliibacillus** | 207 | 154 | 649 | 70 | 35 | 174 | 132 | 176 |
| **Paenisporosarcina** | 159 | 79 | 148 | 112 | 43 | 132 | 215 | 102 |
| **Alicyclobacillus** | 1541 | 713 | 354 | 838 | 1662 | 1261 | 905 | 224 |
| **Tumebacillus** | 1470 | 1064 | 599 | 484 | 490 | 1778 | 710 | 364 |
| **Kyrpidia** | 333 | 251 | 212 | 159 | 95 | 455 | 160 | 120 |
| **Listeria** | 2142 | 1274 | 2600 | 670 | 512 | 2426 | 1411 | 2608 |
| **Brochothrix** | 287 | 160 | 274 | 377 | 78 | 290 | 202 | 440 |
| **Exiguobacterium** | 932 | 771 | 2156 | 597 | 281 | 1182 | 639 | 972 |
| **Gemella** | 892 | 550 | 300 | 289 | 211 | 864 | 424 | 436 |
| **Laceyella** | 209 | 171 | 182 | 76 | 65 | 297 | 139 | 107 |
| **Novibacillus** | 205 | 130 | 129 | 65 | 41 | 256 | 96 | 93 |
| **Sporolactobacillus** | 427 | 219 | 201 | 95 | 83 | 430 | 138 | 172 |
| **Veillonella** | 97926 | 13653 | 4294 | 19520 | 17109 | 22463 | 30962 | 3579 |
| **Dialister** | 48394 | 15197 | 9633 | 7037 | 5021 | 28288 | 15363 | 4184 |
| **Megasphaera** | 11725 | 7717 | 10082 | 2079 | 4427 | 10910 | 16430 | 6749 |
| **Negativicoccus** | 1249 | 649 | 455 | 364 | 381 | 771 | 679 | 186 |
| **Phascolarctobacterium** | 50118 | 21004 | 6828 | 12175 | 27479 | 65509 | 12642 | 6199 |
| **Acidaminococcus** | 6301 | 6370 | 5393 | 1832 | 2426 | 12969 | 14211 | 1156 |
| **Megamonas** | 17569 | 1347 | 596 | 856 | 477 | 1675 | 2949 | 671 |
| **Selenomonas** | 8409 | 7620 | 2986 | 2562 | 1590 | 10432 | 6569 | 3452 |
| **Pelosinus** | 2112 | 701 | 409 | 535 | 425 | 1118 | 718 | 670 |
| **Methylomusa** | 1282 | 449 | 241 | 377 | 206 | 777 | 388 | 207 |
| **Faecalitalea** | 24460 | 12855 | 7972 | 5395 | 6312 | 38398 | 12105 | 2628 |
| **Faecalibaculum** | 1726 | 1098 | 740 | 444 | 451 | 2629 | 855 | 217 |
| **Turicibacter** | 1438 | 1077 | 445 | 314 | 420 | 2056 | 710 | 355 |
| **Erysipelothrix** | 1223 | 567 | 559 | 578 | 179 | 1001 | 447 | 419 |
| **Erysipelatoclostridium** | 61 | 10 | 6 | 27 | 36 | 81 | 38 | 3 |
| **Peptoniphilus** | 1606 | 866 | 609 | 350 | 668 | 1481 | 1129 | 359 |

| | | | | | | | | |
|---|---|---|---|---|---|---|---|---|
| **Finegoldia** | 1324 | 513 | 390 | 331 | 446 | 790 | 573 | 246 |
| **Parvimonas** | 873 | 478 | 267 | 251 | 199 | 843 | 406 | 255 |
| **Murdochiella** | 806 | 576 | 330 | 186 | 172 | 1216 | 485 | 142 |
| **Anaerococcus** | 699 | 418 | 183 | 176 | 291 | 729 | 394 | 165 |
| **Soehngenia** | 555 | 327 | 419 | 100 | 112 | 584 | 218 | 348 |
| **Sporanaerobacter** | 1215 | 443 | 338 | 192 | 180 | 1046 | 350 | 430 |
| **Gottschalkia** | 656 | 352 | 203 | 123 | 100 | 670 | 355 | 209 |
| **Ezakiella** | 712 | 240 | 139 | 93 | 249 | 289 | 248 | 95 |
| **Ndongobacter** | 717 | 496 | 231 | 218 | 123 | 1128 | 355 | 108 |
| **Limnochorda** | 617 | 476 | 313 | 308 | 190 | 711 | 297 | 158 |
| **Bifidobacterium** | 691410 | 139606 | 139402 | 271361 | 208482 | 358783 | 135467 | 80834 |
| **Gardnerella** | 13129 | 2786 | 1747 | 4650 | 5258 | 2004 | 4046 | 360 |
| **Parascardovia** | 338 | 223 | 183 | 203 | 679 | 498 | 184 | 77 |
| **Scardovia** | 236 | 130 | 67 | 60 | 66 | 279 | 95 | 29 |
| **Microbacterium** | 188515 | 395489 | 144098 | 2108052 | 386812 | 189936 | 1014282 | 40738 |
| **Leucobacter** | 7917 | 8132 | 16043 | 24245 | 5786 | 3404 | 8001 | 2388 |
| **Agromyces** | 7771 | 12989 | 10286 | 21332 | 7294 | 5503 | 10423 | 3571 |
| **Leifsonia** | 4819 | 7921 | 5986 | 11531 | 4302 | 3723 | 7793 | 2239 |
| **Rathayibacter** | 3576 | 4807 | 4286 | 7228 | 2893 | 2607 | 3929 | 1643 |
| **Clavibacter** | 3245 | 4340 | 4320 | 6163 | 2972 | 2312 | 3894 | 1761 |
| **Agrococcus** | 2918 | 3716 | 4086 | 5867 | 2383 | 1903 | 3284 | 1112 |
| **Curtobacterium** | 2873 | 3866 | 3742 | 5564 | 2199 | 2002 | 3194 | 1249 |
| **Cryobacterium** | 2772 | 3987 | 3183 | 6074 | 2306 | 2036 | 3469 | 1654 |
| **Lysinimonas** | 2685 | 3247 | 3653 | 5343 | 2405 | 1647 | 3263 | 1111 |
| **Microterricola** | 2606 | 4482 | 3363 | 6353 | 2278 | 1905 | 3309 | 1882 |
| **Frondihabitans** | 1938 | 2767 | 2668 | 5110 | 1712 | 1525 | 2498 | 1094 |
| **Plantibacter** | 1719 | 2885 | 2767 | 4730 | 1488 | 1223 | 2337 | 966 |
| **Gryllotalpicola** | 1260 | 2152 | 1568 | 2804 | 1212 | 922 | 2095 | 590 |
| **Salinibacterium** | 1070 | 1169 | 1131 | 3116 | 1112 | 756 | 2013 | 1040 |
| **Microcella** | 1008 | 1282 | 1391 | 3153 | 962 | 748 | 1711 | 553 |
| **Mycetocola** | 627 | 779 | 763 | 1355 | 433 | 485 | 693 | 373 |
| **Humibacter** | 519 | 611 | 659 | 1094 | 371 | 325 | 631 | 213 |
| **Cnuibacter** | 441 | 621 | 434 | 1163 | 436 | 323 | 648 | 247 |
| **Aurantimicrobium** | 330 | 286 | 275 | 473 | 186 | 255 | 234 | 221 |
| **Rhodoluna** | 164 | 106 | 145 | 121 | 79 | 92 | 85 | 107 |
| **Candidatus Aquiluna** | 65 | 54 | 39 | 83 | 26 | 88 | 45 | 53 |
| **Pontimonas** | 137 | 85 | 75 | 162 | 59 | 159 | 73 | 53 |
| **Arthrobacter** | 9729 | 11123 | 9147 | 11715 | 5485 | 6906 | 8116 | 5510 |
| **Kocuria** | 5006 | 5733 | 7577 | 7761 | 4071 | 4005 | 5161 | 2198 |
| **Rothia** | 2931 | 1531 | 750 | 1173 | 1859 | 1570 | 2199 | 558 |

| | | | | | | | | |
|---|---|---|---|---|---|---|---|---|
| **Pseudarthrobacter** | 2563 | 3923 | 2905 | 4151 | 1846 | 2251 | 2440 | 1047 |
| **Zhihengliuella** | 1709 | 3074 | 1623 | 11718 | 1133 | 1071 | 3401 | 296 |
| **Micrococcus** | 1488 | 1588 | 2488 | 2959 | 1766 | 1242 | 1493 | 782 |
| **Glutamicibacter** | 1349 | 1234 | 1399 | 2712 | 835 | 1157 | 1317 | 707 |
| **Sinomonas** | 1034 | 1408 | 1516 | 1908 | 853 | 806 | 1125 | 425 |
| **Auritidibacter** | 339 | 217 | 249 | 393 | 139 | 158 | 309 | 79 |
| **Neomicrococcus** | 290 | 282 | 294 | 286 | 167 | 242 | 172 | 169 |
| **Psychromicrobium** | 240 | 211 | 203 | 326 | 161 | 163 | 158 | 135 |
| **Renibacterium** | 145 | 118 | 92 | 98 | 54 | 100 | 107 | 66 |
| **Paenarthrobacter** | 110 | 136 | 140 | 172 | 131 | 73 | 136 | 57 |
| **Serinicoccus** | 3116 | 3335 | 5727 | 3793 | 2109 | 2006 | 2761 | 962 |
| **Janibacter** | 2585 | 2805 | 3458 | 3498 | 2026 | 1628 | 2529 | 1109 |
| **Arsenicicoccus** | 2010 | 1676 | 2688 | 1813 | 1177 | 1080 | 1587 | 611 |
| **Phycicoccus** | 1654 | 2221 | 1709 | 1624 | 1345 | 971 | 1430 | 736 |
| **Ornithinimicrobium** | 1635 | 1578 | 2411 | 1835 | 1166 | 980 | 1324 | 438 |
| **Intrasporangium** | 1561 | 1874 | 2253 | 1797 | 1332 | 1112 | 1326 | 659 |
| **Cellulosimicrobium** | 2675 | 2726 | 7308 | 3654 | 1910 | 2120 | 2333 | 1180 |
| **Isoptericola** | 2424 | 2403 | 5330 | 3099 | 1993 | 1924 | 2073 | 964 |
| **Xylanibacterium** | 1227 | 1372 | 2350 | 1933 | 1020 | 961 | 1167 | 429 |
| **Xylanimicrobium** | 1192 | 1223 | 2868 | 1537 | 875 | 880 | 1086 | 365 |
| **Xylanimonas** | 1041 | 1215 | 2354 | 1629 | 860 | 792 | 1061 | 385 |
| **Cellulomonas** | 5874 | 6302 | 21772 | 8761 | 4961 | 4769 | 5192 | 2250 |
| **Paraoerskovia** | 787 | 842 | 1858 | 1072 | 600 | 659 | 684 | 318 |
| **Brachybacterium** | 5459 | 5884 | 7414 | 7667 | 3810 | 4485 | 4960 | 1697 |
| **Dermabacter** | 290 | 225 | 470 | 710 | 188 | 227 | 268 | 111 |
| **Devriesea** | 215 | 195 | 179 | 232 | 134 | 207 | 209 | 102 |
| **Miniimonas** | 2528 | 2664 | 5326 | 7047 | 2032 | 2003 | 4621 | 1060 |
| **Beutenbergia** | 1327 | 1484 | 2734 | 2222 | 1008 | 945 | 1255 | 416 |
| **Brevibacterium** | 3706 | 4317 | 5205 | 12435 | 2693 | 2940 | 4826 | 1396 |
| **Kytococcus** | 1137 | 1417 | 2030 | 2565 | 795 | 745 | 1223 | 363 |
| **Dermacoccus** | 1095 | 1040 | 1771 | 1628 | 836 | 791 | 1055 | 672 |
| **Luteipulveratus** | 1007 | 1119 | 1159 | 1170 | 812 | 699 | 939 | 449 |
| **Sanguibacter** | 1264 | 1224 | 3234 | 1934 | 898 | 1102 | 1097 | 621 |
| **Georgenia** | 1139 | 964 | 1921 | 1582 | 792 | 945 | 1008 | 386 |
| **Austwickia** | 690 | 494 | 1009 | 676 | 438 | 507 | 604 | 229 |
| **Dermatophilus** | 271 | 194 | 297 | 167 | 158 | 195 | 165 | 136 |
| **Jonesia** | 220 | 160 | 333 | 295 | 107 | 161 | 264 | 97 |
| **Tropheryma** | 44 | 14 | 14 | 11 | 5 | 25 | 14 | 10 |
| **Mycobacterium** | 120027 | 72439 | 23556 | 57791 | 194806 | 46079 | 73063 | 16920 |
| **Mycolicibacterium** | 76707 | 48933 | 16254 | 41728 | 141509 | 30280 | 51943 | 12787 |

| | | | | | | | | |
|---|---|---|---|---|---|---|---|---|
| **Mycobacteroides** | 8484 | 5312 | 2047 | 4851 | 12488 | 3934 | 6295 | 1568 |
| **Mycolicibacter** | 7407 | 4774 | 1755 | 3962 | 8466 | 3446 | 5778 | 1135 |
| **Hoyosella** | 376 | 284 | 272 | 355 | 347 | 254 | 318 | 171 |
| **Rhodococcus** | 21602 | 24193 | 15306 | 31121 | 28731 | 14103 | 17470 | 8913 |
| **Nocardia** | 14805 | 12707 | 9643 | 13737 | 14370 | 8964 | 11147 | 4394 |
| **Corynebacterium** | 21785 | 17806 | 19373 | 20841 | 12243 | 19357 | 15559 | 7285 |
| **Gordonia** | 15236 | 11381 | 8543 | 11861 | 12338 | 7180 | 12110 | 3443 |
| **Dietzia** | 5134 | 2714 | 3280 | 3145 | 2513 | 2138 | 2548 | 2270 |
| **Tsukamurella** | 3221 | 2883 | 2624 | 3184 | 3457 | 1729 | 2521 | 1558 |
| **Segniliparus** | 423 | 497 | 255 | 327 | 341 | 388 | 317 | 143 |
| **Lawsonella** | 280 | 207 | 160 | 180 | 259 | 1439 | 179 | 68 |
| **Micropruina** | 43853 | 48015 | 33893 | 21004 | 30226 | 27926 | 55659 | 3846 |
| **Nocardioides** | 8681 | 9944 | 9601 | 9341 | 6455 | 5668 | 7706 | 2860 |
| **Aeromicrobium** | 5581 | 5097 | 6817 | 6052 | 3802 | 3582 | 4577 | 1862 |
| **Friedmanniella** | 4822 | 5743 | 7354 | 6270 | 3953 | 3319 | 5358 | 1158 |
| **Pimelobacter** | 2038 | 2331 | 2174 | 2332 | 1471 | 1270 | 1903 | 606 |
| **Kribbella** | 1838 | 2043 | 2066 | 2152 | 1333 | 1196 | 1709 | 461 |
| **Marmoricola** | 1443 | 1799 | 1817 | 1698 | 1161 | 903 | 1304 | 614 |
| **Actinopolymorpha** | 1152 | 1197 | 1295 | 1335 | 970 | 811 | 1044 | 361 |
| **Tessaracoccus** | 27056 | 25874 | 154037 | 42229 | 15644 | 30860 | 35589 | 6288 |
| **Microlunatus** | 17434 | 7418 | 8026 | 10917 | 3946 | 5294 | 16262 | 1746 |
| **Propionibacterium** | 7112 | 7736 | 11555 | 13742 | 7250 | 6235 | 8209 | 1802 |
| **Acidipropionibacterium** | 6133 | 5793 | 9581 | 7755 | 5351 | 4511 | 6572 | 1373 |
| **Auraticoccus** | 2263 | 2778 | 3984 | 3273 | 1762 | 1572 | 2398 | 561 |
| **Cutibacterium** | 2068 | 1814 | 2996 | 2137 | 1725 | 1822 | 1878 | 612 |
| **Pseudopropionibacterium** | 1939 | 1931 | 5584 | 2709 | 1420 | 1698 | 1845 | 450 |
| **Propionimicrobium** | 255 | 199 | 126 | 143 | 191 | 409 | 187 | 75 |
| **Streptomyces** | 101203 | 102004 | 94520 | 111361 | 73507 | 73500 | 84228 | 36807 |
| **Kitasatospora** | 4113 | 4184 | 3520 | 4265 | 2667 | 2946 | 3559 | 1375 |
| **Streptacidiphilus** | 995 | 855 | 761 | 925 | 593 | 817 | 812 | 329 |
| **Amycolatopsis** | 10267 | 10412 | 8046 | 11435 | 7968 | 6902 | 8451 | 3415 |
| **Pseudonocardia** | 9095 | 8547 | 7151 | 9080 | 6605 | 5312 | 7990 | 2618 |
| **Actinoalloteichus** | 3511 | 3133 | 2619 | 4562 | 2141 | 2367 | 2852 | 1128 |
| **Saccharomonospora** | 3120 | 3171 | 2734 | 3256 | 2402 | 2395 | 2650 | 1157 |
| **Actinosynnema** | 2141 | 2049 | 1850 | 2328 | 1472 | 1596 | 1806 | 853 |
| **Kutzneria** | 1589 | 1492 | 1325 | 1498 | 1165 | 1000 | 1253 | 479 |
| **Lentzea** | 1568 | 1492 | 1755 | 1576 | 1062 | 953 | 1171 | 451 |
| **Saccharothrix** | 1367 | 1429 | 1174 | 1546 | 1013 | 937 | 1238 | 580 |
| **Kibdelosporangium** | 1348 | 1494 | 972 | 1389 | 955 | 882 | 1084 | 476 |
| **Saccharopolyspora** | 1328 | 1461 | 1369 | 1562 | 1037 | 843 | 1104 | 418 |

| | | | | | | | | |
|---|---|---|---|---|---|---|---|---|
| **Allokutzneria** | 1305 | 1250 | 1084 | 1527 | 866 | 867 | 974 | 385 |
| **Alloactinosynnema** | 1195 | 1119 | 953 | 1232 | 920 | 780 | 986 | 399 |
| **Prauserella** | 769 | 691 | 726 | 896 | 562 | 620 | 620 | 269 |
| **Micromonospora** | 17614 | 18358 | 15891 | 20013 | 12088 | 11762 | 14858 | 5732 |
| **Actinoplanes** | 9763 | 10957 | 8141 | 10992 | 6751 | 6788 | 7877 | 3184 |
| **Plantactinospora** | 2434 | 2624 | 2247 | 2586 | 1609 | 1689 | 2000 | 808 |
| **Salinispora** | 940 | 817 | 811 | 952 | 673 | 683 | 811 | 357 |
| **Verrucosispora** | 873 | 788 | 764 | 930 | 664 | 630 | 694 | 296 |
| **Actinomyces** | 13819 | 9063 | 13004 | 10705 | 7588 | 9747 | 8389 | 3062 |
| **Flaviflexus** | 1793 | 923 | 1685 | 2088 | 619 | 832 | 928 | 327 |
| **Schaalia** | 1319 | 670 | 1037 | 725 | 755 | 1208 | 821 | 223 |
| **Trueperella** | 826 | 625 | 541 | 458 | 401 | 889 | 539 | 195 |
| **Actinotignum** | 356 | 189 | 242 | 266 | 1197 | 300 | 2894 | 116 |
| **Actinobaculum** | 292 | 188 | 204 | 146 | 96 | 234 | 137 | 90 |
| **Mobiluncus** | 249 | 107 | 102 | 102 | 115 | 240 | 126 | 55 |
| **Arcanobacterium** | 241 | 176 | 129 | 125 | 103 | 267 | 145 | 65 |
| **Nakamurella** | 17670 | 27840 | 2957 | 6358 | 11649 | 7590 | 10244 | 5305 |
| **Streptosporangium** | 2825 | 2874 | 2484 | 2824 | 2014 | 2189 | 2444 | 960 |
| **Nonomuraea** | 2415 | 2451 | 2286 | 2591 | 1675 | 1651 | 1916 | 806 |
| **Nocardiopsis** | 2794 | 2632 | 2658 | 3038 | 1784 | 2118 | 2133 | 935 |
| **Streptomonospora** | 897 | 824 | 836 | 954 | 576 | 603 | 695 | 297 |
| **Thermobifida** | 420 | 416 | 328 | 355 | 267 | 320 | 341 | 133 |
| **Actinomadura** | 1258 | 1254 | 1291 | 1117 | 806 | 867 | 929 | 473 |
| **Thermomonospora** | 1047 | 964 | 873 | 862 | 689 | 798 | 798 | 403 |
| **Frankia** | 6047 | 5640 | 5128 | 6391 | 4312 | 4260 | 4852 | 2101 |
| **Jatrophihabitans** | 577 | 645 | 432 | 667 | 454 | 454 | 555 | 245 |
| **Modestobacter** | 1463 | 1624 | 1440 | 1593 | 1083 | 990 | 1261 | 528 |
| **Geodermatophilus** | 1276 | 1389 | 1165 | 1327 | 983 | 805 | 1111 | 492 |
| **Blastococcus** | 1161 | 1259 | 1088 | 1264 | 849 | 903 | 998 | 398 |
| **Jiangella** | 3731 | 3921 | 3891 | 4785 | 2611 | 2621 | 3480 | 1068 |
| **Catenulispora** | 1529 | 1494 | 1335 | 1645 | 1016 | 1088 | 1255 | 646 |
| **Kineococcus** | 1273 | 1213 | 1662 | 1478 | 857 | 1037 | 1069 | 616 |
| **Stackebrandtia** | 1055 | 980 | 917 | 1102 | 638 | 824 | 771 | 372 |
| **Thermobispora** | 803 | 766 | 838 | 803 | 549 | 491 | 643 | 212 |
| **Candidatus Planktophila** | 598 | 462 | 1034 | 201 | 160 | 598 | 347 | 2209 |
| **Candidatus Nanopelagicu** | 187 | 155 | 84 | 92 | 55 | 188 | 221 | 278 |
| **Actinopolyspora** | 428 | 476 | 409 | 465 | 305 | 336 | 385 | 162 |
| **Acidothermus** | 272 | 236 | 155 | 224 | 149 | 174 | 181 | 112 |
| **Collinsella** | 120084 | 37312 | 22677 | 19563 | 48884 | 62239 | 36977 | 10742 |
| **Coriobacterium** | 780 | 494 | 251 | 188 | 251 | 1066 | 364 | 131 |

| | | | | | | | | |
|---|---|---|---|---|---|---|---|---|
| **Olsenella** | 13792 | 7775 | 4011 | 2524 | 2620 | 14472 | 5756 | 3581 |
| **Parolsenella** | 2366 | 1781 | 1029 | 526 | 3061 | 4726 | 1172 | 464 |
| **Libanicoccus** | 1937 | 1382 | 719 | 453 | 607 | 2357 | 1126 | 394 |
| **Atopobium** | 667 | 273 | 233 | 504 | 261 | 593 | 244 | 96 |
| **Gordonibacter** | 7572 | 4840 | 2759 | 2804 | 5249 | 9903 | 4473 | 1225 |
| **Eggerthella** | 7525 | 6622 | 2879 | 5465 | 7586 | 12962 | 5956 | 1187 |
| **Adlercreutzia** | 4897 | 4089 | 1990 | 2041 | 1376 | 19378 | 3241 | 824 |
| **Slackia** | 1992 | 1240 | 731 | 530 | 640 | 2658 | 988 | 314 |
| **Denitrobacterium** | 1237 | 790 | 321 | 273 | 419 | 1335 | 567 | 155 |
| **Phoenicibacter** | 1047 | 546 | 277 | 397 | 363 | 1984 | 607 | 154 |
| **Cryptobacterium** | 260 | 151 | 77 | 73 | 74 | 307 | 120 | 80 |
| **Euzebya** | 933 | 823 | 664 | 1033 | 549 | 699 | 669 | 409 |
| **Egicoccus** | 786 | 708 | 758 | 675 | 456 | 577 | 568 | 326 |
| **Egibacter** | 618 | 519 | 416 | 632 | 378 | 457 | 438 | 282 |
| **Conexibacter** | 1598 | 1566 | 1032 | 1662 | 918 | 1232 | 1159 | 519 |
| **Rubrobacter** | 928 | 863 | 660 | 699 | 424 | 955 | 623 | 271 |
| **Acidimicrobium** | 271 | 224 | 169 | 340 | 149 | 218 | 177 | 171 |
| **Oscillatoria** | 14913 | 1180 | 4011 | 1383 | 540 | 590 | 571 | 227 |
| **Moorea** | 287 | 251 | 387 | 177 | 86 | 241 | 237 | 208 |
| **Cyanothece** | 729 | 451 | 283 | 375 | 226 | 615 | 406 | 599 |
| **Microcoleus** | 205 | 317 | 90 | 123 | 52 | 192 | 124 | 143 |
| **Arthrospira** | 165 | 136 | 102 | 57 | 55 | 152 | 110 | 302 |
| **Planktothrix** | 120 | 110 | 75 | 139 | 28 | 89 | 87 | 495 |
| **Trichodesmium** | 107 | 118 | 60 | 54 | 47 | 129 | 167 | 210 |
| **Geitlerinema** | 319 | 215 | 190 | 174 | 102 | 278 | 187 | 134 |
| **Crinalium** | 168 | 96 | 82 | 51 | 38 | 117 | 65 | 147 |
| **Geminocystis** | 451 | 537 | 657 | 341 | 122 | 405 | 632 | 657 |
| **Chondrocystis** | 199 | 122 | 94 | 120 | 39 | 142 | 201 | 90 |
| **Gloeocapsa** | 108 | 112 | 69 | 60 | 33 | 88 | 98 | 248 |
| **Microcystis** | 262 | 243 | 251 | 180 | 98 | 259 | 338 | 370 |
| **Cyanobacterium** | 177 | 106 | 62 | 92 | 24 | 133 | 140 | 206 |
| **Halothece** | 87 | 103 | 48 | 30 | 227 | 90 | 60 | 135 |
| **Candidatus Atelocyanoba** | 50 | 61 | 24 | 13 | 6 | 59 | 28 | 164 |
| **Synechococcus** | 3187 | 2359 | 1838 | 1743 | 1058 | 3488 | 1938 | 1733 |
| **Cyanobium** | 933 | 764 | 590 | 693 | 397 | 847 | 610 | 412 |
| **Thermosynechococcus** | 209 | 125 | 86 | 96 | 65 | 167 | 102 | 130 |
| **Dactylococcopsis** | 52 | 72 | 40 | 94 | 24 | 62 | 197 | 55 |
| **Leptolyngbya** | 715 | 671 | 450 | 420 | 220 | 732 | 450 | 495 |
| **Prochlorococcus** | 563 | 465 | 225 | 272 | 155 | 531 | 498 | 662 |
| **Pseudanabaena** | 273 | 236 | 119 | 107 | 73 | 288 | 217 | 240 |

| | | | | | | | | |
|---|---|---|---|---|---|---|---|---|
| **Synechocystis** | 225 | 166 | 114 | 95 | 66 | 211 | 262 | 112 |
| **Acaryochloris** | 182 | 116 | 66 | 57 | 56 | 156 | 85 | 187 |
| **Halomicronema** | 172 | 175 | 126 | 146 | 60 | 207 | 121 | 99 |
| **Chamaesiphon** | 150 | 159 | 106 | 92 | 48 | 155 | 109 | 90 |
| **Nostoc** | 1859 | 1347 | 2192 | 1277 | 522 | 1329 | 1280 | 1710 |
| **Anabaena** | 293 | 264 | 157 | 253 | 136 | 386 | 261 | 628 |
| **Trichormus** | 207 | 122 | 386 | 67 | 35 | 125 | 78 | 139 |
| **Cylindrospermum** | 149 | 126 | 109 | 58 | 178 | 204 | 151 | 79 |
| **Calothrix** | 1487 | 1136 | 1636 | 882 | 436 | 1297 | 1172 | 1345 |
| **Rivularia** | 210 | 145 | 106 | 90 | 42 | 175 | 172 | 131 |
| **Microchaete** | 87 | 108 | 672 | 38 | 26 | 77 | 77 | 182 |
| **Sphaerospermopsis** | 170 | 162 | 85 | 87 | 39 | 103 | 163 | 211 |
| **Nodularia** | 118 | 120 | 44 | 124 | 42 | 100 | 238 | 204 |
| **Anabaenopsis** | 82 | 71 | 39 | 27 | 21 | 62 | 43 | 62 |
| **Dolichospermum** | 63 | 68 | 45 | 116 | 25 | 71 | 67 | 80 |
| **Raphidiopsis** | 61 | 68 | 42 | 85 | 47 | 56 | 86 | 101 |
| **Scytonema** | 419 | 243 | 233 | 155 | 98 | 243 | 382 | 353 |
| **Fischerella** | 279 | 256 | 192 | 127 | 71 | 246 | 372 | 248 |
| **Tolypothrix** | 1 | 0 | 0 | 0 | 0 | 0 | 0 | 1 |
| **Gloeobacter** | 909 | 674 | 480 | 651 | 303 | 769 | 546 | 425 |
| **Stanieria** | 252 | 189 | 252 | 219 | 90 | 197 | 258 | 210 |
| **Pleurocapsa** | 146 | 95 | 57 | 43 | 39 | 142 | 62 | 99 |
| **Chroococcidiopsis** | 153 | 204 | 78 | 80 | 37 | 171 | 250 | 166 |
| **Gloeomargarita** | 124 | 78 | 94 | 63 | 31 | 100 | 75 | 84 |
| **Meiothermus** | 15916 | 1139 | 15692 | 1099 | 1700 | 2962 | 1073 | 359 |
| **Thermus** | 1612 | 1190 | 2043 | 1203 | 832 | 8114 | 888 | 460 |
| **Oceanithermus** | 600 | 477 | 646 | 492 | 215 | 671 | 384 | 171 |
| **Marinithermus** | 230 | 227 | 295 | 197 | 83 | 237 | 139 | 80 |
| **Deinococcus** | 9056 | 7491 | 7312 | 7641 | 3375 | 8289 | 7218 | 2874 |
| **Truepera** | 480 | 343 | 374 | 293 | 188 | 378 | 291 | 137 |
| **Mycoplasma** | 4109 | 2552 | 2945 | 1466 | 945 | 3152 | 2260 | 3426 |
| **Ureaplasma** | 69 | 97 | 41 | 30 | 21 | 90 | 63 | 67 |
| **Candidatus Hepatoplasma** | 41 | 36 | 20 | 18 | 6 | 65 | 39 | 48 |
| **Spiroplasma** | 1878 | 1016 | 556 | 508 | 371 | 1437 | 998 | 1351 |
| **Mesoplasma** | 394 | 258 | 170 | 192 | 77 | 327 | 250 | 464 |
| **Entomoplasma** | 268 | 181 | 118 | 89 | 54 | 159 | 222 | 178 |
| **Acholeplasma** | 1348 | 832 | 509 | 382 | 248 | 1143 | 885 | 676 |
| **Candidatus Phytoplasma** | 213 | 118 | 66 | 63 | 51 | 155 | 104 | 142 |
| **Candidatus Izimaplasma** | 271 | 142 | 112 | 51 | 58 | 166 | 82 | 123 |
| **Dehalococcoides** | 1237 | 708 | 357 | 215 | 172 | 1008 | 583 | 452 |

| | | | | | | | | |
|---|---|---|---|---|---|---|---|---|
| **Dehalogenimonas** | 375 | 258 | 232 | 130 | 93 | 442 | 179 | 130 |
| **Pelolinea** | 489 | 289 | 192 | 164 | 129 | 509 | 204 | 150 |
| **Anaerolinea** | 301 | 230 | 186 | 170 | 157 | 347 | 135 | 114 |
| **Brevefilum** | 222 | 126 | 866 | 64 | 70 | 317 | 122 | 90 |
| **Roseiflexus** | 917 | 688 | 604 | 497 | 311 | 772 | 488 | 316 |
| **Chloroflexus** | 396 | 270 | 239 | 205 | 143 | 352 | 178 | 211 |
| **Candidatus Promineofilum** | 905 | 616 | 483 | 658 | 524 | 881 | 539 | 285 |
| **Sphaerobacter** | 620 | 610 | 406 | 468 | 267 | 539 | 352 | 162 |
| **Thermomicrobium** | 161 | 143 | 132 | 90 | 71 | 173 | 107 | 91 |
| **Caldilinea** | 482 | 354 | 272 | 337 | 185 | 546 | 274 | 179 |
| **Fimbriimonas** | 314 | 282 | 184 | 172 | 143 | 352 | 219 | 86 |
| **Chthonomonas** | 172 | 131 | 53 | 48 | 35 | 152 | 60 | 53 |
| **Thermobaculum** | 203 | 138 | 156 | 135 | 56 | 146 | 89 | 37 |
| **Bacteroides** | 1121241 | 526894 | 316994 | 147173 | 185952 | 920958 | 176632 | 37921 |
| **Alistipes** | 147586 | 145580 | 115989 | 18115 | 67921 | 198666 | 35764 | 10874 |
| **Mucinivorans** | 753 | 804 | 374 | 185 | 160 | 712 | 572 | 150 |
| **Prevotella** | 40658 | 103659 | 27116 | 6700 | 10062 | 72625 | 17398 | 4272 |
| **Paraprevotella** | 17797 | 14049 | 6500 | 3635 | 3731 | 11438 | 3852 | 873 |
| **Parabacteroides** | 43726 | 44390 | 30886 | 13880 | 13407 | 158352 | 18329 | 6612 |
| **Tannerella** | 2704 | 3290 | 1700 | 655 | 697 | 3184 | 1264 | 352 |
| **Odoribacter** | 16654 | 17881 | 14631 | 3720 | 4250 | 24171 | 5690 | 1176 |
| **Butyricimonas** | 15366 | 15677 | 8930 | 3422 | 5663 | 26281 | 4917 | 1299 |
| **Porphyromonas** | 3256 | 3908 | 1961 | 892 | 946 | 17656 | 1448 | 450 |
| **Petrimonas** | 2705 | 1811 | 2651 | 699 | 373 | 1501 | 602 | 204 |
| **Fermentimonas** | 949 | 738 | 599 | 371 | 164 | 630 | 598 | 357 |
| **Barnesiella** | 5059 | 8831 | 3340 | 994 | 1210 | 6253 | 1478 | 662 |
| **Paludibacter** | 3398 | 1348 | 883 | 764 | 319 | 816 | 917 | 681 |
| **Muribaculum** | 1963 | 3577 | 1232 | 450 | 443 | 2118 | 612 | 182 |
| **Proteiniphilum** | 1439 | 1580 | 1082 | 469 | 298 | 1083 | 591 | 249 |
| **Candidatus Azobacteroide** | 390 | 323 | 218 | 179 | 82 | 311 | 190 | 200 |
| **Draconibacterium** | 802 | 1059 | 698 | 615 | 220 | 691 | 835 | 447 |
| **Alkalitalea** | 378 | 425 | 302 | 251 | 101 | 237 | 350 | 246 |
| **Salinivirga** | 276 | 329 | 224 | 217 | 77 | 280 | 314 | 205 |
| **Chryseobacterium** | 125535 | 146556 | 133353 | 285209 | 27541 | 58781 | 228942 | 256387 |
| **Flavobacterium** | 71969 | 167145 | 70921 | 346394 | 66037 | 58190 | 758629 | 126960 |
| **Cloacibacterium** | 69689 | 44001 | 208941 | 40793 | 16979 | 18882 | 60709 | 96479 |
| **Elizabethkingia** | 9660 | 14907 | 5709 | 10325 | 1627 | 16083 | 13900 | 10159 |
| **Polaribacter** | 5129 | 9629 | 4453 | 17973 | 2464 | 3981 | 34335 | 13932 |
| **Capnocytophaga** | 4395 | 5582 | 4704 | 8069 | 1658 | 3654 | 12931 | 5039 |
| **Myroides** | 3348 | 7251 | 2061 | 6787 | 1589 | 1642 | 19141 | 4322 |

| | | | | | | | | |
|---|---|---|---|---|---|---|---|---|
| **Tenacibaculum** | 3303 | 6497 | 3425 | 12917 | 1691 | 2719 | 24055 | 8934 |
| **Riemerella** | 2932 | 2359 | 2071 | 3100 | 651 | 1086 | 4304 | 4572 |
| **Ornithobacterium** | 2698 | 3130 | 1361 | 1204 | 595 | 3538 | 1986 | 1412 |
| **Gramella** | 1971 | 3184 | 2398 | 4783 | 761 | 1752 | 8284 | 2658 |
| **Nonlabens** | 1777 | 2805 | 2192 | 5091 | 665 | 1421 | 8958 | 2956 |
| **Maribacter** | 1692 | 3063 | 1961 | 5084 | 722 | 1578 | 9686 | 3096 |
| **Cellulophaga** | 1530 | 3214 | 1669 | 5827 | 811 | 1399 | 12655 | 3909 |
| **Winogradskyella** | 1521 | 2788 | 2009 | 6054 | 812 | 1301 | 9756 | 3964 |
| **Formosa** | 1401 | 2535 | 1392 | 5378 | 696 | 1173 | 9159 | 3052 |
| **Lacinutrix** | 1376 | 3104 | 1799 | 5156 | 774 | 1228 | 11193 | 3970 |
| **Aquimarina** | 1347 | 2438 | 1937 | 4557 | 614 | 1177 | 8584 | 3084 |
| **Salegentibacter** | 975 | 1730 | 986 | 2665 | 369 | 654 | 5142 | 1505 |
| **Lutibacter** | 959 | 1884 | 892 | 3903 | 495 | 851 | 7688 | 2874 |
| **Dokdonia** | 904 | 1441 | 809 | 2914 | 415 | 736 | 5360 | 1817 |
| **Olleya** | 782 | 1688 | 849 | 3695 | 423 | 655 | 6929 | 2306 |
| **Aequorivita** | 705 | 1349 | 848 | 2357 | 400 | 556 | 4461 | 1626 |
| **Mariniflexile** | 701 | 1679 | 884 | 3153 | 415 | 668 | 7570 | 1696 |
| **Weeksella** | 590 | 595 | 865 | 847 | 239 | 350 | 1731 | 8012 |
| **Zunongwangia** | 575 | 794 | 491 | 1409 | 194 | 416 | 2717 | 836 |
| **Kordia** | 568 | 1411 | 654 | 3013 | 322 | 589 | 4765 | 1518 |
| **Arenibacter** | 520 | 611 | 308 | 1033 | 155 | 290 | 1222 | 528 |
| **Siansivirga** | 519 | 986 | 483 | 1840 | 240 | 460 | 4187 | 1212 |
| **Seonamhaeicola** | 489 | 844 | 542 | 1995 | 283 | 464 | 3211 | 1164 |
| **Algibacter** | 489 | 931 | 420 | 1897 | 255 | 444 | 3821 | 1093 |
| **Muricauda** | 483 | 880 | 598 | 1274 | 195 | 455 | 2690 | 789 |
| **Flavivirga** | 450 | 721 | 434 | 1457 | 196 | 365 | 2179 | 838 |
| **Wenyingzhuangia** | 438 | 841 | 405 | 1701 | 209 | 365 | 2917 | 1224 |
| **Tamlana** | 433 | 848 | 418 | 1678 | 196 | 369 | 2814 | 1112 |
| **Apibacter** | 407 | 375 | 301 | 433 | 98 | 259 | 816 | 1116 |
| **Gillisia** | 385 | 575 | 249 | 1197 | 120 | 241 | 1937 | 838 |
| **Euzebyella** | 337 | 471 | 422 | 903 | 134 | 277 | 1348 | 493 |
| **Zobellia** | 326 | 442 | 323 | 608 | 104 | 209 | 844 | 333 |
| **Croceibacter** | 312 | 505 | 289 | 875 | 124 | 236 | 1442 | 791 |
| **Psychroflexus** | 290 | 451 | 299 | 873 | 121 | 232 | 1552 | 821 |
| **Robiginitalea** | 270 | 389 | 236 | 219 | 107 | 269 | 290 | 89 |
| **Flagellimonas** | 252 | 440 | 232 | 633 | 111 | 347 | 1268 | 395 |
| **Sediminicola** | 200 | 388 | 250 | 919 | 121 | 190 | 1347 | 323 |
| **Aureitalea** | 181 | 271 | 198 | 409 | 72 | 142 | 526 | 267 |
| **Gilvibacter** | 180 | 305 | 201 | 463 | 80 | 152 | 801 | 209 |
| **Muriicola** | 154 | 229 | 138 | 356 | 63 | 136 | 717 | 229 |

| | | | | | | | | |
|---|---|---|---|---|---|---|---|---|
| **Fluviicola** | 562 | 986 | 788 | 2898 | 328 | 486 | 3545 | 1192 |
| **Blattabacterium** | 485 | 470 | 330 | 855 | 152 | 361 | 1237 | 851 |
| **Owenweeksia** | 320 | 418 | 620 | 585 | 111 | 291 | 738 | 324 |
| **Candidatus Sulcia** | 109 | 97 | 48 | 116 | 21 | 38 | 84 | 177 |
| **Candidatus Walczuchella** | 15 | 10 | 19 | 26 | 5 | 22 | 27 | 22 |
| **Ichthyobacterium** | 70 | 76 | 54 | 132 | 17 | 53 | 164 | 158 |
| **Runella** | 2566 | 1732 | 962 | 1030 | 254 | 577 | 1487 | 891 |
| **Spirosoma** | 2232 | 2576 | 1329 | 1388 | 605 | 1726 | 2058 | 795 |
| **Dyadobacter** | 1401 | 1547 | 530 | 589 | 517 | 704 | 710 | 193 |
| **Fibrella** | 933 | 1194 | 712 | 579 | 468 | 930 | 623 | 371 |
| **Cytophaga** | 666 | 806 | 371 | 474 | 218 | 351 | 684 | 385 |
| **Leadbetterella** | 546 | 675 | 687 | 866 | 199 | 291 | 1546 | 521 |
| **Pseudarcicella** | 341 | 558 | 288 | 851 | 100 | 191 | 548 | 341 |
| **Arcticibacterium** | 294 | 353 | 216 | 545 | 103 | 275 | 781 | 302 |
| **Hymenobacter** | 5759 | 6828 | 3762 | 3421 | 2231 | 5836 | 3263 | 2055 |
| **Pontibacter** | 1304 | 1480 | 1213 | 704 | 323 | 1234 | 676 | 391 |
| **Rufibacter** | 1100 | 1236 | 656 | 628 | 284 | 985 | 772 | 443 |
| **Echinicola** | 632 | 730 | 466 | 844 | 195 | 601 | 1097 | 336 |
| **Cyclobacterium** | 591 | 801 | 1098 | 573 | 215 | 568 | 1865 | 651 |
| **Algoriphagus** | 576 | 679 | 382 | 859 | 185 | 558 | 1262 | 408 |
| **Belliella** | 474 | 1056 | 582 | 1554 | 308 | 545 | 5166 | 861 |
| **Aquiflexum** | 318 | 426 | 292 | 402 | 114 | 290 | 523 | 299 |
| **Flammeovirga** | 752 | 962 | 539 | 1332 | 218 | 595 | 2050 | 1301 |
| **Marivirga** | 306 | 372 | 265 | 387 | 76 | 190 | 657 | 421 |
| **Chryseolinea** | 591 | 739 | 458 | 296 | 223 | 510 | 342 | 205 |
| **Persicobacter** | 398 | 478 | 283 | 356 | 87 | 382 | 351 | 252 |
| **Bernardetia** | 351 | 588 | 401 | 581 | 132 | 296 | 1367 | 775 |
| **Candidatus Cardinium** | 122 | 82 | 325 | 33 | 22 | 84 | 100 | 53 |
| **Candidatus Amoebophilus** | 66 | 60 | 31 | 36 | 26 | 46 | 141 | 66 |
| **Sphingobacterium** | 5795 | 5867 | 2095 | 5339 | 1540 | 2236 | 10235 | 3261 |
| **Pedobacter** | 5788 | 14743 | 4638 | 5027 | 2239 | 3295 | 9142 | 3461 |
| **Mucilaginibacter** | 3256 | 4421 | 2758 | 2628 | 1039 | 2355 | 3565 | 1508 |
| **Pseudopedobacter** | 656 | 855 | 2169 | 1136 | 274 | 458 | 1092 | 536 |
| **Solitalea** | 570 | 669 | 523 | 1030 | 157 | 469 | 1193 | 621 |
| **Niabella** | 3011 | 3511 | 1921 | 1448 | 675 | 1296 | 2238 | 683 |
| **Chitinophaga** | 2044 | 3027 | 2278 | 1391 | 651 | 1241 | 2105 | 761 |
| **Niastella** | 1190 | 2089 | 1464 | 648 | 549 | 728 | 842 | 429 |
| **Arachidicoccus** | 1155 | 1713 | 989 | 1031 | 332 | 815 | 1963 | 1167 |
| **Pseudoflavitalea** | 1153 | 2363 | 1107 | 759 | 344 | 723 | 1080 | 241 |
| **Filimonas** | 902 | 1622 | 1045 | 843 | 311 | 578 | 1206 | 277 |

| | | | | | | | | |
|---|---|---|---|---|---|---|---|---|
| **Flavisolibacter** | 602 | 1033 | 683 | 413 | 170 | 392 | 529 | 258 |
| **Rhodothermus** | 842 | 897 | 509 | 588 | 307 | 931 | 482 | 283 |
| **Salinibacter** | 555 | 674 | 386 | 384 | 214 | 558 | 362 | 175 |
| **Haliscomenobacter** | 709 | 769 | 561 | 580 | 218 | 526 | 1257 | 467 |
| **Saprospira** | 149 | 165 | 779 | 151 | 63 | 125 | 117 | 126 |
| **Chlorobium** | 1070 | 840 | 524 | 522 | 282 | 1134 | 904 | 433 |
| **Pelodictyon** | 428 | 383 | 257 | 258 | 142 | 434 | 428 | 155 |
| **Chlorobaculum** | 1242 | 1071 | 708 | 625 | 368 | 1253 | 637 | 422 |
| **Prosthecochloris** | 760 | 672 | 551 | 346 | 185 | 948 | 402 | 296 |
| **Chloroherpeton** | 239 | 168 | 81 | 77 | 43 | 219 | 86 | 77 |
| **Ignavibacterium** | 336 | 209 | 379 | 171 | 70 | 247 | 345 | 182 |
| **Melioribacter** | 167 | 122 | 120 | 67 | 40 | 177 | 142 | 95 |
| **Candidatus Cyclonatronur** | 304 | 248 | 152 | 193 | 62 | 325 | 350 | 174 |
| **Gemmatirosa** | 1513 | 2095 | 1024 | 1432 | 756 | 1327 | 1035 | 413 |
| **Gemmatimonas** | 1295 | 3112 | 938 | 1054 | 545 | 838 | 1110 | 420 |
| **Fibrobacter** | 460 | 407 | 203 | 156 | 124 | 590 | 245 | 105 |
| **Candidatus Cloacimonas** | 193 | 152 | 72 | 45 | 42 | 170 | 78 | 252 |
| **Akkermansia** | 175455 | 101738 | 30092 | 34977 | 53754 | 204761 | 104285 | 32366 |
| **Verrucomicrobium** | 1624 | 1176 | 803 | 722 | 555 | 1186 | 673 | 537 |
| **Opitutus** | 2054 | 1747 | 990 | 1179 | 817 | 1416 | 970 | 424 |
| **Lacunisphaera** | 1007 | 625 | 595 | 484 | 381 | 628 | 373 | 250 |
| **Ereboglobus** | 676 | 473 | 314 | 303 | 194 | 595 | 282 | 185 |
| **Coraliomargarita** | 138 | 140 | 58 | 62 | 35 | 117 | 77 | 52 |
| **Methylacidiphilum** | 196 | 98 | 64 | 67 | 34 | 130 | 99 | 47 |
| **Candidatus Xiphinematob** | 51 | 36 | 18 | 12 | 13 | 50 | 20 | 16 |
| **Planctomyces** | 2293 | 2157 | 1104 | 1474 | 1630 | 1474 | 1371 | 590 |
| **Pirellula** | 367 | 260 | 245 | 168 | 261 | 233 | 146 | 97 |
| **Fuerstia** | 284 | 194 | 632 | 137 | 117 | 205 | 153 | 131 |
| **Rubinisphaera** | 274 | 204 | 158 | 121 | 141 | 276 | 150 | 180 |
| **Thermogutta** | 239 | 186 | 121 | 132 | 76 | 261 | 101 | 80 |
| **Rhodopirellula** | 210 | 197 | 131 | 245 | 119 | 197 | 122 | 110 |
| **Planctopirus** | 199 | 165 | 106 | 74 | 95 | 152 | 110 | 112 |
| **Paludisphaera** | 1364 | 1045 | 654 | 862 | 821 | 819 | 818 | 306 |
| **Singulisphaera** | 647 | 551 | 350 | 378 | 431 | 338 | 347 | 169 |
| **Isosphaera** | 278 | 212 | 207 | 182 | 167 | 235 | 187 | 105 |
| **Gemmata** | 2068 | 1392 | 1227 | 1004 | 1409 | 1300 | 1002 | 675 |
| **Phycisphaera** | 762 | 681 | 506 | 519 | 361 | 677 | 407 | 209 |
| **Sedimentisphaera** | 316 | 197 | 118 | 87 | 70 | 341 | 302 | 143 |
| **Candidatus Kuenenia** | 159 | 186 | 82 | 75 | 48 | 154 | 73 | 103 |
| **Candidatus Protochlamyd** | 1024 | 301 | 342 | 61 | 154 | 201 | 131 | 128 |

| | | | | | | | | |
|---|---|---|---|---|---|---|---|---|
| **Parachlamydia** | 94 | 44 | 22 | 81 | 40 | 47 | 49 | 36 |
| **Neochlamydia** | 64 | 37 | 30 | 14 | 13 | 27 | 32 | 24 |
| **Simkania** | 196 | 51 | 23 | 99 | 44 | 80 | 79 | 53 |
| **Waddlia** | 64 | 45 | 38 | 34 | 22 | 54 | 49 | 98 |
| **Chlamydia** | 692 | 354 | 571 | 300 | 200 | 521 | 369 | 573 |
| **Kiritimatiella** | 416 | 363 | 248 | 238 | 158 | 510 | 249 | 149 |
| **Fusobacterium** | 22106 | 3325 | 2528 | 1758 | 1437 | 5676 | 4060 | 5941 |
| **Ilyobacter** | 254 | 212 | 96 | 58 | 57 | 271 | 104 | 215 |
| **Sebaldella** | 3088 | 338 | 166 | 163 | 98 | 529 | 246 | 7971 |
| **Leptotrichia** | 1091 | 756 | 390 | 334 | 244 | 1099 | 536 | 5988 |
| **Sneathia** | 369 | 233 | 70 | 120 | 292 | 340 | 227 | 798 |
| **Streptobacillus** | 263 | 216 | 68 | 139 | 64 | 224 | 218 | 1345 |
| **Treponema** | 6149 | 2703 | 1745 | 1592 | 916 | 5707 | 2218 | 1180 |
| **Sphaerochaeta** | 1090 | 530 | 358 | 288 | 204 | 1270 | 450 | 269 |
| **Spirochaeta** | 716 | 521 | 300 | 311 | 172 | 723 | 348 | 140 |
| **Salinispira** | 272 | 228 | 86 | 64 | 53 | 310 | 104 | 56 |
| **Sediminispirochaeta** | 272 | 151 | 125 | 122 | 81 | 376 | 151 | 69 |
| **Borrelia** | 410 | 503 | 139 | 224 | 115 | 319 | 327 | 392 |
| **Borreliella** | 318 | 190 | 98 | 124 | 74 | 270 | 154 | 314 |
| **Brachyspira** | 1287 | 885 | 455 | 517 | 283 | 1348 | 981 | 975 |
| **Leptospira** | 862 | 864 | 625 | 818 | 292 | 884 | 794 | 810 |
| **Turneriella** | 242 | 138 | 260 | 97 | 70 | 206 | 198 | 87 |
| **Cloacibacillus** | 3841 | 1114 | 691 | 415 | 744 | 8452 | 1671 | 325 |
| **Aminomonas** | 1058 | 335 | 212 | 263 | 141 | 663 | 313 | 156 |
| **Jonquetella** | 489 | 229 | 143 | 136 | 151 | 458 | 244 | 51 |
| **Thermanaerovibrio** | 316 | 225 | 156 | 150 | 100 | 551 | 177 | 84 |
| **Acetomicrobium** | 245 | 93 | 80 | 70 | 28 | 154 | 84 | 40 |
| **Aminobacterium** | 163 | 62 | 58 | 37 | 31 | 128 | 107 | 43 |
| **Thermovirga** | 108 | 55 | 27 | 33 | 23 | 103 | 35 | 36 |
| **Terriglobus** | 966 | 828 | 563 | 634 | 376 | 974 | 590 | 299 |
| **Granulicella** | 863 | 670 | 887 | 529 | 301 | 753 | 518 | 271 |
| **Acidobacterium** | 552 | 488 | 382 | 324 | 205 | 512 | 339 | 170 |
| **Candidatus Koribacter** | 454 | 395 | 307 | 313 | 167 | 443 | 240 | 140 |
| **Candidatus Solibacter** | 844 | 825 | 613 | 528 | 378 | 765 | 491 | 262 |
| **Luteitalea** | 1326 | 2013 | 946 | 1201 | 1374 | 1099 | 1159 | 462 |
| **Chloracidobacterium** | 326 | 292 | 176 | 210 | 143 | 299 | 204 | 129 |
| **Fervidobacterium** | 609 | 356 | 424 | 216 | 346 | 545 | 226 | 165 |
| **Thermosipho** | 280 | 201 | 237 | 110 | 89 | 319 | 282 | 209 |
| **Thermotoga** | 489 | 320 | 182 | 140 | 306 | 446 | 347 | 153 |
| **Pseudothermotoga** | 301 | 160 | 79 | 184 | 89 | 267 | 87 | 86 |

| | | | | | | | | |
|---|---|---|---|---|---|---|---|---|
| **Marinitoga** | 184 | 173 | 83 | 48 | 40 | 225 | 86 | 178 |
| **Defluviitoga** | 137 | 106 | 3054 | 59 | 39 | 123 | 122 | 84 |
| **Petrotoga** | 122 | 83 | 60 | 32 | 30 | 165 | 66 | 84 |
| **Mesotoga** | 192 | 72 | 62 | 42 | 29 | 179 | 57 | 23 |
| **Kosmotoga** | 181 | 79 | 92 | 27 | 55 | 160 | 92 | 129 |
| **Candidatus Saccharimona** | 621 | 514 | 623 | 809 | 502 | 449 | 525 | 120 |
| **Candidatus Babela** | 91 | 54 | 22 | 13 | 20 | 42 | 24 | 58 |
| **Candidatus Bipolaricaulis** | 118 | 124 | 87 | 82 | 52 | 146 | 90 | 44 |
| **Vampirococcus** | 187 | 107 | 93 | 73 | 33 | 221 | 87 | 49 |
| **Nitrospira** | 1392 | 1742 | 5257 | 1374 | 630 | 1361 | 895 | 555 |
| **Leptospirillum** | 181 | 174 | 100 | 126 | 69 | 213 | 91 | 136 |
| **Thermodesulfovibrio** | 134 | 90 | 102 | 45 | 29 | 139 | 62 | 63 |
| **Thermocrinis** | 177 | 66 | 41 | 35 | 28 | 148 | 47 | 45 |
| **Hydrogenobaculum** | 100 | 85 | 15 | 40 | 15 | 80 | 58 | 125 |
| **Aquifex** | 86 | 86 | 26 | 24 | 8 | 111 | 38 | 23 |
| **Hydrogenobacter** | 32 | 26 | 20 | 6 | 13 | 34 | 17 | 26 |
| **Sulfurihydrogenibium** | 205 | 187 | 117 | 97 | 122 | 214 | 274 | 138 |
| **Persephonella** | 56 | 48 | 14 | 11 | 23 | 81 | 44 | 34 |
| **Thermosulfidibacter** | 126 | 114 | 66 | 96 | 16 | 115 | 61 | 22 |
| **Desulfurobacterium** | 171 | 72 | 46 | 48 | 26 | 148 | 45 | 89 |
| **Thermovibrio** | 76 | 57 | 72 | 25 | 14 | 113 | 36 | 28 |
| **Geovibrio** | 256 | 185 | 106 | 104 | 58 | 357 | 156 | 67 |
| **Calditerrivibrio** | 233 | 234 | 109 | 57 | 67 | 222 | 104 | 63 |
| **Denitrovibrio** | 166 | 117 | 40 | 30 | 42 | 187 | 71 | 72 |
| **Deferribacter** | 146 | 103 | 71 | 59 | 41 | 171 | 66 | 259 |
| **Flexistipes** | 96 | 211 | 487 | 37 | 22 | 106 | 78 | 85 |
| **Endomicrobium** | 249 | 127 | 58 | 66 | 51 | 204 | 96 | 52 |
| **Elusimicrobium** | 103 | 64 | 43 | 30 | 35 | 104 | 59 | 35 |
| **Thermodesulfobacterium** | 162 | 116 | 61 | 219 | 32 | 100 | 188 | 126 |
| **Thermodesulfatator** | 108 | 118 | 53 | 33 | 20 | 124 | 63 | 50 |
| **Caldimicrobium** | 71 | 31 | 64 | 19 | 19 | 40 | 37 | 50 |
| **Desulfurispirillum** | 334 | 250 | 126 | 207 | 79 | 290 | 154 | 117 |
| **Caldithrix** | 308 | 255 | 214 | 108 | 54 | 256 | 169 | 110 |
| **Dictyoglomus** | 247 | 153 | 130 | 171 | 51 | 274 | 223 | 165 |
| **Caldisericum** | 125 | 40 | 28 | 17 | 17 | 74 | 44 | 72 |
| **Coprothermobacter** | 62 | 18 | 21 | 24 | 7 | 42 | 11 | 17 |
| **Homo** | 1861572 | 1839757 | 197372 | 768172 | 10254514 | 1374974 | 623585 | 154989 |
| **Halorubrum** | 1112 | 1080 | 870 | 1197 | 784 | 869 | 847 | 334 |
| **Salinigranum** | 284 | 274 | 200 | 227 | 159 | 226 | 214 | 96 |
| **Halopenitus** | 197 | 175 | 145 | 193 | 120 | 196 | 143 | 52 |

| | | | | | | | | |
|---|---|---|---|---|---|---|---|---|
| **Halohasta** | 83 | 105 | 81 | 94 | 46 | 84 | 60 | 41 |
| **Haloferax** | 582 | 552 | 497 | 559 | 278 | 537 | 374 | 163 |
| **Haloplanus** | 458 | 405 | 281 | 572 | 277 | 347 | 346 | 168 |
| **Halogeometricum** | 95 | 78 | 49 | 92 | 42 | 80 | 49 | 28 |
| **Haloquadratum** | 95 | 54 | 28 | 19 | 20 | 37 | 21 | 35 |
| **Haloarcula** | 439 | 375 | 321 | 295 | 204 | 389 | 259 | 164 |
| **Natronomonas** | 397 | 264 | 167 | 269 | 160 | 230 | 208 | 121 |
| **Halorhabdus** | 282 | 268 | 185 | 308 | 158 | 214 | 212 | 85 |
| **Halomicrobium** | 220 | 159 | 153 | 191 | 127 | 192 | 145 | 55 |
| **Halorientalis** | 184 | 191 | 169 | 261 | 172 | 209 | 159 | 71 |
| **Halobacterium** | 692 | 613 | 581 | 632 | 379 | 509 | 443 | 205 |
| **Halalkalicoccus** | 211 | 168 | 127 | 215 | 87 | 153 | 111 | 48 |
| **Halorussus** | 175 | 151 | 180 | 132 | 93 | 139 | 116 | 63 |
| **Halodesulfurarchaeum** | 130 | 49 | 86 | 74 | 40 | 114 | 63 | 40 |
| **Halanaeroarchaeum** | 68 | 79 | 61 | 54 | 33 | 43 | 64 | 27 |
| **Haloterrigena** | 409 | 404 | 274 | 486 | 237 | 276 | 341 | 152 |
| **Natronolimnobius** | 353 | 397 | 317 | 299 | 213 | 293 | 312 | 131 |
| **Natrinema** | 344 | 331 | 256 | 310 | 241 | 264 | 303 | 119 |
| **Natronococcus** | 230 | 209 | 216 | 284 | 163 | 135 | 147 | 73 |
| **Halopiger** | 220 | 283 | 152 | 291 | 137 | 168 | 173 | 66 |
| **Salinarchaeum** | 202 | 158 | 147 | 157 | 119 | 180 | 162 | 52 |
| **Halobiforma** | 200 | 185 | 215 | 341 | 166 | 203 | 188 | 81 |
| **Natronobacterium** | 178 | 110 | 96 | 109 | 80 | 86 | 86 | 36 |
| **Natrialba** | 145 | 117 | 80 | 87 | 81 | 113 | 110 | 44 |
| **Halostagnicola** | 144 | 129 | 146 | 100 | 73 | 147 | 96 | 53 |
| **Methanosarcina** | 1073 | 812 | 475 | 454 | 331 | 1242 | 850 | 1159 |
| **Methanococcoides** | 148 | 89 | 72 | 59 | 48 | 133 | 66 | 35 |
| **Methanohalophilus** | 140 | 67 | 55 | 45 | 40 | 142 | 59 | 42 |
| **Methanolobus** | 76 | 57 | 29 | 25 | 27 | 77 | 41 | 26 |
| **Methanomethylovorans** | 58 | 75 | 27 | 56 | 16 | 70 | 57 | 35 |
| **Methanosalsum** | 58 | 40 | 26 | 16 | 6 | 48 | 38 | 44 |
| **Methanohalobium** | 41 | 23 | 18 | 13 | 16 | 42 | 19 | 9 |
| **Methanothrix** | 362 | 185 | 107 | 212 | 614 | 356 | 124 | 942 |
| **Methanoculleus** | 403 | 635 | 278 | 273 | 301 | 415 | 209 | 107 |
| **Methanofollis** | 98 | 103 | 80 | 73 | 75 | 115 | 109 | 32 |
| **Methanolacinia** | 94 | 105 | 37 | 46 | 28 | 142 | 54 | 14 |
| **Methanoplanus** | 86 | 51 | 59 | 29 | 12 | 72 | 91 | 26 |
| **Methanoregula** | 225 | 114 | 119 | 101 | 155 | 254 | 101 | 92 |
| **Methanosphaerula** | 76 | 95 | 51 | 35 | 36 | 102 | 52 | 327 |
| **Methanolinea** | 71 | 38 | 28 | 62 | 33 | 57 | 23 | 14 |

| | | | | | | | | |
|---|---|---|---|---|---|---|---|---|
| **Methanocorpusculum** | 124 | 73 | 52 | 90 | 17 | 172 | 67 | 48 |
| **Methanospirillum** | 49 | 66 | 14 | 50 | 43 | 40 | 27 | 34 |
| **Methanocella** | 386 | 273 | 194 | 161 | 172 | 347 | 223 | 102 |
| **Methanobrevibacter** | 2632 | 4260 | 2937 | 2841 | 9933 | 25590 | 37895 | 7644 |
| **Methanobacterium** | 571 | 348 | 242 | 426 | 727 | 844 | 466 | 784 |
| **Methanosphaera** | 176 | 128 | 51 | 60 | 44 | 238 | 182 | 130 |
| **Methanothermobacter** | 105 | 82 | 33 | 85 | 43 | 141 | 58 | 79 |
| **Methanothermus** | 37 | 17 | 12 | 8 | 7 | 38 | 20 | 21 |
| **Methanococcus** | 422 | 279 | 385 | 157 | 105 | 460 | 415 | 382 |
| **Methanothermococcus** | 55 | 21 | 11 | 13 | 12 | 31 | 38 | 19 |
| **Methanocaldococcus** | 208 | 152 | 69 | 180 | 53 | 168 | 312 | 234 |
| **Methanotorris** | 29 | 18 | 13 | 6 | 9 | 38 | 25 | 61 |
| **Thermococcus** | 1136 | 934 | 576 | 432 | 330 | 1428 | 551 | 400 |
| **Pyrococcus** | 137 | 80 | 64 | 68 | 45 | 157 | 80 | 83 |
| **Palaeococcus** | 39 | 35 | 13 | 23 | 7 | 37 | 25 | 28 |
| **Methanomassiliicoccus** | 393 | 126 | 25 | 22 | 33 | 253 | 162 | 9 |
| **Candidatus Methanoplasn** | 30 | 16 | 14 | 13 | 11 | 32 | 12 | 5 |
| **Candidatus Methanometh** | 69 | 95 | 43 | 42 | 17 | 160 | 52 | 11 |
| **Picrophilus** | 46 | 24 | 15 | 14 | 12 | 33 | 44 | 13 |
| **Thermoplasma** | 34 | 23 | 16 | 11 | 9 | 23 | 14 | 28 |
| **Cuniculiplasma** | 29 | 44 | 11 | 14 | 6 | 50 | 22 | 49 |
| **Ferroplasma** | 14 | 15 | 41 | 8 | 11 | 95 | 9 | 20 |
| **Aciduliprofundum** | 57 | 34 | 20 | 19 | 15 | 61 | 75 | 22 |
| **Archaeoglobus** | 103 | 69 | 43 | 46 | 34 | 145 | 60 | 57 |
| **Geoglobus** | 60 | 50 | 48 | 57 | 27 | 79 | 24 | 20 |
| **Ferroglobus** | 22 | 24 | 7 | 8 | 4 | 19 | 9 | 5 |
| **Methanopyrus** | 22 | 30 | 12 | 51 | 10 | 33 | 10 | 6 |
| **Sulfolobus** | 107 | 74 | 69 | 65 | 28 | 91 | 61 | 123 |
| **Sulfodiicoccus** | 81 | 31 | 11 | 16 | 11 | 17 | 42 | 8 |
| **Acidianus** | 77 | 91 | 28 | 48 | 34 | 86 | 97 | 81 |
| **Metallosphaera** | 55 | 37 | 45 | 16 | 13 | 56 | 29 | 26 |
| **Sulfurisphaera** | 49 | 40 | 12 | 19 | 9 | 51 | 28 | 38 |
| **Saccharolobus** | 18 | 20 | 19 | 13 | 6 | 36 | 8 | 23 |
| **Pyrobaculum** | 75 | 50 | 27 | 94 | 26 | 83 | 43 | 36 |
| **Thermoproteus** | 55 | 39 | 47 | 25 | 14 | 47 | 38 | 26 |
| **Vulcanisaeta** | 36 | 17 | 2 | 14 | 9 | 38 | 14 | 50 |
| **Caldivirga** | 21 | 14 | 15 | 5 | 1 | 9 | 2 | 6 |
| **Thermofilum** | 71 | 64 | 266 | 35 | 40 | 74 | 35 | 32 |
| **Staphylothermus** | 31 | 18 | 8 | 3 | 3 | 23 | 12 | 19 |
| **Desulfurococcus** | 27 | 21 | 12 | 3 | 4 | 23 | 21 | 6 |

| | | | | | | | | |
|---|---|---|---|---|---|---|---|---|
| **Ignicoccus** | 19 | 15 | 10 | 13 | 7 | 35 | 19 | 8 |
| **Thermogladius** | 12 | 13 | 11 | 12 | 3 | 12 | 12 | 23 |
| **Thermosphaera** | 6 | 7 | 1 | 7 | 0 | 6 | 3 | 11 |
| **Pyrodictium** | 38 | 21 | 11 | 11 | 13 | 27 | 15 | 9 |
| **Pyrolobus** | 25 | 26 | 14 | 8 | 2 | 20 | 4 | 3 |
| **Hyperthermus** | 18 | 9 | 51 | 11 | 3 | 22 | 4 | 32 |
| **Acidilobus** | 46 | 61 | 22 | 31 | 24 | 58 | 41 | 19 |
| **Caldisphaera** | 22 | 18 | 11 | 14 | 14 | 22 | 11 | 51 |
| **Candidatus Nitrosotenuis** | 135 | 91 | 33 | 25 | 24 | 52 | 33 | 17 |
| **Candidatus Nitrosopelagic** | 24 | 10 | 18 | 2 | 7 | 32 | 12 | 44 |
| **Nitrosopumilus** | 54 | 51 | 16 | 31 | 17 | 62 | 28 | 83 |
| **Candidatus Nitrosomarinu** | 22 | 23 | 15 | 15 | 11 | 21 | 19 | 49 |
| **Nitrososphaera** | 58 | 43 | 59 | 18 | 36 | 52 | 28 | 21 |
| **Candidatus Nitrosocaldus** | 5 | 10 | 6 | 21 | 1 | 8 | 2 | 7 |
| **Candidatus Korarchaeum** | 10 | 14 | 15 | 5 | 1 | 10 | 6 | 5 |
| **Candidatus Mancarchaeu** | 8 | 16 | 11 | 8 | 4 | 18 | 14 | 10 |
| **Vequintavirus** | 21943 | 962 | 218 | 206 | 428 | 388 | 352 | 166 |
| **Certrevirus** | 6 | 2 | 4 | 5 | 1 | 4 | 6 | 4 |
| **Seunavirus** | 6 | 5 | 5 | 0 | 12 | 3 | 4 | 13 |
| **Felixounavirus** | 3121 | 273 | 116 | 4181 | 1398 | 210 | 407 | 72 |
| **Mooglevirus** | 309 | 3 | 4 | 37 | 3 | 10 | 6 | 4 |
| **Suspvirus** | 29 | 3 | 1 | 5 | 1 | 1 | 3 | 0 |
| **Kolesnikvirus** | 3 | 10 | 7 | 6 | 6 | 7 | 9 | 1 |
| **Asteriusvirus** | 711 | 13 | 5 | 12 | 24 | 16 | 20 | 34 |
| **Bixzunavirus** | 353 | 36 | 11 | 19 | 27 | 51 | 16 | 7 |
| **Obolenskvirus** | 230 | 53 | 82 | 117 | 460 | 92 | 146 | 118 |
| **Tequatrovirus** | 24 | 273 | 11 | 158 | 11 | 300 | 36 | 83 |
| **Mosigvirus** | 7 | 11 | 1 | 0 | 2 | 73 | 222 | 14 |
| **Dhakavirus** | 7 | 1 | 2 | 1 | 7 | 70 | 5 | 4 |
| **Schizotequatrovirus** | 6 | 4 | 10 | 3 | 2 | 13 | 6 | 5 |
| **Gaprivervirus** | 5 | 7 | 0 | 0 | 0 | 12 | 2 | 6 |
| **Jiaodavirus** | 4 | 1 | 0 | 1 | 1 | 29 | 10 | 8 |
| **Slopekvirus** | 2 | 1 | 1 | 1 | 0 | 0 | 5 | 17 |
| **Karamvirus** | 1 | 1 | 0 | 1 | 1 | 3 | 0 | 7 |
| **Moonvirus** | 1 | 1 | 2 | 0 | 2 | 36 | 3 | 6 |
| **Gelderlandvirus** | 1 | 2 | 3 | 47 | 1 | 21 | 6 | 13 |
| **Pbunavirus** | 160 | 16 | 2 | 25 | 6 | 26 | 6 | 8 |
| **Mieseafarmvirus** | 36 | 10 | 3 | 21 | 110 | 16 | 51 | 24 |
| **Peduovirus** | 20 | 34 | 7 | 25 | 12 | 17 | 4 | 18 |
| **Hpunavirus** | 11 | 4 | 10 | 5 | 3 | 13 | 8 | 76 |

| | | | | | | | | |
|---|---|---|---|---|---|---|---|---|
| **Muvirus** | 30 | 0 | 0 | 0 | 1 | 1 | 29 | 0 |
| **Eneladusvirus** | 21 | 3 | 6 | 8 | 3 | 58 | 8 | 17 |
| **Chiangmaivirus** | 20 | 15 | 12 | 7 | 2 | 35 | 11 | 6 |
| **Siminovitchvirus** | 17 | 0 | 1 | 0 | 0 | 1 | 0 | 4 |
| **Phikzvirus** | 13 | 22 | 3 | 4 | 9 | 9 | 19 | 13 |
| **Tegunavirus** | 12 | 4 | 1 | 4 | 2 | 41 | 8 | 18 |
| **Alcyoneusvirus** | 9 | 4 | 2 | 1 | 3 | 4 | 5 | 8 |
| **Agricanvirus** | 9 | 11 | 11 | 2 | 6 | 17 | 9 | 0 |
| **Fletchervirus** | 6 | 2 | 0 | 0 | 2 | 1 | 1 | 11 |
| **Firehammervirus** | 3 | 2 | 1 | 1 | 0 | 5 | 1 | 13 |
| **Sepunavirus** | 8 | 3 | 0 | 4 | 11 | 3 | 4 | 6 |
| **Nazgulvirus** | 8 | 4 | 2 | 8 | 1 | 0 | 0 | 1 |
| **Pakpunavirus** | 6 | 0 | 4 | 4 | 4 | 2 | 3 | 2 |
| **Tulanevirus** | 5 | 4 | 1 | 4 | 1 | 67 | 6 | 22 |
| **Shalavirus** | 5 | 2 | 1 | 0 | 1 | 3 | 0 | 0 |
| **Jilinvirus** | 4 | 2 | 6 | 6 | 4 | 3 | 4 | 9 |
| **Otagovirus** | 4 | 9 | 6 | 3 | 4 | 9 | 7 | 4 |
| **Seoulvirus** | 4 | 2 | 6 | 1 | 0 | 3 | 0 | 3 |
| **Biquartavirus** | 3 | 0 | 7 | 2 | 2 | 10 | 1 | 2 |
| **Vidavervirus** | 3 | 7 | 3 | 6 | 8 | 3 | 5 | 1 |
| **Hapunavirus** | 3 | 7 | 0 | 0 | 3 | 1 | 4 | 8 |
| **Elvirus** | 3 | 0 | 0 | 0 | 0 | 0 | 0 | 0 |
| **Yokohamavirus** | 3 | 2 | 0 | 0 | 0 | 1 | 0 | 2 |
| **Nankokuvirus** | 3 | 0 | 1 | 0 | 1 | 0 | 0 | 1 |
| **Emdodecavirus** | 2 | 4 | 0 | 4 | 5 | 2 | 1 | 1 |
| **Radnorvirus** | 2 | 2 | 3 | 0 | 1 | 2 | 2 | 4 |
| **Erskinevirus** | 2 | 8 | 0 | 3 | 1 | 1 | 0 | 9 |
| **Vhmlvirus** | 2 | 0 | 1 | 7 | 6 | 0 | 13 | 7 |
| **Bcepmuvirus** | 2 | 1 | 1 | 0 | 2 | 3 | 2 | 2 |
| **Bequatrovirus** | 1 | 6 | 0 | 1 | 3 | 3 | 1 | 2 |
| **Viunavirus** | 1 | 1 | 0 | 0 | 4 | 1 | 3 | 2 |
| **Machinavirus** | 1 | 16 | 4 | 0 | 0 | 4 | 1 | 72 |
| **Svunavirus** | 1 | 2 | 0 | 5 | 3 | 2 | 0 | 0 |
| **Skunavirus** | 1714 | 1140 | 87 | 72 | 95 | 366 | 243 | 77 |
| **Cronusvirus** | 1150 | 23 | 11 | 18 | 9 | 14 | 24 | 5 |
| **Barnyardvirus** | 649 | 19 | 11 | 10 | 70 | 23 | 158 | 5 |
| **Liefievirus** | 378 | 517 | 52 | 94 | 865 | 165 | 268 | 73 |
| **Coopervirus** | 120 | 162 | 243 | 30 | 35 | 73 | 150 | 29 |
| **Acadianvirus** | 20 | 29 | 11 | 6 | 17 | 11 | 26 | 3 |
| **Pipefishvirus** | 16 | 25 | 12 | 6 | 7 | 16 | 12 | 9 |

| | | | | | | | | |
|---|---|---|---|---|---|---|---|---|
| **Rosebushvirus** | 16 | 14 | 6 | 9 | 6 | 34 | 24 | 7 |
| **Pegunavirus** | 13 | 22 | 5 | 2 | 10 | 7 | 12 | 9 |
| **Kagunavirus** | 139 | 7 | 7 | 4 | 68 | 5 | 3 | 32 |
| **Jerseyvirus** | 47 | 7 | 3 | 9 | 65 | 97 | 10 | 21 |
| **Cornellvirus** | 11 | 0 | 0 | 0 | 2 | 1 | 2 | 2 |
| **Ceduovirus** | 197 | 159 | 127 | 116 | 51 | 179 | 234 | 28 |
| **Fromanvirus** | 178 | 109 | 61 | 79 | 106 | 98 | 114 | 78 |
| **Pamexvirus** | 177 | 512 | 42 | 53 | 82 | 26 | 14 | 20 |
| **Septimatrevirus** | 142 | 17 | 8 | 656 | 10 | 21 | 50 | 211 |
| **Efquatrovirus** | 141 | 31 | 21 | 23 | 741 | 27 | 39 | 2 |
| **Trigintaduovirus** | 138 | 25 | 2 | 3 | 158 | 53 | 24 | 3 |
| **Phicbkvirus** | 95 | 53 | 27 | 29 | 19 | 25 | 28 | 12 |
| **Timquatrovirus** | 93 | 46 | 50 | 47 | 41 | 54 | 64 | 38 |
| **Cheoctovirus** | 77 | 50 | 88 | 60 | 35 | 29 | 40 | 19 |
| **Bronvirus** | 69 | 22 | 24 | 8 | 16 | 46 | 21 | 11 |
| **Moineauvirus** | 67 | 54 | 4 | 14 | 104 | 119 | 28 | 9 |
| **Roufvirus** | 52 | 113 | 75 | 87 | 14488 | 190 | 134 | 72 |
| **Cequinquevirus** | 40 | 11 | 2 | 8 | 2 | 48 | 16 | 3 |
| **Patiencevirus** | 39 | 7 | 2 | 47 | 8 | 13 | 33 | 3 |
| **Wizardvirus** | 37 | 5 | 1 | 12 | 10 | 7 | 1 | 0 |
| **Brussowvirus** | 35 | 37 | 21 | 4 | 173 | 29 | 47 | 10 |
| **Rtpvirus** | 10 | 14 | 6 | 22 | 54 | 27 | 29 | 6 |
| **Rogunavirus** | 9 | 5 | 1 | 2 | 22 | 8 | 5 | 2 |
| **Tlsvirus** | 6 | 1 | 0 | 3 | 8 | 9 | 24 | 6 |
| **Webervirus** | 5 | 6 | 2 | 2 | 10 | 12 | 9 | 7 |
| **Buttersvirus** | 17 | 4 | 2 | 6 | 7 | 9 | 5 | 1 |
| **Charlievirus** | 6 | 7 | 0 | 0 | 3 | 2 | 4 | 1 |
| **Redivirus** | 4 | 6 | 0 | 1 | 7 | 4 | 1 | 0 |
| **Mapvirus** | 30 | 18 | 4 | 2 | 12 | 16 | 13 | 5 |
| **Smoothievirus** | 27 | 2 | 2 | 2 | 2 | 5 | 4 | 1 |
| **Chivirus** | 25 | 10 | 10 | 33 | 3 | 12 | 10 | 12 |
| **Phayoncevirus** | 19 | 5 | 1 | 2 | 7 | 1 | 6 | 4 |
| **Fishburnevirus** | 3 | 6 | 3 | 2 | 4 | 6 | 3 | 1 |
| **Nymphadoravirus** | 16 | 8 | 1 | 3 | 4 | 4 | 17 | 0 |
| **Baxtervirus** | 6 | 5 | 1 | 0 | 4 | 2 | 2 | 1 |
| **Pepyhexavirus** | 21 | 94 | 4 | 42 | 10 | 19 | 61 | 12 |
| **Tinduovirus** | 21 | 3 | 4 | 11 | 54 | 38 | 11 | 2 |
| **Abidjanvirus** | 18 | 4 | 14 | 3 | 4 | 11 | 4 | 2 |
| **Phietavirus** | 18 | 2 | 1 | 13 | 2 | 2 | 5 | 0 |
| **Nipunavirus** | 16 | 7 | 38 | 3 | 7 | 5 | 3 | 10 |

| | | | | | | | | |
|---|---|---|---|---|---|---|---|---|
| **Chenonavirus** | 15 | 6 | 1 | 7 | 14 | 6 | 5 | 0 |
| **Nonagvirus** | 15 | 4 | 0 | 30 | 2 | 5 | 0 | 5 |
| **Likavirus** | 12 | 6 | 17 | 9 | 4 | 3 | 19 | 5 |
| **Camvirus** | 2 | 6 | 10 | 4 | 1 | 2 | 9 | 0 |
| **Arequatrovirus** | 1 | 1 | 0 | 1 | 1 | 1 | 1 | 1 |
| **Pahexavirus** | 14 | 4 | 5 | 6 | 5 | 3 | 5 | 2 |
| **Corndogvirus** | 14 | 7 | 2 | 6 | 29 | 1 | 17 | 14 |
| **Seuratvirus** | 13 | 5 | 7 | 2 | 2 | 24 | 9 | 3 |
| **Emalynvirus** | 13 | 0 | 1 | 7 | 6 | 4 | 4 | 0 |
| **Yuavirus** | 12 | 6 | 3 | 10 | 40 | 1 | 8 | 3 |
| **Biseptimavirus** | 12 | 1 | 0 | 0 | 0 | 2 | 2 | 0 |
| **Minunavirus** | 12 | 20 | 4 | 39 | 9 | 4 | 19 | 1 |
| **Betterkatzvirus** | 11 | 25 | 5 | 7 | 5 | 4 | 2 | 0 |
| **Tequintavirus** | 11 | 11 | 1 | 35 | 6 | 23 | 406 | 108 |
| **Brujitavirus** | 11 | 7 | 1 | 6 | 14 | 2 | 8 | 1 |
| **Phifelvirus** | 10 | 0 | 1 | 16 | 10 | 11 | 4 | 0 |
| **Vividuovirus** | 10 | 6 | 1 | 2 | 1 | 4 | 3 | 2 |
| **Woodruffvirus** | 8 | 9 | 1 | 9 | 4 | 2 | 6 | 4 |
| **Kostyavirus** | 8 | 3 | 3 | 2 | 4 | 3 | 3 | 3 |
| **Casadabanvirus** | 7 | 6 | 3 | 2 | 0 | 1 | 3 | 0 |
| **Gordtnkvirus** | 7 | 6 | 2 | 2 | 1 | 7 | 0 | 0 |
| **Sextaecvirus** | 7 | 0 | 0 | 3 | 4 | 4 | 11 | 4 |
| **Omegavirus** | 6 | 4 | 10 | 8 | 7 | 4 | 9 | 2 |
| **Lambdavirus** | 6 | 0 | 0 | 0 | 0 | 0 | 0 | 0 |
| **Hedwigvirus** | 6 | 3 | 0 | 7 | 0 | 2 | 2 | 0 |
| **Unahavirus** | 6 | 1 | 2 | 5 | 0 | 0 | 32 | 17 |
| **Ahduovirus** | 6 | 5 | 6 | 1 | 1 | 3 | 2 | 0 |
| **Xipdecavirus** | 5 | 2 | 1 | 0 | 1 | 13 | 1 | 0 |
| **Dhillonvirus** | 5 | 21 | 2 | 1 | 13 | 53 | 3 | 10 |
| **Gilesvirus** | 5 | 3 | 0 | 1 | 81 | 6 | 1 | 0 |
| **Bantamvirus** | 5 | 6 | 5 | 26 | 8 | 2 | 1 | 1 |
| **Hawkeyevirus** | 5 | 1 | 0 | 1 | 0 | 1 | 0 | 1 |
| **Soupsvirus** | 4 | 6 | 2 | 0 | 1 | 13 | 0 | 3 |
| **Titanvirus** | 4 | 6 | 3 | 4 | 0 | 0 | 110 | 16 |
| **Papyrusvirus** | 4 | 5 | 0 | 0 | 10 | 0 | 2 | 1 |
| **Woesvirus** | 4 | 1 | 2 | 5 | 2 | 3 | 3 | 1 |
| **Nyceiraevirus** | 4 | 0 | 1 | 3 | 3 | 0 | 4 | 0 |
| **Andromedavirus** | 4 | 0 | 1 | 0 | 0 | 2 | 1 | 2 |
| **Attisvirus** | 4 | 0 | 2 | 0 | 1 | 4 | 0 | 0 |
| **Hendrixvirus** | 4 | 8 | 4 | 2 | 5 | 7 | 12 | 14 |

| | | | | | | | | |
|---|---|---|---|---|---|---|---|---|
| **Wildcatvirus** | 3 | 0 | 0 | 0 | 5 | 2 | 2 | 0 |
| **Bongovirus** | 3 | 1 | 1 | 0 | 2 | 2 | 2 | 1 |
| **Vendettavirus** | 3 | 0 | 1 | 0 | 0 | 1 | 0 | 1 |
| **Limdunavirus** | 3 | 4 | 5 | 0 | 1 | 4 | 3 | 1 |
| **Pulverervirus** | 3 | 0 | 2 | 1 | 1 | 3 | 0 | 0 |
| **Bernalvirus** | 3 | 0 | 1 | 1 | 3 | 0 | 1 | 0 |
| **Mudcatvirus** | 3 | 2 | 0 | 9 | 1 | 13 | 1 | 3 |
| **Cinunavirus** | 3 | 1 | 0 | 1 | 0 | 3 | 1 | 0 |
| **Eyrevirus** | 3 | 0 | 0 | 4 | 5 | 0 | 1 | 1 |
| **Stanholtvirus** | 3 | 1 | 0 | 1 | 0 | 0 | 1 | 1 |
| **Beetrevirus** | 2 | 2 | 0 | 2 | 0 | 1 | 1 | 1 |
| **Bendigovirus** | 2 | 0 | 0 | 1 | 0 | 0 | 1 | 1 |
| **Bowservirus** | 2 | 0 | 0 | 2 | 4 | 0 | 0 | 0 |
| **Chunghsingvirus** | 2 | 0 | 0 | 1 | 0 | 0 | 0 | 0 |
| **Galunavirus** | 2 | 1 | 0 | 6 | 5 | 1 | 0 | 1 |
| **Gesputvirus** | 2 | 0 | 0 | 2 | 1 | 0 | 1 | 3 |
| **Klementvirus** | 2 | 13 | 0 | 1 | 0 | 1 | 0 | 0 |
| **Yvonnevirus** | 2 | 0 | 1 | 2 | 1 | 0 | 1 | 1 |
| **Xiamenvirus** | 2 | 1 | 1 | 1 | 0 | 1 | 0 | 0 |
| **Slashvirus** | 2 | 1 | 1 | 0 | 0 | 2 | 1 | 0 |
| **Demosthenesvirus** | 2 | 3 | 2 | 51 | 2 | 5 | 2 | 1 |
| **Decurrovirus** | 1 | 0 | 2 | 0 | 0 | 1 | 0 | 1 |
| **Helsingorvirus** | 1 | 0 | 0 | 1 | 0 | 0 | 0 | 9 |
| **Homburgvirus** | 1 | 2 | 4 | 0 | 2 | 1 | 0 | 1 |
| **Myunavirus** | 1 | 1 | 1 | 0 | 0 | 0 | 0 | 0 |
| **Lomovskayavirus** | 1 | 5 | 1 | 5 | 9 | 3 | 1 | 0 |
| **Saphexavirus** | 1 | 0 | 1 | 0 | 0 | 0 | 1 | 9 |
| **Marvinvirus** | 1 | 3 | 2 | 1 | 2 | 2 | 5 | 0 |
| **Samistivirus** | 1 | 0 | 0 | 1 | 2 | 1 | 0 | 0 |
| **Gaiavirus** | 1 | 0 | 1 | 0 | 12 | 3 | 6 | 2 |
| **Sasvirus** | 1 | 0 | 1 | 0 | 0 | 0 | 0 | 1 |
| **Pbi1virus** | 1 | 0 | 0 | 1 | 3 | 1 | 2 | 0 |
| **Steinhofvirus** | 1 | 0 | 0 | 0 | 0 | 2 | 0 | 2 |
| **Kelleziovirus** | 1 | 1 | 0 | 0 | 3 | 0 | 1 | 1 |
| **Gamtrevirus** | 1 | 1 | 0 | 0 | 0 | 0 | 0 | 0 |
| **Jwalphavirus** | 387 | 12 | 23 | 91 | 72 | 41 | 35 | 58 |
| **Teseptimavirus** | 181 | 147 | 70 | 83 | 78 | 154 | 275 | 53 |
| **Friunavirus** | 51 | 72 | 35 | 44 | 8400 | 59 | 71 | 21 |
| **Phikmvvirus** | 18 | 9 | 5 | 468 | 12 | 15 | 28 | 7 |
| **Pradovirus** | 13 | 18 | 2 | 65 | 10 | 6 | 10 | 3 |

| | | | | | | | | |
|---|---|---|---|---|---|---|---|---|
| **Zindervirus** | 9 | 5 | 11 | 10 | 513 | 24 | 31 | 27 |
| **Przondovirus** | 2 | 0 | 0 | 1 | 0 | 2 | 0 | 5 |
| **Phimunavirus** | 2 | 10 | 3 | 0 | 2 | 1 | 2 | 81 |
| **Johnsonvirus** | 80 | 2 | 6 | 38 | 11 | 5 | 8 | 31 |
| **Gamaleyavirus** | 40 | 6 | 4 | 12 | 71 | 6 | 11 | 21 |
| **Rosenblumvirus** | 4 | 1 | 0 | 1 | 7 | 1 | 3 | 2 |
| **Salasvirus** | 1 | 0 | 0 | 0 | 0 | 4 | 2 | 1 |
| **Lessievirus** | 23 | 19 | 23 | 10 | 21 | 10 | 20 | 17 |
| **Lederbergvirus** | 22 | 2 | 0 | 0 | 2 | 4 | 2 | 1 |
| **Litunavirus** | 21 | 7 | 2 | 16 | 0 | 7 | 16 | 6 |
| **Luzseptimavirus** | 11 | 1 | 3 | 6 | 2 | 5 | 12 | 4 |
| **Kochitakasuvirus** | 8 | 1 | 11 | 8 | 2 | 3 | 8 | 1 |
| **Pagevirus** | 4 | 0 | 0 | 0 | 2 | 4 | 0 | 1 |
| **Ithacavirus** | 4 | 2 | 0 | 0 | 7 | 6 | 8 | 20 |
| **Hollowayvirus** | 4 | 0 | 0 | 3 | 6 | 6 | 6 | 4 |
| **Bruynoghevirus** | 4 | 0 | 0 | 0 | 0 | 1 | 3 | 1 |
| **Rauchvirus** | 3 | 1 | 1 | 0 | 1 | 2 | 0 | 0 |
| **Fipvunavirus** | 3 | 5 | 1 | 2 | 3 | 2 | 28 | 48 |
| **Kuravirus** | 2 | 2 | 2 | 0 | 1 | 1 | 1 | 4 |
| **Uetakevirus** | 2 | 1 | 0 | 1 | 1 | 0 | 4 | 0 |
| **Lightbulbvirus** | 2 | 1 | 1 | 1 | 2 | 2 | 1 | 4 |
| **Schmidvirus** | 1 | 0 | 0 | 0 | 1 | 0 | 0 | 0 |
| **Baltimorevirus** | 1 | 0 | 0 | 0 | 0 | 0 | 1 | 0 |
| **Bifseptvirus** | 1 | 1 | 0 | 3 | 2 | 1 | 0 | 1 |
| **Enhodamvirus** | 1 | 0 | 5 | 1 | 0 | 2 | 1 | 0 |
| **Wphvirus** | 12 | 2 | 9 | 1 | 2 | 14 | 8 | 6 |
| **Caeruleovirus** | 6 | 1 | 3 | 4 | 2 | 1 | 2 | 8 |
| **Tsarbombavirus** | 5 | 1 | 1 | 0 | 0 | 6 | 2 | 0 |
| **Bastillevirus** | 3 | 2 | 0 | 1 | 0 | 4 | 1 | 5 |
| **Agatevirus** | 1 | 0 | 0 | 0 | 0 | 1 | 0 | 2 |
| **Silviavirus** | 5 | 2 | 1 | 0 | 1 | 3 | 1 | 8 |
| **Kayvirus** | 2 | 0 | 0 | 0 | 14 | 4 | 1 | 14 |
| **Twortvirus** | 1 | 3 | 2 | 1 | 2 | 5 | 4 | 5 |
| **Okubovirus** | 1 | 2 | 0 | 1 | 1 | 0 | 2 | 0 |
| **Kochikohdavirus** | 2 | 0 | 0 | 2 | 1 | 0 | 2 | 1 |
| **Agtrevirus** | 1 | 0 | 0 | 0 | 0 | 0 | 2 | 1 |
| **Limestonevirus** | 1 | 0 | 0 | 0 | 0 | 0 | 4 | 2 |
| **Kuttervirus** | 2 | 0 | 7 | 3 | 1 | 5 | 26 | 4 |
| **Pandoravirus** | 310 | 269 | 214 | 240 | 147 | 266 | 233 | 119 |
| **Pithovirus** | 3 | 3 | 3 | 0 | 3 | 3 | 2 | 8 |

| | | | | | | | | |
|---|---|---|---|---|---|---|---|---|
| **Betabaculovirus** | 204 | 129 | 25 | 70 | 60 | 111 | 184 | 48 |
| **Alphabaculovirus** | 91 | 65 | 40 | 50 | 33 | 76 | 100 | 83 |
| **Gammabaculovirus** | 3 | 0 | 0 | 0 | 1 | 0 | 0 | 0 |
| **Varicellovirus** | 65 | 41 | 50 | 35 | 37 | 46 | 114 | 21 |
| **Simplexvirus** | 51 | 26 | 13 | 50 | 26 | 22 | 22 | 8 |
| **Mardivirus** | 7 | 3 | 3 | 3 | 3 | 2 | 3 | 27 |
| **Iltovirus** | 4 | 1 | 11 | 3 | 1 | 2 | 3 | 1 |
| **Scutavirus** | 2 | 1 | 0 | 0 | 0 | 0 | 1 | 0 |
| **Cytomegalovirus** | 31 | 20 | 24 | 15 | 6 | 5 | 19 | 16 |
| **Muromegalovirus** | 16 | 31 | 17 | 8 | 4 | 7 | 17 | 1 |
| **Roseolovirus** | 6 | 5 | 5 | 0 | 2 | 2 | 6 | 6 |
| **Proboscivirus** | 5 | 3 | 1 | 5 | 2 | 3 | 2 | 4 |
| **Rhadinovirus** | 7 | 5 | 0 | 10 | 4 | 3 | 6 | 4 |
| **Lymphocryptovirus** | 4 | 2 | 2 | 0 | 7 | 2 | 2 | 1 |
| **Percavirus** | 3 | 1 | 2 | 1 | 1 | 0 | 1 | 3 |
| **Cyprinivirus** | 19 | 16 | 13 | 18 | 9 | 7 | 10 | 2 |
| **Batrachovirus** | 1 | 3 | 14 | 0 | 1 | 2 | 3 | 0 |
| **Aurivirus** | 3 | 0 | 1 | 1 | 0 | 1 | 2 | 0 |
| **Betapolyomavirus** | 169 | 7 | 2 | 14 | 6 | 6 | 30 | 4 |
| **Alphapolyomavirus** | 6 | 1 | 1 | 0 | 1 | 0 | 0 | 2 |
| **Nucleorhabdovirus** | 5 | 3 | 0 | 0 | 0 | 0 | 0 | 0 |
| **Almendravirus** | 2 | 0 | 0 | 0 | 0 | 1 | 0 | 0 |
| **Cytorhabdovirus** | 1 | 0 | 0 | 0 | 0 | 0 | 0 | 0 |
| **Rubulavirus** | 5 | 2 | 2 | 0 | 1 | 1 | 4 | 0 |
| **Morbillivirus** | 1 | 0 | 0 | 0 | 0 | 0 | 3 | 2 |
| **Orthobornavirus** | 3 | 0 | 0 | 0 | 0 | 0 | 0 | 0 |
| **Metapneumovirus** | 1 | 0 | 0 | 0 | 1 | 0 | 0 | 0 |
| **Ophiovirus** | 1 | 0 | 0 | 0 | 0 | 0 | 0 | 0 |
| **Tospovirus** | 5 | 11 | 0 | 0 | 0 | 0 | 0 | 0 |
| **Orthobunyavirus** | 2 | 3 | 1 | 2 | 1 | 2 | 7 | 0 |
| **Mammarenavirus** | 2 | 1 | 0 | 1 | 0 | 3 | 1 | 0 |
| **Orthonairovirus** | 2 | 0 | 0 | 1 | 1 | 0 | 1 | 0 |
| **Orthotospovirus** | 2 | 0 | 1 | 0 | 0 | 0 | 2 | 0 |
| **Emaravirus** | 1 | 0 | 0 | 1 | 1 | 0 | 0 | 5 |
| **Tenuivirus** | 1 | 1 | 0 | 0 | 0 | 0 | 0 | 0 |
| **Alphainfluenzavirus** | 1 | 1 | 0 | 0 | 1 | 0 | 0 | 0 |
| **Epsilonarterivirus** | 7 | 0 | 0 | 1 | 0 | 0 | 1 | 0 |
| **Betaarterivirus** | 1 | 1 | 0 | 0 | 0 | 1 | 0 | 0 |
| **Gammaarterivirus** | 1 | 0 | 0 | 0 | 0 | 0 | 0 | 0 |
| **Betacoronavirus** | 5 | 4 | 3 | 3 | 2 | 7 | 4 | 6 |

| | | | | | | | | |
|---|---|---|---|---|---|---|---|---|
| **Alphacoronavirus** | 3 | 3 | 2 | 6 | 0 | 10 | 0 | 1 |
| **Deltacoronavirus** | 1 | 1 | 0 | 0 | 0 | 1 | 0 | 0 |
| **Bostovirus** | 2 | 0 | 0 | 0 | 1 | 0 | 1 | 0 |
| **Tobamovirus** | 6 | 2 | 0 | 3 | 0 | 2 | 4 | 3 |
| **Furovirus** | 6 | 0 | 0 | 1 | 0 | 0 | 0 | 2 |
| **Alphaendornavirus** | 9 | 5 | 4 | 9 | 0 | 8 | 10 | 3 |
| **Nepovirus** | 2 | 0 | 0 | 0 | 0 | 0 | 0 | 9 |
| **Fabavirus** | 1 | 0 | 0 | 0 | 0 | 0 | 0 | 0 |
| **Waikavirus** | 1 | 0 | 0 | 0 | 0 | 0 | 0 | 0 |
| **Iflavirus** | 1 | 0 | 0 | 1 | 0 | 0 | 0 | 0 |
| **Kobuvirus** | 1 | 0 | 0 | 0 | 0 | 0 | 0 | 0 |
| **Cosavirus** | 1 | 1 | 5 | 0 | 0 | 1 | 3 | 0 |
| **Robigovirus** | 6 | 0 | 0 | 0 | 0 | 3 | 0 | 2 |
| **Carlavirus** | 3 | 2 | 0 | 1 | 0 | 0 | 1 | 0 |
| **Tymovirus** | 1 | 1 | 0 | 0 | 0 | 4 | 0 | 1 |
| **Aquareovirus** | 5 | 2 | 0 | 11 | 0 | 2 | 0 | 0 |
| **Fijivirus** | 1 | 0 | 0 | 0 | 0 | 0 | 0 | 0 |
| **Orbivirus** | 2 | 4 | 0 | 1 | 2 | 0 | 3 | 2 |
| **Negevirus** | 1 | 0 | 0 | 0 | 0 | 0 | 0 | 0 |
| **Mamastrovirus** | 1 | 1 | 0 | 0 | 0 | 2 | 1 | 1 |
| **Polerovirus** | 3 | 6 | 1 | 0 | 0 | 1 | 0 | 0 |
| **Flavivirus** | 2 | 0 | 3 | 9 | 0 | 4 | 3 | 0 |
| **Potyvirus** | 2 | 2 | 1 | 7 | 1 | 3 | 9 | 1 |
| **Ilarvirus** | 2 | 0 | 1 | 0 | 1 | 1 | 0 | 2 |
| **Velarivirus** | 1 | 0 | 0 | 0 | 0 | 0 | 1 | 0 |
| **Narnavirus** | 1 | 0 | 0 | 0 | 5 | 0 | 0 | 0 |
| **Benyvirus** | 1 | 0 | 0 | 0 | 1 | 1 | 30 | 0 |
| **Picobirnavirus** | 1 | 1 | 0 | 0 | 0 | 1 | 0 | 0 |
| **Polemovirus** | 1 | 0 | 0 | 0 | 0 | 0 | 0 | 0 |
| **Totivirus** | 1 | 0 | 0 | 0 | 0 | 0 | 0 | 0 |
| **Mimivirus** | 141 | 79 | 63 | 75 | 32 | 103 | 88 | 135 |
| **Cafeteriavirus** | 8 | 6 | 4 | 4 | 1 | 8 | 5 | 16 |
| **Parapoxvirus** | 24 | 25 | 15 | 16 | 11 | 21 | 13 | 14 |
| **Molluscipoxvirus** | 19 | 3 | 1 | 5 | 2 | 2 | 4 | 2 |
| **Orthopoxvirus** | 17 | 6 | 0 | 1 | 3 | 7 | 6 | 10 |
| **Avipoxvirus** | 7 | 6 | 8 | 5 | 5 | 10 | 6 | 10 |
| **Suipoxvirus** | 7 | 8 | 0 | 0 | 1 | 3 | 3 | 0 |
| **Cervidpoxvirus** | 3 | 0 | 0 | 1 | 0 | 0 | 0 | 1 |
| **Centapoxvirus** | 3 | 2 | 1 | 1 | 2 | 2 | 0 | 9 |
| **Crocodylidpoxvirus** | 2 | 3 | 1 | 2 | 2 | 5 | 2 | 0 |

| | | | | | | | | |
|---|---|---|---|---|---|---|---|---|
| **Capripoxvirus** | 1 | 1 | 0 | 1 | 0 | 2 | 0 | 14 |
| **Leporipoxvirus** | 1 | 3 | 1 | 1 | 1 | 4 | 2 | 0 |
| **Yatapoxvirus** | 1 | 0 | 0 | 2 | 0 | 0 | 1 | 1 |
| **Betaentomopoxvirus** | 17 | 16 | 4 | 5 | 2 | 8 | 9 | 17 |
| **Alphaentomopoxvirus** | 7 | 5 | 2 | 1 | 1 | 5 | 2 | 7 |
| **Prasinovirus** | 33 | 11 | 8 | 29 | 8 | 13 | 11 | 5 |
| **Chlorovirus** | 18 | 25 | 3 | 26 | 8 | 18 | 22 | 17 |
| **Coccolithovirus** | 15 | 11 | 1 | 6 | 1 | 11 | 6 | 5 |
| **Raphidovirus** | 5 | 1 | 1 | 2 | 1 | 1 | 2 | 3 |
| **Prymnesiovirus** | 4 | 6 | 0 | 1 | 4 | 4 | 3 | 13 |
| **Phaeovirus** | 3 | 1 | 0 | 2 | 1 | 3 | 0 | 2 |
| **Alphapapillomavirus** | 20 | 0 | 0 | 3 | 0 | 8 | 0 | 0 |
| **Gammapapillomavirus** | 11 | 1 | 2 | 3 | 0 | 7 | 3 | 0 |
| **Betapapillomavirus** | 2 | 0 | 0 | 3 | 0 | 1 | 0 | 0 |
| **Rhopapillomavirus** | 1 | 0 | 0 | 0 | 0 | 0 | 0 | 0 |
| **Iotapapillomavirus** | 1 | 0 | 0 | 0 | 0 | 0 | 0 | 2 |
| **Mastadenovirus** | 25 | 8 | 8 | 16 | 6 | 21 | 15 | 25 |
| **Aviadenovirus** | 5 | 6 | 4 | 5 | 4 | 3 | 8 | 0 |
| **Atadenovirus** | 3 | 1 | 0 | 1 | 5 | 3 | 0 | 0 |
| **Iridovirus** | 9 | 3 | 0 | 4 | 7 | 9 | 4 | 35 |
| **Chloriridovirus** | 5 | 1 | 2 | 3 | 2 | 3 | 7 | 16 |
| **Ranavirus** | 7 | 2 | 2 | 1 | 1 | 5 | 4 | 2 |
| **Lymphocystivirus** | 3 | 3 | 2 | 3 | 2 | 3 | 39 | 14 |
| **Megalocytivirus** | 1 | 1 | 0 | 0 | 0 | 0 | 1 | 4 |
| **Marseillevirus** | 20 | 14 | 10 | 14 | 8 | 9 | 14 | 6 |
| **Inovirus** | 6 | 0 | 0 | 4 | 0 | 2 | 0 | 0 |
| **Gemycircularvirus** | 7 | 2 | 1 | 0 | 2 | 24 | 2 | 0 |
| **Gemykibivirus** | 5 | 6 | 18 | 12 | 3 | 1 | 1 | 0 |
| **Muscavirus** | 8 | 0 | 0 | 1 | 1 | 0 | 0 | 0 |
| **Glossinavirus** | 3 | 1 | 0 | 0 | 2 | 1 | 1 | 3 |
| **Begomovirus** | 8 | 6 | 2 | 2 | 7 | 17 | 7 | 5 |
| **Ichnovirus** | 5 | 0 | 0 | 1 | 0 | 1 | 1 | 13 |
| **Bracovirus** | 5 | 2 | 1 | 0 | 0 | 1 | 0 | 3 |
| **Alphasphaerolipovirus** | 9 | 1 | 1 | 3 | 2 | 1 | 2 | 3 |
| **Alphanudivirus** | 2 | 1 | 0 | 0 | 0 | 1 | 2 | 0 |
| **Betanudivirus** | 2 | 3 | 0 | 3 | 1 | 0 | 0 | 1 |
| **Rudivirus** | 6 | 1 | 0 | 1 | 0 | 1 | 0 | 1 |
| **Betalipothrixvirus** | 1 | 0 | 1 | 0 | 0 | 0 | 2 | 1 |
| **Badnavirus** | 5 | 6 | 5 | 1 | 0 | 2 | 3 | 8 |
| **Caulimovirus** | 2 | 5 | 0 | 0 | 0 | 0 | 0 | 0 |

| | | | | | | | | |
|---|---|---|---|---|---|---|---|---|
| **Whispovirus** | 6 | 5 | 5 | 1 | 2 | 3 | 6 | 0 |
| **Circovirus** | 2 | 8 | 0 | 1 | 0 | 4 | 2 | 1 |
| **Bicaudavirus** | 6 | 8 | 0 | 3 | 1 | 5 | 1 | 1 |
| **Bocaparvovirus** | 5 | 5 | 4 | 0 | 1 | 4 | 3 | 1 |
| **Ambidensovirus** | 1 | 0 | 0 | 0 | 0 | 0 | 1 | 0 |
| **Sputnikvirus** | 2 | 0 | 0 | 1 | 0 | 0 | 1 | 2 |
| **Mavirus** | 1 | 0 | 0 | 0 | 0 | 0 | 0 | 0 |
| **Alphafusellovirus** | 1 | 0 | 0 | 0 | 0 | 0 | 0 | 0 |
| **Betafusellovirus** | 1 | 0 | 0 | 0 | 0 | 3 | 1 | 0 |
| **Alphatectivirus** | 2 | 1 | 0 | 0 | 0 | 0 | 6 | 6 |
| **Nanovirus** | 2 | 0 | 0 | 0 | 0 | 0 | 2 | 0 |
| **Alphaturrivirus** | 2 | 0 | 0 | 0 | 0 | 0 | 0 | 0 |
| **Alphatorquevirus** | 1 | 0 | 0 | 0 | 0 | 0 | 1 | 0 |
| **Ampullavirus** | 1 | 0 | 0 | 0 | 0 | 0 | 0 | 0 |
| **Porprismacovirus** | 1 | 0 | 0 | 0 | 0 | 0 | 0 | 0 |
| **Betasatellite** | 1 | 1 | 0 | 0 | 0 | 2 | 0 | 0 |
| **Drulisvirus** | 0 | 2 | 1 | 1 | 1 | 4 | 0 | 25 |
| **Enquatrovirus** | 0 | 1 | 0 | 4 | 1 | 2 | 2 | 3 |
| **Tunavirus** | 0 | 0 | 0 | 0 | 3 | 2 | 1 | 1 |
| **Ravinvirus** | 0 | 2 | 204 | 0 | 1 | 4 | 3 | 7 |
| **Bignuzvirus** | 0 | 2 | 1 | 0 | 1 | 2 | 1 | 0 |
| **Detrevirus** | 0 | 1 | 1 | 0 | 0 | 1 | 1 | 0 |
| **Psavirus** | 0 | 0 | 0 | 0 | 0 | 0 | 0 | 1 |
| **Cimpunavirus** | 0 | 0 | 2 | 0 | 0 | 1 | 0 | 3 |
| **Coetzeevirus** | 0 | 0 | 0 | 0 | 0 | 0 | 0 | 0 |
| **Ghobesvirus** | 0 | 0 | 1 | 0 | 1 | 0 | 0 | 0 |
| **Krischvirus** | 0 | 2 | 6 | 1 | 0 | 6 | 1 | 7 |
| **Punavirus** | 0 | 1 | 0 | 5 | 0 | 0 | 0 | 4 |
| **Lambdaarterivirus** | 0 | 1 | 0 | 0 | 0 | 22 | 0 | 1 |
| **Phlebovirus** | 0 | 2 | 0 | 0 | 1 | 7 | 1 | 3 |
| **Isavirus** | 0 | 0 | 0 | 0 | 0 | 0 | 0 | 5 |
| **Sigmavirus** | 0 | 0 | 0 | 0 | 0 | 0 | 0 | 0 |
| **Sprivivirus** | 0 | 0 | 0 | 0 | 0 | 0 | 0 | 0 |
| **Hapavirus** | 0 | 0 | 0 | 0 | 1 | 0 | 1 | 0 |
| **Potexvirus** | 0 | 4 | 0 | 0 | 1 | 0 | 1 | 0 |
| **Chrysovirus** | 0 | 0 | 0 | 1 | 0 | 0 | 0 | 0 |
| **Macavirus** | 0 | 10 | 2 | 1 | 2 | 6 | 1 | 2 |
| **Lentivirus** | 0 | 0 | 0 | 0 | 4 | 0 | 0 | 0 |
| **Deltapolyomavirus** | 0 | 0 | 0 | 0 | 0 | 0 | 0 | 0 |
| **Gequatrovirus** | 0 | 0 | 0 | 0 | 0 | 0 | 1 | 0 |

| | | | | | | | | |
|---|---|---|---|---|---|---|---|---|
| **Ascovirus** | 0 | 2 | 1 | 0 | 0 | 1 | 2 | 0 |
| **Betatorquevirus** | 0 | 34 | 0 | 1 | 0 | 0 | 0 | 0 |
| **Unaquatrovirus** | 0 | 27 | 0 | 0 | 1 | 11 | 1 | 2 |
| **Rerduovirus** | 0 | 2 | 0 | 0 | 0 | 0 | 1 | 0 |
| **Incheonvrus** | 0 | 2 | 0 | 3 | 0 | 1 | 11 | 24 |
| **Plotvirus** | 0 | 1 | 0 | 0 | 0 | 0 | 0 | 1 |
| **Mardecavirus** | 0 | 2 | 0 | 0 | 0 | 2 | 0 | 0 |
| **Oshimavirus** | 0 | 1 | 0 | 11 | 2 | 0 | 1 | 0 |
| **Nonanavirus** | 0 | 1 | 1 | 0 | 0 | 1 | 1 | 2 |
| **Inhavirus** | 0 | 1 | 0 | 0 | 1 | 1 | 0 | 2 |
| **Eiauvirus** | 0 | 1 | 0 | 1 | 0 | 1 | 0 | 0 |
| **Tijeunavirus** | 0 | 2 | 0 | 7 | 2 | 1 | 2 | 0 |
| **Jimmervirus** | 0 | 1 | 0 | 0 | 0 | 0 | 1 | 0 |
| **Nitunavirus** | 0 | 1 | 0 | 0 | 1 | 1 | 11 | 2 |
| **Krylovvirus** | 0 | 2 | 2 | 0 | 0 | 3 | 2 | 1 |
| **Myxoctovirus** | 0 | 1 | 2 | 3 | 1 | 2 | 0 | 0 |
| **Pecentumvirus** | 0 | 2 | 0 | 0 | 0 | 11 | 1 | 5 |
| **Deltabaculovirus** | 0 | 1 | 1 | 0 | 0 | 4 | 0 | 0 |
| **Ictalurivirus** | 0 | 1 | 2 | 2 | 1 | 0 | 0 | 0 |
| **Herbevirus** | 0 | 1 | 0 | 0 | 0 | 0 | 1 | 0 |
| **Horwuvirus** | 0 | 1 | 0 | 0 | 0 | 0 | 0 | 0 |
| **Shaspivirus** | 0 | 1 | 0 | 0 | 0 | 0 | 0 | 0 |
| **Novirhabdovirus** | 0 | 1 | 0 | 0 | 0 | 0 | 0 | 0 |
| **Avulavirus** | 0 | 1 | 0 | 1 | 0 | 1 | 0 | 0 |
| **Ebolavirus** | 0 | 1 | 4 | 0 | 0 | 0 | 1 | 2 |
| **Yingvirus** | 0 | 1 | 0 | 0 | 0 | 0 | 0 | 1 |
| **Foveavirus** | 0 | 4 | 0 | 0 | 1 | 0 | 1 | 0 |
| **Maculavirus** | 0 | 1 | 0 | 0 | 0 | 0 | 0 | 0 |
| **Gammacoronavirus** | 0 | 1 | 0 | 0 | 0 | 0 | 0 | 0 |
| **Paguronivirus** | 0 | 1 | 0 | 0 | 0 | 0 | 0 | 0 |
| **Ampivirus** | 0 | 1 | 0 | 0 | 1 | 0 | 0 | 0 |
| **Torradovirus** | 0 | 1 | 0 | 0 | 0 | 0 | 0 | 0 |
| **Orthoreovirus** | 0 | 2 | 0 | 0 | 0 | 0 | 0 | 0 |
| **Victorivirus** | 0 | 2 | 0 | 0 | 0 | 5 | 3 | 0 |
| **Pestivirus** | 0 | 1 | 0 | 0 | 0 | 1 | 0 | 0 |
| **Pegivirus** | 0 | 1 | 0 | 2 | 1 | 1 | 0 | 0 |
| **Betatetravirus** | 0 | 1 | 0 | 0 | 0 | 0 | 0 | 0 |
| **Alphavirus** | 0 | 1 | 2 | 0 | 0 | 0 | 0 | 1 |
| **Norovirus** | 0 | 1 | 0 | 0 | 0 | 0 | 0 | 0 |
| **Gammatorquevirus** | 0 | 1 | 0 | 0 | 0 | 0 | 0 | 0 |

| | | | | | | | | |
|---|---|---|---|---|---|---|---|---|
| **Solendovirus** | 0 | 1 | 0 | 0 | 0 | 0 | 0 | 0 |
| **Gammaretrovirus** | 0 | 10 | 1 | 0 | 2 | 1 | 0 | 2 |
| **Betaretrovirus** | 0 | 1 | 0 | 0 | 0 | 0 | 0 | 0 |
| **Mastrevirus** | 0 | 1 | 0 | 1 | 1 | 0 | 0 | 0 |
| **Etapapillomavirus** | 0 | 1 | 0 | 0 | 0 | 0 | 0 | 0 |
| **Dyoiotapapillomavirus** | 0 | 1 | 0 | 0 | 1 | 0 | 0 | 0 |
| **Chipapillomavirus** | 0 | 1 | 0 | 0 | 0 | 0 | 1 | 0 |
| **Gammasphaerolipovirus** | 0 | 1 | 0 | 0 | 0 | 0 | 0 | 0 |
| **Cecivirus** | 0 | 0 | 1 | 0 | 0 | 0 | 1 | 0 |
| **Lubbockvirus** | 0 | 0 | 1 | 0 | 0 | 0 | 0 | 0 |
| **Ledantevirus** | 0 | 0 | 1 | 0 | 0 | 0 | 1 | 0 |
| **Dichorhavirus** | 0 | 0 | 1 | 0 | 0 | 0 | 0 | 0 |
| **Tibrovirus** | 0 | 0 | 1 | 0 | 0 | 0 | 0 | 0 |
| **Luteovirus** | 0 | 0 | 2 | 1 | 0 | 1 | 0 | 0 |
| **Orthohepevirus** | 0 | 0 | 1 | 0 | 0 | 2 | 0 | 0 |
| **Pipapillomavirus** | 0 | 0 | 2 | 0 | 0 | 0 | 0 | 1 |
| **Dyomupapillomavirus** | 0 | 0 | 1 | 0 | 0 | 0 | 1 | 5 |
| **Alphatristromavirus** | 0 | 0 | 1 | 0 | 2 | 0 | 0 | 1 |
| **Kleczkowskavirus** | 0 | 0 | 0 | 2 | 0 | 0 | 0 | 0 |
| **Marthavirus** | 0 | 0 | 0 | 1 | 1 | 0 | 3 | 0 |
| **Gorjumvirus** | 0 | 0 | 0 | 1 | 0 | 0 | 0 | 0 |
| **Perisivirus** | 0 | 0 | 0 | 2 | 0 | 1 | 0 | 2 |
| **Rotavirus** | 0 | 0 | 0 | 1 | 0 | 1 | 2 | 1 |
| **Seadornavirus** | 0 | 0 | 0 | 1 | 0 | 0 | 0 | 0 |
| **Orthohantavirus** | 0 | 0 | 0 | 2 | 0 | 6 | 1 | 1 |
| **Sripuvirus** | 0 | 0 | 0 | 2 | 0 | 0 | 2 | 0 |
| **Bymovirus** | 0 | 0 | 0 | 1 | 1 | 0 | 0 | 0 |
| **Trichovirus** | 0 | 0 | 0 | 1 | 0 | 0 | 1 | 0 |
| **Marafivirus** | 0 | 0 | 0 | 1 | 0 | 1 | 0 | 0 |
| **Allexivirus** | 0 | 0 | 0 | 1 | 0 | 0 | 0 | 0 |
| **Siadenovirus** | 0 | 0 | 0 | 1 | 0 | 0 | 1 | 0 |
| **Omikronpapillomavirus** | 0 | 0 | 0 | 1 | 0 | 0 | 0 | 0 |
| **Equispumavirus** | 0 | 0 | 0 | 1 | 0 | 0 | 0 | 0 |
| **Alphatrevirus** | 0 | 0 | 0 | 2 | 11 | 2 | 6 | 0 |
| **Getseptimavirus** | 0 | 0 | 0 | 0 | 1 | 0 | 0 | 0 |
| **Sitaravirus** | 0 | 0 | 0 | 0 | 1 | 0 | 0 | 0 |
| **Cbastvirus** | 0 | 0 | 0 | 0 | 1 | 0 | 1 | 1 |
| **Marburgvirus** | 0 | 0 | 0 | 0 | 1 | 0 | 0 | 0 |
| **Hepatovirus** | 0 | 0 | 0 | 0 | 1 | 0 | 1 | 0 |
| **Mosavirus** | 0 | 0 | 0 | 0 | 1 | 0 | 0 | 0 |

| | | | | | | | | |
|---|---|---|---|---|---|---|---|---|
| **Comovirus** | 0 | 0 | 0 | 0 | 1 | 0 | 0 | 0 |
| **Sobemovirus** | 0 | 0 | 0 | 0 | 2 | 0 | 0 | 0 |
| **Invictavirus** | 0 | 0 | 0 | 0 | 2 | 0 | 0 | 0 |
| **Pomovirus** | 0 | 0 | 0 | 0 | 1 | 0 | 0 | 0 |
| **Alphapartitivirus** | 0 | 0 | 0 | 0 | 1 | 1 | 3 | 0 |
| **Vesivirus** | 0 | 0 | 0 | 0 | 1 | 0 | 0 | 0 |
| **Xipapillomavirus** | 0 | 0 | 0 | 0 | 1 | 0 | 0 | 0 |
| **Dyorhopapillomavirus** | 0 | 0 | 0 | 0 | 1 | 0 | 0 | 0 |
| **Amdoparvovirus** | 0 | 0 | 0 | 0 | 1 | 2 | 0 | 3 |
| **Jedunavirus** | 0 | 0 | 0 | 0 | 0 | 1 | 0 | 31 |
| **Traversvirus** | 0 | 0 | 0 | 0 | 0 | 1 | 0 | 0 |
| **Torovirus** | 0 | 0 | 0 | 0 | 0 | 1 | 0 | 0 |
| **Bafinivirus** | 0 | 0 | 0 | 0 | 0 | 1 | 1 | 6 |
| **Wuhivirus** | 0 | 0 | 0 | 0 | 0 | 1 | 0 | 0 |
| **Vesiculovirus** | 0 | 0 | 0 | 0 | 0 | 4 | 0 | 0 |
| **Respirovirus** | 0 | 0 | 0 | 0 | 0 | 1 | 0 | 10 |
| **Anphevirus** | 0 | 0 | 0 | 0 | 0 | 1 | 0 | 0 |
| **Betaendornavirus** | 0 | 0 | 0 | 0 | 0 | 1 | 0 | 1 |
| **Coltivirus** | 0 | 0 | 0 | 0 | 0 | 1 | 0 | 0 |
| **Cypovirus** | 0 | 0 | 0 | 0 | 0 | 1 | 2 | 1 |
| **Oryzavirus** | 0 | 0 | 0 | 0 | 0 | 1 | 0 | 0 |
| **Hypovirus** | 0 | 0 | 0 | 0 | 0 | 2 | 0 | 0 |
| **Ampelovirus** | 0 | 0 | 0 | 0 | 0 | 1 | 1 | 0 |
| **Lineavirus** | 0 | 0 | 0 | 0 | 0 | 5 | 2 | 1 |
| **Erythroparvovirus** | 0 | 0 | 0 | 0 | 0 | 1 | 0 | 0 |
| **Toursvirus** | 0 | 0 | 0 | 0 | 0 | 3 | 0 | 0 |
| **Corticovirus** | 0 | 0 | 0 | 0 | 0 | 1 | 0 | 0 |
| **Enterovirus** | 0 | 0 | 0 | 0 | 0 | 0 | 2 | 3 |
| **Parechovirus** | 0 | 0 | 0 | 0 | 0 | 0 | 1 | 0 |
| **Sopolycivirus** | 0 | 0 | 0 | 0 | 0 | 0 | 1 | 2 |
| **Dinovernavirus** | 0 | 0 | 0 | 0 | 0 | 0 | 5 | 0 |
| **Ipomovirus** | 0 | 0 | 0 | 0 | 0 | 0 | 1 | 0 |
| **Hepacivirus** | 0 | 0 | 0 | 0 | 0 | 0 | 1 | 1 |
| **Rubivirus** | 0 | 0 | 0 | 0 | 0 | 0 | 3 | 0 |
| **Panicovirus** | 0 | 0 | 0 | 0 | 0 | 0 | 1 | 0 |
| **Spiromicrovirus** | 0 | 0 | 0 | 0 | 0 | 0 | 2 | 0 |
| **Deltapapillomavirus** | 0 | 0 | 0 | 0 | 0 | 0 | 1 | 0 |
| **Taupapillomavirus** | 0 | 0 | 0 | 0 | 0 | 0 | 1 | 0 |
| **Epsilonretrovirus** | 0 | 0 | 0 | 0 | 0 | 0 | 1 | 0 |
| **Prosimiispumavirus** | 0 | 0 | 0 | 0 | 0 | 0 | 1 | 0 |

| | | | | | | | | |
|---|---|---|---|---|---|---|---|---|
| **Iteradensovirus** | 0 | 0 | 0 | 0 | 0 | 0 | 1 | 1 |
| **Orchidvirus** | 0 | 0 | 0 | 0 | 0 | 0 | 0 | 1 |
| **Cepunavirus** | 0 | 0 | 0 | 0 | 0 | 0 | 0 | 1 |
| **Jonvirus** | 0 | 0 | 0 | 0 | 0 | 0 | 0 | 1 |
| **Megrivirus** | 0 | 0 | 0 | 0 | 0 | 0 | 0 | 1 |
| **Harkavirus** | 0 | 0 | 0 | 0 | 0 | 0 | 0 | 1 |
| **Infratovirus** | 0 | 0 | 0 | 0 | 0 | 0 | 0 | 1 |
| **Amalgavirus** | 0 | 0 | 0 | 0 | 0 | 0 | 0 | 1 |
| **Tombusvirus** | 0 | 0 | 0 | 0 | 0 | 0 | 0 | 1 |
| **Bovispumavirus** | 0 | 0 | 0 | 0 | 0 | 0 | 0 | 1 |
| **Dyoxipapillomavirus** | 0 | 0 | 0 | 0 | 0 | 0 | 0 | 1 |
| **Thetapapillomavirus** | 0 | 0 | 0 | 0 | 0 | 0 | 0 | 1 |
| **Gammapolyomavirus** | 0 | 0 | 0 | 0 | 0 | 0 | 0 | 1 |
| **Chapparvovirus** | 0 | 0 | 0 | 0 | 0 | 0 | 0 | 1 |
| **Deltalipothrixvirus** | 0 | 0 | 0 | 0 | 0 | 0 | 0 | 2 |
| **Iotatorquevirus** | 0 | 0 | 0 | 0 | 0 | 0 | 0 | 2 |

Table S3

| Gene group | CP1 | CP3 | CP4 | CP5 | CP6 | CP7 | CP8 | Seafield |
|---|---|---|---|---|---|---|---|---|
| aac(3)-I_1_AJ877225 | 0.00000 | 29.35374 | 0.00000 | 36.41902 | 2.40914 | 10.44301 | 20.17039 | 29.47549 |
| aac(3)-Ia_1_X15852 | 8.30881 | 31.89019 | 7.02658 | 49.30890 | 5.50686 | 14.03017 | 24.73817 | 41.30760 |
| aac(3)-Ib_1_L06157 | 51.52712 | 47.49352 | 1.24699 | 13.99149 | 13.97678 | 31.61560 | 38.06324 | 0.00000 |
| aac(3)-Ib-aac(6_)-Ib__1_AF355189 | 6.03360 | 1.94023 | 0.00000 | 0.00000 | 0.00000 | 1.10442 | 2.10311 | 0.00000 |
| aac(3)-Ic_1_AJ511268 | 0.00000 | 0.00000 | 0.00000 | 0.00000 | 1.18923 | 1.76742 | 0.00000 | 0.00000 |
| aac(3)-IIa_4_L22613 | 99.97203 | 28.98846 | 17.43179 | 951.20903 | 42.93649 | 266.85033 | 208.66258 | 18.90360 |
| aac(3)-IIb_1_M97172 | 11.32049 | 0.00000 | 1.63494 | 0.00000 | 2.76605 | 2.39802 | 0.00000 | 0.00000 |
| aac(3)-IIIb_2_LLLC01000048 | 126.17946 | 15.40683 | 15.00300 | 30.56341 | 66.01921 | 48.72624 | 57.21534 | 0.00000 |
| aac(3)-IIIc_1_L06161 | 86.62525 | 18.36439 | 11.91172 | 18.03609 | 32.67402 | 26.20698 | 40.88866 | 0.00000 |
| aac(3)-IVa_1_X01385 | 0.00000 | 0.00000 | 0.00000 | 0.00000 | 0.00000 | 0.00000 | 0.00000 | 0.36433 |
| aac(6_)-30-aac(6_)-Ib__1_AJ584652 | 6.89286 | 1083.81965 | 1.90081 | 62.76470 | 6.81004 | 6.46623 | 59.15806 | 0.00000 |
| aac(6_)-31_1_AM283489 | 56.70801 | 55.35438 | 6.37911 | 11.78882 | 5.66602 | 6.14853 | 13.99923 | 0.00000 |
| aac(6_)-32_1_EF614235 | 0.00000 | 0.00000 | 0.00000 | 0.00000 | 0.67282 | 0.41665 | 1.34879 | 0.00000 |
| aac(6_)-aph(2__)_1_M13771 | 8.45415 | 2.95443 | 1.82411 | 8.05131 | 4.91846 | 10.41527 | 8.37009 | 0.00000 |
| aac(6_)-Ib-cr_1_DQ303918 | 1.17578 | 43.03644 | 13.84966 | 342.28918 | 37.69162 | 129.52483 | 322.19043 | 0.00000 |
| aac(6_)-Ib-Suzhou_1_EU085533 | 120.25517 | 1966.62524 | 42.31476 | 505.16482 | 34.89546 | 170.37656 | 567.09609 | 0.00000 |
| aac(6_)-If_1_X55353 | 0.00000 | 0.00000 | 0.00000 | 0.00000 | 0.00000 | 0.00000 | 1.21473 | 0.00000 |
| aac(6_)-Ii_1_L12710 | 0.00000 | 0.00000 | 0.53605 | 11.27729 | 15.98417 | 0.00000 | 0.00000 | 0.00000 |
| aac(6_)-IIa_1_M29695 | 19.36426 | 7.65137 | 2.25357 | 5.51207 | 7.06465 | 77.16268 | 15.39204 | 0.00000 |
| aac(6_)-Im_1_AF337947 | 0.00000 | 0.00000 | 0.00000 | 0.00000 | 0.00000 | 1.20571 | 0.00000 | 0.00000 |
| aac(6_)-Iz_1_AF140221 | 0.00000 | 0.00000 | 0.00000 | 0.00000 | 0.00000 | 0.00000 | 3.43122 | 0.00000 |
| aadA10_1_U37105 | 1.53690 | 1.97435 | 2.73471 | 7.24887 | 1.56710 | 2.10721 | 0.52798 | 7.27555 |
| aadA11_1_AY144590 | 124.71104 | 81.62007 | 51.64048 | 115.48550 | 30.93333 | 55.36531 | 65.58024 | 48.15585 |
| aadA13_2_NC010643 | 3.45957 | 12.92349 | 2.39710 | 4.83761 | 7.60405 | 3.12954 | 3.09010 | 3.22042 |
| aadA15_1_DQ393783 | 2.74924 | 38.77337 | 5.12862 | 50.68199 | 4.73279 | 11.37140 | 24.66794 | 5.42859 |
| aadA16_1_EU675686 | 2.68056 | 0.00000 | 0.00000 | 1.37755 | 0.00000 | 3.82663 | 0.00000 | 1.60323 |
| aadA4_1_Z50802 | 46.36223 | 7.13957 | 3.63666 | 12.55510 | 13.84342 | 3.86862 | 16.63132 | 1.99048 |
| aadA7_1_AF224733 | 0.00000 | 0.00000 | 1.38294 | 6.57185 | 0.00000 | 0.92727 | 0.00000 | 0.00000 |
| aadA8b_1_AY139603 | 45.82913 | 2.83829 | 9.91623 | 16.42325 | 3.29261 | 7.45671 | 25.29300 | 2.24802 |
| aadD_1_AF181950 | 0.00000 | 0.00000 | 0.00000 | 0.00000 | 0.00000 | 0.00000 | 0.00000 | 0.55879 |
| ampS_1_X80276 | 0.00000 | 0.00000 | 1.48070 | 4.48939 | 0.00000 | 1.16346 | 1.55088 | 5.61210 |
| ant(2__)-Ia_5_AY139594 | 429.10371 | 982.18550 | 123.17177 | 603.57369 | 1356.26028 | 958.55538 | 2059.37018 | 4.94622 |
| ant(3__)-Ia_1_X02340 | 71.97034 | 276.03906 | 14.49497 | 176.88704 | 49.51046 | 73.79634 | 128.38703 | 6.71997 |
| ant(3__)-Ii-aac(6_)-IId_1_AF453998 | 52.69885 | 66.58515 | 12.05065 | 12.16584 | 8.14839 | 17.14351 | 25.05379 | 0.00000 |
| ant(6)-Ia_2_KF421157 | 3.82295 | 0.00000 | 0.00000 | 0.00000 | 0.00000 | 0.00000 | 1.32510 | 0.41311 |
| ant(6)-Ia_3_KF864551 | 39.29130 | 8.29650 | 3.22463 | 3.23445 | 5.27608 | 28.75140 | 7.82148 | 0.90570 |
| ant(6)-Ia_5_AB247327 | 0.48811 | 0.95339 | 0.00000 | 8.81428 | 1.48915 | 1.93334 | 0.62975 | 0.00000 |
| ant(6)-Ib_1_FN594949 | 0.00000 | 0.40403 | 0.00000 | 0.00000 | 0.00000 | 1.23974 | 0.00000 | 0.00000 |
| ant(9)-Ia_1_X02588 | 0.00000 | 0.00000 | 0.00000 | 0.18605 | 0.00000 | 0.00000 | 0.00000 | 0.00000 |
| aph(2__)-Id_1_AF016483 | 0.00000 | 0.00000 | 0.00000 | 0.00000 | 0.00000 | 0.00000 | 0.00000 | 1.41826 |
| aph(2__)-If_1_KF652097 | 0.00000 | 0.00000 | 0.00000 | 0.00000 | 0.00000 | 0.00000 | 2.07219 | 0.00000 |
| aph(3__)-Ib_1_M28829 | 90.62662 | 48.55954 | 20.08600 | 261.54656 | 70.94466 | 79.32271 | 92.72321 | 33.69524 |
| aph(3_)-Ia_2_EU287476 | 102.70608 | 284.52367 | 38.85993 | 178.34610 | 408.76846 | 209.75786 | 177.26900 | 1.70591 |
| aph(3_)-Ib_2_AJ744860 | 1.93329 | 0.00000 | 0.00000 | 0.00000 | 0.00000 | 1.53025 | 0.70152 | 0.00000 |
| aph(3_)-IIa_2_V00618 | 3.41062 | 2.23472 | 3.97939 | 1.14525 | 4.69707 | 0.87260 | 2.32632 | 0.00000 |
| aph(3_)-IIb_1_X90856 | 0.00000 | 5.47687 | 0.00000 | 1.08309 | 0.00000 | 0.00000 | 0.00000 | 0.00000 |
| aph(3_)-IIc_1_AM743169 | 0.00000 | 0.79948 | 0.00000 | 0.00000 | 0.00000 | 0.00000 | 0.00000 | 0.00000 |
| aph(3_)-III_1_M26832 | 1.17821 | 2.45274 | 0.64781 | 9.07040 | 3.28795 | 5.70096 | 1.77244 | 0.35917 |
| aph(3_)-VI_1_KC170992 | 3.91863 | 18.27709 | 0.42446 | 0.51360 | 0.71811 | 24.72468 | 4.34694 | 0.86944 |
| aph(3_)-XV_2_GQ626879 | 0.00000 | 0.65079 | 1.88765 | 0.45581 | 0.00000 | 0.00000 | 0.00000 | 0.00000 |
| aph(6)-Ic_1_X01702 | 5.72384 | 5.24741 | 0.00000 | 3.77376 | 0.00000 | 2.65592 | 3.79319 | 0.00000 |
| aph(6)-Id_2_AF024602 | 176.37927 | 104.23454 | 36.52218 | 542.84383 | 144.88907 | 148.31322 | 199.62756 | 69.33660 |
| ARR-3_4_FM207631 | 1.72501 | 0.55860 | 0.33873 | 0.00000 | 0.60173 | 11.49803 | 2.67610 | 0.00000 |
| ARR-6_3_JF922883 | 5.98552 | 0.28696 | 0.00000 | 0.72356 | 2.57600 | 44.61414 | 10.30375 | 0.00000 |

| | | | | | | | | |
|---|---|---|---|---|---|---|---|---|
| **blaACI-1_1_AJ007350** | 43.53296 | 3.04082 | 0.00000 | 0.00000 | 15.83202 | 2.97499 | 4.37764 | 0.00000 |
| **blaACT-14_1_JX440354** | 0.00000 | 0.00000 | 0.00000 | 0.00000 | 0.00000 | 0.00000 | 0.00000 | 0.00000 |
| **blaACT-6_1_FJ237366** | 14.67802 | 0.00000 | 0.00000 | 0.00000 | 6.72456 | 9.77193 | 0.00000 | 0.00000 |
| **blaACT-9_1_HQ693810** | 0.00000 | 0.00000 | 0.00000 | 0.00000 | 0.00000 | 8.45648 | 0.00000 | 0.00000 |
| **blaADC-25_1_EF016355** | 0.00000 | 0.00000 | 0.00000 | 0.00000 | 0.52674 | 0.00000 | 0.00000 | 0.99147 |
| **blaAER-1_1_U14748** | 0.00000 | 1.04186 | 0.00000 | 0.00000 | 0.00000 | 0.00000 | 0.00000 | 1.17026 |
| **blaBEL-2_1_FJ666063** | 0.00000 | 20.14009 | 2.84964 | 0.00000 | 5.42376 | 0.00000 | 7.39067 | 0.00000 |
| **blaBKC-1_1_KP689347** | 114.55980 | 21.25186 | 34.79459 | 169.25953 | 11.44631 | 51.30441 | 125.97821 | 0.00000 |
| **blaCARB-16_1_HF953351** | 3.46247 | 0.00000 | 0.00000 | 0.00000 | 0.00000 | 34.95639 | 3.82904 | 11.34058 |
| **blaCARB-2_1_M69058** | 2.38818 | 2.99873 | 0.00000 | 7.93909 | 7.75261 | 3.36055 | 7.00886 | 0.00000 |
| **blaCMY-93_1_KF992025** | 0.00000 | 0.00000 | 0.00000 | 20.62473 | 0.00000 | 0.00000 | 0.00000 | 0.00000 |
| **blaCMY-94_1_JX514368** | 0.00000 | 0.00000 | 10.94796 | 3.49635 | 16.58032 | 5.77191 | 10.56852 | 0.00000 |
| **blaCTX-M-60_1_AM411407** | 182.31766 | 2.67083 | 0.00000 | 32.92303 | 0.00000 | 3.80074 | 5.42822 | 0.00000 |
| **blaCTX-M-83_1_FJ214366** | 13.67513 | 0.00000 | 2.26760 | 16.46308 | 6.71371 | 0.00000 | 10.85748 | 0.00000 |
| **blaDES-1_1_AF426161** | 0.00000 | 0.00000 | 0.00000 | 0.00000 | 0.00000 | 0.00000 | 1.03875 | 0.00000 |
| **blaEBR-1_1_AF416700** | 0.00000 | 0.00000 | 0.00000 | 0.00000 | 0.00000 | 0.00000 | 0.00000 | 59.99243 |
| **blaFOX-5_1_AY007369** | 0.00000 | 0.00000 | 7.39566 | 19.17625 | 25.91822 | 52.68766 | 41.73452 | 55.69844 |
| **blaIMP-13_1_AJ550807** | 39.08814 | 40.08300 | 2.77781 | 257.82075 | 52.87058 | 33.17655 | 292.60959 | 0.00000 |
| **blaIMP-6_1_AB753460** | 152.75347 | 8344.83416 | 90.35207 | 3540.85297 | 111.11812 | 540.86614 | 2743.77219 | 0.00000 |
| **blaIMP-68_1_MF669572** | 0.00000 | 0.00000 | 0.00000 | 8.96519 | 0.00000 | 0.00000 | 0.00000 | 0.00000 |
| **blaKPC-28_1_KY282958** | 0.00000 | 0.00000 | 0.00000 | 0.00000 | 0.00000 | 43.62886 | 0.00000 | 0.00000 |
| **blaLCR-1_1_X56809** | 0.00000 | 0.00000 | 0.00000 | 0.00000 | 2.26530 | 0.47252 | 0.00000 | 0.00000 |
| **blaLEN7_1_AJ635425** | 10.91318 | 0.00000 | 0.00000 | 0.00000 | 0.00000 | 3.59704 | 0.00000 | 0.00000 |
| **blaMOX-4_1_FJ262599** | 1.67333 | 5.77002 | 0.00000 | 0.28527 | 0.48749 | 0.00000 | 0.84312 | 16.40199 |
| **blaOKP-B-8_1_AM051157** | 0.00000 | 0.00000 | 0.00000 | 0.00000 | 0.00000 | 1.18653 | 3.08109 | 0.00000 |
| **blaOXA-18_1_U85514** | 0.00000 | 15.49056 | 0.00000 | 0.00000 | 0.00000 | 6.70255 | 8.50897 | 0.00000 |
| **blaOXA-198_1_HQ634775** | 106.97053 | 4.61326 | 0.00000 | 5.90828 | 4.25951 | 0.00000 | 2.67887 | 0.00000 |
| **blaOXA-209_1_JF268688** | 0.00000 | 0.00000 | 1.38227 | 0.00000 | 0.00000 | 0.00000 | 15.31847 | 0.00000 |
| **blaOXA-210_1_JF795487** | 231.74254 | 359.77401 | 49.62260 | 186.25233 | 160.23349 | 94.24912 | 581.22010 | 12.94298 |
| **blaOXA-233_1_KJ657570** | 356.90019 | 196.21192 | 37.05201 | 75.05037 | 55.48289 | 247.37532 | 176.69224 | 11.29885 |
| **blaOXA-275_1_APPJ01000001** | 0.00000 | 0.00000 | 0.00000 | 4.43399 | 0.00000 | 1.91332 | 2.34223 | 14.38689 |
| **blaOXA-285_1_APRY01000059** | 0.00000 | 0.00000 | 4.84388 | 0.00000 | 0.00000 | 0.00000 | 0.00000 | 1.94034 |
| **blaOXA-296_1_APOH01000009** | 0.00000 | 20.77895 | 62.38595 | 0.51114 | 1.31024 | 0.59500 | 8.18877 | 16.74021 |
| **blaOXA-299_1_APQD01000016** | 0.00000 | 1.87043 | 0.48519 | 0.00000 | 0.00000 | 0.00000 | 2.11194 | 0.00000 |
| **blaOXA-334_1_KF203108** | 86.55822 | 488.30516 | 448.24214 | 409.78822 | 278.44336 | 350.13284 | 487.78820 | 235.88505 |
| **blaOXA-347_1_JN086160** | 7.76831 | 4.98971 | 2.80913 | 3.57569 | 3.28155 | 4.37250 | 34.42654 | 0.00000 |
| **blaOXA-37_1_AY007784** | 5.66230 | 0.00000 | 0.00000 | 0.00000 | 0.00000 | 4.15709 | 1.53926 | 1.42594 |
| **blaOXA-427_1_KX827604** | 10.23185 | 11.99117 | 0.00000 | 10.58213 | 10.33356 | 37.11444 | 20.10606 | 65.68406 |
| **blaOXA-437_1_KP410856** | 0.00000 | 46.15769 | 0.26657 | 11.56788 | 79.48649 | 404.72246 | 27.12236 | 0.00000 |
| **blaOXA-444_1_CP010800** | 0.00000 | 0.00000 | 0.00000 | 0.84799 | 0.00000 | 0.00000 | 0.00000 | 0.00000 |
| **blaOXA-45_1_AJ519683** | 0.00000 | 0.00000 | 12.58597 | 2.93185 | 0.00000 | 0.00000 | 0.00000 | 0.00000 |
| **blaOXA-46_1_AF317511** | 0.67701 | 0.00000 | 2.29627 | 2.72802 | 0.00000 | 0.00000 | 0.00000 | 1.38138 |
| **blaOXA-47_1_AY237830** | 19.99246 | 1.92933 | 8.41080 | 14.06800 | 22.69263 | 27.77083 | 63.95784 | 0.68723 |
| **blaOXA-486_1_AY597426** | 0.00000 | 0.00000 | 0.00000 | 0.00000 | 0.00000 | 0.00000 | 4.01831 | 0.00000 |
| **blaOXA-490_1_KU721147** | 0.00000 | 0.00000 | 0.00000 | 0.00000 | 0.00000 | 0.00000 | 0.98238 | 5.66778 |
| **blaOXA-5_1_AF347074** | 4.66010 | 1.67075 | 0.00000 | 23.59989 | 0.46445 | 0.69026 | 2.19074 | 2.75244 |
| **blaOXA-552_1_KY682754** | 0.00000 | 0.00000 | 0.00000 | 0.00000 | 0.00000 | 0.00000 | 0.00000 | 8.37443 |
| **blaOXA-58_1_AY665723** | 1147.38412 | 2331.53401 | 195.32032 | 2250.98072 | 2650.79821 | 2285.38912 | 3215.57473 | 118.12978 |
| **blaOXA-9_1_KQ089875** | 3.46586 | 0.00000 | 0.00000 | 0.00000 | 0.00000 | 1.79385 | 0.00000 | 0.00000 |
| **blaOXY-3-1_1_AF491278** | 0.00000 | 0.00000 | 0.00000 | 0.00000 | 0.00000 | 0.00000 | 0.00000 | 0.00000 |
| **blaOXY-6-4_4_AJ871877** | 11.16202 | 0.00000 | 9.25439 | 85.66487 | 0.00000 | 16.87020 | 14.40104 | 0.00000 |
| **blaPAO_2_FJ666065** | 0.00000 | 0.00000 | 0.00000 | 0.00000 | 0.00000 | 0.00000 | 9.29360 | 0.00000 |
| **blaPER-7_1_HQ713678** | 0.00000 | 0.00000 | 0.00000 | 0.00000 | 0.00000 | 11.02797 | 0.00000 | 0.00000 |
| **blaPLA-4A_1_AY507664** | 0.00000 | 0.00000 | 0.00000 | 0.00000 | 0.00000 | 0.00000 | 0.00000 | 0.00000 |
| **blaRAHN-1_1_GU645205** | 0.00000 | 0.00000 | 0.00000 | 0.00000 | 0.00000 | 0.00000 | 0.00000 | 0.96468 |
| **blaSGM-5_1_NG049987** | 0.00000 | 0.00000 | 0.00000 | 0.00000 | 0.00000 | 1.17858 | 0.00000 | 0.00000 |
| **blaSGM-6_1_NG049988** | 5.92994 | 2.12021 | 0.00000 | 2.15321 | 0.00000 | 0.00000 | 2.15457 | 0.00000 |

| | | | | | | | | |
|---|---|---|---|---|---|---|---|---|
| **blaSHV-129_1_GU827715** | 105.52083 | 1.05625 | 0.55510 | 3.84689 | 2.11305 | 9.01859 | 31.84311 | 0.00000 |
| **blaSST-1_1_AB008455** | 0.00000 | 0.00000 | 0.00000 | 10.89047 | 0.00000 | 0.00000 | 0.00000 | 0.00000 |
| **blaTEM-4_1_LK391770** | 185.48938 | 27.23164 | 42.39038 | 674.22610 | 356.23829 | 119.31872 | 136.77908 | 11.19482 |
| **blaTER-2_1_FJ263090** | 0.00000 | 0.00000 | 0.00000 | 0.00000 | 0.00000 | 0.00000 | 0.00000 | 0.00000 |
| **blaVEB-1_1_HM370393** | 0.00000 | 0.00000 | 0.00000 | 0.00000 | 0.00000 | 10.68834 | 24.16974 | 0.00000 |
| **blaVIM-42_1_KP071470** | 297.29205 | 121.94347 | 42.43807 | 63.02172 | 93.82729 | 146.31697 | 167.79185 | 18.09287 |
| **cat_2_M35190** | 7.34744 | 3.19432 | 1.29695 | 0.00000 | 0.89764 | 16.45348 | 1.76418 | 0.57200 |
| **cat(pC194)_1_NC_002013** | 0.00000 | 0.00000 | 0.28254 | 0.00000 | 0.00000 | 0.00000 | 0.00000 | 0.00000 |
| **cat(pC233)_1_AY355285** | 0.00000 | 0.00000 | 0.00000 | 0.00000 | 2.80927 | 0.35685 | 0.00000 | 0.00000 |
| **catA1_1_V00622** | 14.86437 | 0.00000 | 0.61310 | 1.71059 | 1.06084 | 2.31238 | 1.86811 | 0.00000 |
| **catA2_1_X53796** | 12.51670 | 0.00000 | 0.00000 | 0.00000 | 0.00000 | 0.00000 | 0.00000 | 0.00000 |
| **catB1_1_M58472** | 0.00000 | 0.00000 | 0.00000 | 0.00000 | 0.00000 | 0.00000 | 0.00000 | 0.00000 |
| **catB2_1_AF047479** | 0.00000 | 0.00000 | 0.00000 | 0.00000 | 0.58992 | 0.29224 | 0.00000 | 0.00000 |
| **catB3_2_U13880** | 4.12771 | 2.66973 | 0.17434 | 1.84109 | 1.69601 | 3.43387 | 14.46929 | 0.33832 |
| **catB9_1_AF462019** | 0.00000 | 0.00000 | 0.00000 | 0.00000 | 0.00000 | 0.00000 | 0.00000 | 0.33993 |
| **catP_1_U15027** | 0.00000 | 0.00000 | 0.00000 | 0.00000 | 0.00000 | 0.88938 | 3.88120 | 0.00000 |
| **catQ_1_M55620** | 3.95885 | 0.00000 | 0.00000 | 0.00000 | 0.00000 | 0.00000 | 0.00000 | 0.00000 |
| **catS_1_X74948** | 5.21045 | 1.76145 | 1.27107 | 0.37011 | 0.75898 | 3.57197 | 1.61100 | 0.00000 |
| **cepA_6_FR688022** | 8.78973 | 0.00000 | 0.00000 | 0.00000 | 0.00000 | 0.30729 | 0.00000 | 0.00000 |
| **cfr(C)_2_CANB01000378** | 92.32861 | 33.39631 | 13.32525 | 0.00000 | 30.21875 | 126.89133 | 44.11879 | 7.70253 |
| **cfxA4_1_AY769933** | 15.36126 | 28.52895 | 6.05487 | 2.03585 | 12.51489 | 25.03882 | 6.83757 | 0.77593 |
| **cfxA6_1_GQ342996** | 24.94644 | 29.19235 | 8.64254 | 1.97453 | 3.98349 | 36.35731 | 6.27793 | 1.11093 |
| **cmlA1_1_M64556** | 0.00000 | 0.00000 | 0.00000 | 0.00000 | 3.88976 | 0.33034 | 0.00000 | 0.00000 |
| **cmlB1_1_AM296481** | 42.28959 | 91.38679 | 11.82621 | 25.05606 | 197.73276 | 57.82778 | 68.83347 | 0.78942 |
| **cmx_1_U85507** | 0.79650 | 0.40531 | 1.15739 | 2.16780 | 0.00000 | 0.00000 | 13.92910 | 0.00000 |
| **cphA1_4_AY261376** | 0.00000 | 0.00000 | 0.00000 | 0.00000 | 0.00000 | 1.69272 | 1.15121 | 1.58635 |
| **cphA6_1_AY227052** | 0.00000 | 0.00000 | 0.00000 | 0.00000 | 0.00000 | 1.26954 | 1.09365 | 1.25975 |
| **dfrA1_5_EU089668** | 2.39214 | 40.31495 | 1.00890 | 55.70420 | 0.00000 | 4.19546 | 15.23537 | 0.22590 |
| **dfrA10_1_L06418** | 0.00000 | 0.00000 | 0.91313 | 0.00000 | 0.00000 | 0.00000 | 0.00000 | 0.00000 |
| **dfrA12_1_FJ763641** | 0.00000 | 0.00000 | 0.22160 | 0.80444 | 0.00000 | 0.00000 | 0.00000 | 0.00000 |
| **dfrA14_2_Z50805** | 3.52283 | 0.00000 | 0.00000 | 0.68888 | 0.00000 | 0.58320 | 0.00000 | 0.60013 |
| **dfrA16_3_AY878718** | 16.64099 | 0.27425 | 0.00000 | 0.30733 | 0.39390 | 0.68298 | 3.43725 | 0.00000 |
| **dfrA17_8_AM932673** | 13.72881 | 0.27425 | 0.00000 | 0.69150 | 3.44663 | 0.48784 | 0.00000 | 0.30121 |
| **dfrA26_1_AM403715** | 0.00000 | 0.00000 | 0.00000 | 0.00000 | 0.00000 | 0.00000 | 0.00000 | 0.00000 |
| **dfrA28_2_FM877476** | 0.00000 | 0.00000 | 0.00000 | 0.00000 | 0.00000 | 2.14651 | 0.00000 | 0.00000 |
| **dfrA5_2_FJ001870** | 1.66410 | 2.28543 | 0.62086 | 0.30733 | 34.17084 | 2.53679 | 14.67798 | 0.00000 |
| **dfrA7_2_AJ884724** | 0.31202 | 0.00000 | 0.00000 | 0.00000 | 0.00000 | 0.00000 | 0.00000 | 0.00000 |
| **dfrA7_5_AJ419170** | 3.54606 | 4.94134 | 0.00000 | 5.04755 | 4.09449 | 0.00000 | 4.09438 | 1.31500 |
| **dfrB1_1_U36276** | 0.00000 | 3.06933 | 0.00000 | 0.00000 | 0.00000 | 0.00000 | 0.00000 | 0.00000 |
| **dfrB2_1_J01773** | 0.00000 | 0.00000 | 0.00000 | 0.00000 | 0.00000 | 0.00000 | 0.00000 | 0.30121 |
| **dfrB3_2_FM877478** | 0.00000 | 0.00000 | 0.00000 | 0.00000 | 0.00000 | 0.00000 | 0.00000 | 0.45181 |
| **dfrG_1_AB205645** | 0.00000 | 1.56621 | 0.44321 | 0.00000 | 9.84157 | 2.41453 | 0.61895 | 0.00000 |
| **ere(A)_5_FN396877** | 30.58136 | 14.05638 | 8.28437 | 5.41844 | 1.21437 | 3.64717 | 17.00498 | 3.07598 |
| **ere(D)_1_KP265721** | 6.79015 | 4.97944 | 2.69825 | 20.95504 | 3.38571 | 6.52065 | 59.32201 | 12.01402 |
| **erm(A)_1_X03216** | 4.44498 | 0.00000 | 0.00000 | 1.79110 | 0.00000 | 0.00000 | 0.00000 | 0.00000 |
| **erm(A)_2_AF002716** | 0.00000 | 0.00000 | 0.00000 | 0.00000 | 0.00000 | 0.00000 | 1.44374 | 0.00000 |
| **erm(B)_9_AF299292** | 121.08273 | 94.83569 | 126.13263 | 151.44252 | 48.05188 | 258.66673 | 156.33775 | 115.44040 |
| **erm(F)_1_M14730** | 284.71511 | 281.08812 | 73.29685 | 84.29571 | 74.12405 | 155.89083 | 842.41715 | 10.96189 |
| **erm(G)_2_L42817** | 2.41464 | 11.67303 | 1.50148 | 0.00000 | 1.90519 | 22.65189 | 2.87569 | 0.00000 |
| **erm(T)_3_AF310974** | 0.00000 | 0.00000 | 0.00000 | 0.00000 | 29.72097 | 0.00000 | 7.90814 | 1.45686 |
| **erm(X)_1_M36726** | 0.00000 | 0.00000 | 0.00000 | 18.65676 | 2.62047 | 0.00000 | 0.00000 | 0.00000 |
| **erm(X)_3_U21300** | 0.00000 | 0.00000 | 0.00000 | 38.42637 | 12.86379 | 0.00000 | 0.00000 | 0.00000 |
| **floR_2_AF118107** | 34.97586 | 14.83621 | 16.95496 | 21.40180 | 17.97935 | 74.68133 | 30.51570 | 1.35134 |
| **fosA_1_M85195** | 0.00000 | 0.00000 | 0.43176 | 1.28236 | 2.08185 | 0.00000 | 1.86059 | 2.34602 |
| **fosA_2_AGDM01000012** | 0.00000 | 0.00000 | 0.00000 | 0.00000 | 0.00000 | 1.21125 | 0.62906 | 0.00000 |
| **fosA_4_ACWU01000146** | 0.00000 | 0.95585 | 0.00000 | 0.00000 | 0.00000 | 0.00000 | 0.00000 | 0.00000 |
| **fosA_6_ACZD01000244** | 23.33124 | 0.71775 | 0.95752 | 2.67153 | 0.00000 | 2.29815 | 12.08693 | 2.28043 |

| | | | | | | | | |
|---|---|---|---|---|---|---|---|---|
| **fosA7_1_LAPJ01000014** | 5.47766 | 0.00000 | 0.00000 | 0.86097 | 0.00000 | 0.00000 | 0.00000 | 0.00000 |
| **fosE_1_AB901041** | 0.00000 | 0.85594 | 0.00000 | 0.00000 | 1.61353 | 0.00000 | 0.00000 | 0.00000 |
| **fosE_2_AY029772** | 0.00000 | 0.00000 | 0.00000 | 0.00000 | 0.52299 | 0.00000 | 0.00000 | 0.00000 |
| **imiS_1_Y10415** | 0.00000 | 0.00000 | 0.67268 | 0.00000 | 0.00000 | 0.84570 | 2.47317 | 1.39863 |
| **lnu(B)_1_AJ238249** | 5.27327 | 0.00000 | 0.00000 | 0.00000 | 3.13503 | 1.03539 | 0.00000 | 5.06094 |
| **lnu(C)_1_AY928180** | 4.08334 | 6.30279 | 0.59452 | 1.10361 | 0.75438 | 9.34295 | 2.57977 | 0.57686 |
| **lnu(G)_1_KX470419** | 0.00000 | 0.00000 | 0.00000 | 0.00000 | 0.34834 | 0.00000 | 0.00000 | 1.64259 |
| **lsa(A)_3_AY737526** | 0.00000 | 0.00000 | 0.00000 | 0.00000 | 3.89756 | 0.00000 | 0.00000 | 0.00000 |
| **lsa(E)_1_JX560992** | 31.87001 | 0.00000 | 0.00000 | 0.00000 | 9.61833 | 3.92404 | 0.00000 | 17.73833 |
| **mcr-3.11_1_MG489958** | 0.00000 | 0.00000 | 0.00000 | 0.00000 | 0.00000 | 0.00000 | 0.00000 | 0.72490 |
| **mcr-3.17_1_MH332767** | 0.00000 | 0.00000 | 0.00000 | 0.00000 | 0.00000 | 0.00000 | 0.00000 | 0.81370 |
| **mcr-4.1_1_MF543359** | 0.57606 | 0.00000 | 0.00000 | 0.00000 | 0.94732 | 0.00000 | 0.00000 | 4.41223 |
| **mcr-5.2_1_MG384740** | 0.87042 | 0.00000 | 0.00000 | 0.00000 | 0.00000 | 0.00000 | 0.00000 | 0.00000 |
| **mcr-9_1_NZ_NAAN01000063.1** | 1.67373 | 0.00000 | 0.00000 | 0.85427 | 0.31694 | 0.00000 | 0.57081 | 0.44065 |
| **mdf(A)_1_Y08743** | 145.61773 | 14.97105 | 8.59240 | 16.30438 | 129.46953 | 35.55777 | 52.06933 | 0.00000 |
| **mef(A)_1_AJ971089** | 25.49944 | 33.22810 | 8.03377 | 0.00000 | 1.07304 | 13.06172 | 2.81991 | 56.96810 |
| **mef(B)_1_FJ196385** | 10.18043 | 0.00000 | 0.00000 | 0.00000 | 0.00000 | 0.00000 | 0.00000 | 0.00000 |
| **mef(C)_1_AB571865** | 7.45123 | 21.87827 | 12.29212 | 32.90804 | 3.05080 | 5.21419 | 49.89794 | 6.24044 |
| **mph(A)_1_D16251** | 89.02101 | 2.29572 | 6.98369 | 8.36110 | 20.40194 | 24.29786 | 34.21622 | 4.09721 |
| **mph(A)_2_U36578** | 2.99755 | 0.00000 | 0.00000 | 0.00000 | 0.00000 | 0.00000 | 0.00000 | 0.00000 |
| **mph(E)_1_DQ839391** | 97.48374 | 668.92414 | 39.52958 | 252.01140 | 319.72544 | 393.49663 | 574.43149 | 265.13631 |
| **mph(F)_1_AM260957** | 0.00000 | 0.00000 | 1.18533 | 0.00000 | 0.00000 | 0.00000 | 0.00000 | 0.00000 |
| **mph(G)_1_AB571865** | 6.57319 | 20.41732 | 10.51628 | 29.34098 | 1.89873 | 4.49412 | 50.15391 | 6.65462 |
| **msr(C)_2_AF313494** | 1.83329 | 0.00000 | 0.00000 | 29.17954 | 52.23172 | 2.34521 | 5.50796 | 0.00000 |
| **msr(D)_3_AF227520** | 32.93329 | 34.62985 | 19.44844 | 0.00000 | 9.56499 | 37.52880 | 7.76008 | 180.51284 |
| **msr(E)_1_FR751518** | 652.64288 | 4246.86017 | 261.19198 | 1759.01904 | 1986.94219 | 2736.78705 | 3407.26054 | 1397.12855 |
| **nimA_1_X71444** | 0.00000 | 0.00000 | 0.00000 | 1.50889 | 0.00000 | 2.09029 | 0.00000 | 0.00000 |
| **nimE_1_AM042593** | 0.00000 | 0.00000 | 0.00000 | 0.00000 | 0.00000 | 0.00000 | 0.00000 | 0.23982 |
| **oqxA_1_EU370913** | 31.85985 | 1.47387 | 2.12709 | 6.00790 | 3.25470 | 4.64049 | 11.68247 | 2.73161 |
| **oqxB_1_EU370913** | 69.10917 | 28.69541 | 24.17412 | 39.96505 | 33.10186 | 24.34857 | 38.46162 | 28.25553 |
| **poxtA_1_MF095097** | 0.00000 | 0.00000 | 0.00000 | 22.35667 | 28.22407 | 19.58923 | 0.00000 | 24.75940 |
| **qepA_1_FJ167861** | 0.00000 | 4.48551 | 0.00000 | 0.00000 | 0.00000 | 0.00000 | 0.00000 | 0.00000 |
| **qnrA6_1_DQ151889** | 1.42569 | 0.00000 | 0.00000 | 0.44346 | 0.00000 | 0.35196 | 1.27343 | 0.00000 |
| **qnrB43_1_JQ349151** | 0.00000 | 0.00000 | 0.00000 | 1.41177 | 0.00000 | 1.21908 | 0.40967 | 0.00000 |
| **qnrB60_1_AB734055** | 0.00000 | 0.00000 | 0.22813 | 0.00000 | 0.00000 | 0.00000 | 0.00000 | 0.00000 |
| **qnrS5_1_HQ631377** | 0.00000 | 0.00000 | 0.00000 | 0.00000 | 0.00000 | 0.00000 | 0.00000 | 0.97806 |
| **qnrVC1_1_EU436855** | 0.37518 | 0.32977 | 0.00000 | 0.00000 | 0.00000 | 0.00000 | 0.00000 | 0.00000 |
| **str_2_FN435330** | 0.46454 | 0.00000 | 0.17332 | 0.00000 | 0.00000 | 0.00000 | 0.67425 | 1.34532 |
| **sul1_38_BX248359** | 157.46302 | 952.21802 | 27.75928 | 408.51686 | 106.16778 | 234.41568 | 384.53112 | 11.73083 |
| **sul2_12_AF497970** | 56.07602 | 112.91549 | 9.89021 | 137.47959 | 49.68677 | 74.26624 | 103.87082 | 3.77772 |
| **tet(32)_1_EU722333** | 20.72095 | 13.20263 | 2.64401 | 2.21928 | 2.91732 | 31.93976 | 26.97079 | 2.50966 |
| **tet(33)_1_AY255627** | 0.00000 | 0.00000 | 1.86335 | 0.00000 | 0.00000 | 0.00000 | 1.87072 | 0.00000 |
| **tet(36)_1_AJ514254** | 0.00000 | 0.00000 | 1.20516 | 0.00000 | 0.00000 | 0.00000 | 0.00000 | 0.00000 |
| **tet(39)_1_KT346360** | 71.35601 | 133.70267 | 33.76986 | 545.31157 | 78.71052 | 326.12398 | 712.15679 | 362.91041 |
| **tet(40)_2_AM419751** | 40.05285 | 11.92421 | 8.61659 | 9.18678 | 17.12643 | 108.78226 | 21.49405 | 1.22777 |
| **tet(42)_1_EU523697** | 3.29426 | 6.19505 | 0.00000 | 35.59839 | 6.09306 | 0.00000 | 13.13833 | 0.00000 |
| **tet(44)_2_FN594949** | 6.22966 | 3.17721 | 0.00000 | 0.00000 | 0.00000 | 3.89605 | 3.84696 | 0.00000 |
| **tet(A)_3_AY196695** | 26.50960 | 15.72450 | 4.12612 | 38.86698 | 12.24193 | 27.53878 | 27.01959 | 19.54650 |
| **tet(B)_1_AP000342** | 4.41483 | 4.09603 | 0.00000 | 5.25449 | 0.00000 | 7.93802 | 7.88668 | 0.00000 |
| **tet(C)_2_AY046276** | 56.19911 | 7.45335 | 22.08413 | 27.20679 | 5.90352 | 9.04675 | 13.81166 | 15.46896 |
| **tet(D)_1_AF467077** | 0.00000 | 1.46267 | 0.00000 | 0.00000 | 0.00000 | 0.78055 | 1.18910 | 0.00000 |
| **tet(E)_1_Y19116** | 0.00000 | 0.00000 | 0.00000 | 0.00000 | 0.00000 | 0.76400 | 1.16389 | 6.54502 |
| **tet(G)_2_AF133140** | 32.97755 | 15.15506 | 21.04745 | 24.79461 | 12.67971 | 14.52599 | 30.95085 | 2.90877 |
| **tet(H)_2_AJ245947** | 3.70669 | 0.50680 | 0.00000 | 0.00000 | 0.00000 | 2.00909 | 0.00000 | 1.55057 |
| **tet(L)_7_X60828** | 0.00000 | 0.00000 | 0.00000 | 0.52896 | 18.77934 | 14.97926 | 0.00000 | 0.00000 |
| **tet(M)_9_X56353** | 34.25904 | 4.80443 | 29.46999 | 5.91479 | 61.52119 | 45.06627 | 11.82752 | 12.09540 |
| **tet(O)_3_Y07780** | 73.71729 | 49.62835 | 21.78433 | 17.46975 | 45.94784 | 217.00247 | 55.59285 | 6.19050 |

| | | | | | | | | |
|---|---|---|---|---|---|---|---|---|
| **tet(O/32/O)_2_AJ295238** | 8.55028 | 5.95811 | 0.57478 | 1.99167 | 0.00000 | 10.76702 | 0.00000 | 0.00000 |
| **tet(O/W/32/O)_5_JQ740053** | 31.50509 | 21.39503 | 8.21942 | 4.60928 | 1.16693 | 69.15463 | 20.57211 | 1.28272 |
| **tet(Q)_2_X58717** | 197.34849 | 146.33057 | 52.16485 | 21.73006 | 43.54848 | 141.69939 | 51.25066 | 6.41671 |
| **tet(S/M)_2_AY534326** | 1.45447 | 0.00000 | 0.00000 | 20.07570 | 2.10192 | 0.00000 | 1.50427 | 1.10848 |
| **tet(W)_3_AJ427421** | 379.47901 | 184.95548 | 66.02108 | 49.73701 | 56.19076 | 990.86006 | 209.12869 | 17.55734 |
| **tet(X)_1_GU014535** | 85.84011 | 251.74723 | 34.67422 | 160.78047 | 31.35810 | 116.35218 | 268.65610 | 14.19155 |
| **tet(X)_3_AB097942** | 0.00000 | 0.00000 | 1.03532 | 0.00000 | 4.92635 | 0.00000 | 0.00000 | 0.00000 |
| **VanG2XY_1_FJ872410** | 0.68055 | 0.00000 | 0.00000 | 0.00000 | 0.00000 | 0.00000 | 0.72944 | 0.00000 |
| **VanHAX_2_M97297** | 0.00000 | 1.69537 | 0.02822 | 19.30613 | 58.11818 | 6.56372 | 6.18198 | 0.00000 |
| **VanHBX_1_AF192329** | 0.43493 | 0.00000 | 0.00000 | 0.00000 | 0.00000 | 5.00262 | 1.31747 | 0.00000 |
| **vat(F)_1_AF170730** | 0.00000 | 0.00000 | 0.00000 | 0.00000 | 0.00000 | 0.00000 | 0.00000 | 2.73325 |
